# A new SARS-CoV-2 variant poorly detected by RT-PCR on nasopharyngeal samples, with high lethality

**DOI:** 10.1101/2021.05.05.21256690

**Authors:** Pierre Fillatre, Marie-José Dufour, Sylvie Behillil, Remi Vatan, Pascale Reusse, Alice Gabellec, Nicolas Velmans, Catherine Montagne, Sophie Geffroy, Edith Droumaguet, Véronique Merour, Vincent Enouf, Rodolphe Buzele, Marion Valence, Elena Guillotel, Bertrand Gagniere, Artem Baidaluk, Anna Zhukova, Mathieu Tourdjman, Vincent Thibault, Claire Grolhier, Charlotte Pronier, Xavier Lescure, Etienne Simon-Loriere, Dominique Costagliola, Sylvie Van Der Werf, Pierre Tattevin, Nicolas Massart

## Abstract

**Background:** In early January 2021, an outbreak of nosocomial cases of COVID-19 emerged in Western France, with RT-PCR tests repeatedly negative on nasopharyngeal samples but positive on lower respiratory tract samples. Whole genome sequencing (WGS) revealed a new variant, currently defining a novel SARS-CoV-2 lineage: B.1.616. In March, WHO classified this variant as ‘under investigation’ (VUI). We analyzed the characteristics and outcomes of COVID-19 cases related to this new variant.

**Methods:** Clinical, virological, and radiological data were retrospectively collected from medical charts in the two hospitals involved. We enrolled patients with at least one of the following: i) positive SARS-CoV-2 RT-PCR on a respiratory sample; ii) seroconversion with anti-SARS-CoV-2 IgG/IgM; iii) suggestive symptoms and typical features of COVID-19 on chest CT scan. Cases were categorized as either: i) B.1.616; ii) variant of concern (VOC); iii) unknown.

**Findings:** From January 1^st^ to March 24^th^, 2021, 114 patients fulfilled the inclusion criteria: B.1.616 (n=34), VOC (n=32), and unknown (n=48). B.1.616-related cases were older than VOC-related cases (81 years [73-88], vs 73 years [67-82], *P*<0.05) and their first RT-PCR tests were less often positive (5/34, 15% vs 31/32, 97%, *P*<0.05). The B.1.616 variant was independently associated with severe disease (multivariable Cox model HR 4.2 [1.3– 13.5], *P*=0.018), and increased lethality (logrank test *P*=0.01): 28-day mortality 15/34 (44%) with B.1.616, vs. 5/32 (16%) for VOC, *P*=0.036.

**Interpretation:** We report a nosocomial outbreak of COVID-19 cases related to a new variant, B.1.616, poorly detected by RT-PCR on nasopharyngeal samples, with high lethality.

**Research in context:** *Evidence before this study:* Among the numerous SARS-CoV-2 variants described worldwide, only 3 are currently classified as Variant of Concern (VOC) by the WHO, since they are associated with either an increased risk in transmissibility, severity, or significant reduction in neutralization by antibodies: B.1.1.7, B.1.351 and P.1 (Pango lineage nomenclature). With the ongoing circulation of SARS-CoV-2 in many places worldwide, the emergence of new variants may reduce the efficacy of vaccines and jeopardize our prospects to control the pandemic. In early January 2021, an outbreak of cases highly suggestive of COVID-19 despite negative RT-PCR tests on repeated nasopharyngeal (NP) samples was reported in Western France, leading to several nosocomial clusters. Whole-genome sequencing (WGS) from lower respiratory tract samples identified a new lineage of SARS-CoV-2 virus, classified as B1.616. Consequently, the French public health agency (Santé publique France) and the WHO classified B.1.616 as ‘variant under investigation’ (VUI).

*Added value of this study:* Our observational study, conducted from January 1^st^ to March 24^th^ 2021 in the B.1.616 identified area, provides the first clinical and virological description of B.1.616-associated COVID-19. The 34 cases had clinical, biological and radiological findings in line with classical features of COVID-19, while RT-PCR tests on nasopharyngeal (NP) samples failed to detect SARS-CoV-2 in most patients. Indeed, this gold-standard test was positive in only 15% of the first tests in B.1.616-related COVID-19 patients. Of note, the diagnostic performance of RT-PCR tests was satisfactory on lower respiratory tract samples, suggesting that failure to detect B.1.616 on NP samples would be due to a viral load below the limit of detection in the upper respiratory tract, rather than to genomic mismatches between routine RT-PCR targets and this variant. In our cohort, B.1.616 was independently associated with worse clinical outcome, with high 28-day mortality (44%).

*Implications of all the available evidence:* Diagnosis of B.1.616-related COVID-19 cases should not rely on RT-PCR tests on NP samples. In the epidemic area, strict infection control measures must be maintained as long as COVID-19 diagnosis is not ruled out, in order to limit nosocomial clusters and case fatality. Further studies are needed to confirm and investigate the association between genomic characteristics of B.1.616, and i) poor detection by RT-PCR tests on NP samples; ii) prognosis.

## INTRODUCTION

Since the emergence of COVID-19, numerous SARS-CoV-2 variants have been described ^1^. At the end of 2020, novel concerns were raised with the detection of rapidly spreading variants of concern (VOCs) in distinct regions of the world. Those VOCs are characterized by multiple mutations and associated with increased transmissibility, increased severity and/or immune escape properties ^2–4^. The WHO issued working definitions for the classification of VOCs and variants under investigation (VUI), including viruses with decreased effectiveness of public health and social control measures ^5^. In January 2021, an outbreak of cases highly suggestive of COVID-19 despite negative RT-PCR tests on repeated nasopharyngeal samples was reported at the Lannion hospital (Côtes d’Armor, Western France). Of note, when applied on lower respiratory tract samples, the performance of RT-PCR tests was preserved, suggesting that failure to detect this variant on nasopharyngeal samples was due to a viral load below the limit of detection in the upper respiratory tract, rather than to genomic mismatches between routine RT-PCR targets and this variant, which was confirmed by the genomic data. Whole-genome sequencing (WGS) from lower respiratory tract samples of several cases identified an original variant of SARS-CoV-2 belonging to clade GH/20C (GISAID/Nextstrain nomenclatures) carrying a unique constellation of changes, including several amino acid substitutions or deletions in the Spike (S) protein, which received the B.1.616 Pango lineage designation. In March, the French public health agency (Santé publique France), the National Reference Center for respiratory viruses (Institut Pasteur, Paris), and the WHO classified B.1.616 as VUI ^5,6^. We aimed to characterize this variant in terms of virological features, clinical presentation, and outcomes.

## MATERIAL AND METHODS

### Setting and Patients

We conducted a retrospective observational study in two hospitals of the *Groupement Hospitalier du Territoire d’Armor*, a network of 6 public hospitals that serve as secondary care centers in the area. During the study period, COVID-19 patients who required hospitalization could be admitted either to the Lannion hospital, a 550 acute care beds hospital in an agglomeration of 50,000 inhabitants, or the Saint-Brieuc hospital, a 750 acute care beds hospital in an agglomeration of 150,000 inhabitants. Patients who required endotracheal intubation were transferred to the intensive care unit (ICU) of the Saint-Brieuc hospital.

We screened medical files of all patients who were diagnosed with SARS-CoV-2 infection at the Lannion and Saint-Brieuc hospitals from January 1^st^ to March 24^th^, 2021. SARS-CoV-2 infection was defined by at least one of the following: i) positive SARS-CoV-2 RT-PCR on a respiratory sample; ii) seroconversion based on paired sera tested for anti-SARS-CoV-2 IgG/IgM; iii) suggestive symptoms and typical features of COVID-19 on chest CT scan.

COVID-19 cases were categorized in one of the three following groups: i) B.1.616; ii) VOC; iii) unknown. The B.1.616 group included COVID-19 patients with B.1.616 infection documented by WGS (confirmed B.1.616 case), and COVID-19 patients for which the SARS-CoV-2 isolate could not be characterized, but who were close contacts of at least one patient with documented B.1.616 infection (probable B.1.616 case). Close contact was defined as either household contact, occupational, or nosocomial (hospitalization in the same ward during the same period). Since all cases with B.1.616 infection confirmed by WGS lived in the Lannion district, this place was considered as the epidemic area.

The VOC group included all COVID-19 cases due to VOCs B.1.1.7, B.1.351 and P.1. Routine screening for these VOCs has been performed by a prescreening RT-PCR in all patients with positive RT-PCR since February 10^th^, 2021 in the participating hospitals. COVID-19 patients not fulfilling the criteria for the first two groups were categorized in the ‘unknown’ group. These patients were included in the study for clinical, biological, virological, and radiological description. However, since it was not possible to assign them to either the B.1.616 group, or the VOC group, they were not included in the primary analysis. Cases were categorized as nosocomial COVID-19 if symptoms appeared at least 2 days after hospital admission ^7^.

### Management

All patients with COVID-19 confirmation or suspicion, based on clinical and/or radiological findings, were isolated with droplet transmission precautions for at least 10 days, including 48 h after clinical improvement and apyrexia. Patients who required ICU management were isolated during 14 days, and immunocompromised patients were isolated during 24 days, including 48 h after clinical improvement and apyrexia. Infection control measures included admission to a single room, wearing gown and gloves, protection glasses, surgical mask (if treated with O_2_ flow <6 L/min) or FFP2 mask (if treated with O_2_ flow ≥6 L/min) and reinforced hand hygiene. All COVID-19 patients received prophylactic therapy with heparin, according to national guidelines, and those who required treatment with O_2_ received dexamethasone, 6 mg daily for 10 days or until they no longer required O_2._ Tocilizumab could be prescribed at the discretion of the physicians in charge.

### Virological methods

<u>D</u>ifferent commercially available RT-PCR assay were used at Lannion and Saint-Brieuc hospitals: Xpert^®^ Xpress SARS-CoV-2, GeneXpert Cepheid^®^, BD SARS-CoV-2 reagents for BD Max, BD^®^, Viasure SARS-CoV-2 Real Time PCR Detection, Certest^®^. Screening for VOCs was performed on positive RT-PCR samples using ID SARS-CoV-2/UK/SA Variant Triplex, ID Solutions^®^. When B.1.616-related COVID-19 was suspected, samples were sent to the National Reference Center (Institut Pasteur, Paris) for WGS.

SARS-CoV-2 serology (IgM/IgG) of suspected COVID-19 cases was performed using AdviseDx SARS-CoV-2 Alinity Abbott^®^.

#### Whole Genome Sequencing

RNA extracted from clinical samples was used for SARS-CoV-2 WGS using a highly multiplexed PCR amplicon approach^8^ with the ARTIC Network multiplex PCR primers set v3 (https://artic.network/ncov-2019). Synthesized cDNA was used as template and amplicons were generated using two pooled primer mixtures for 35 rounds of amplification. Libraries were prepared using the Nextera XT DNA Library Prep Kit (Illumina) and sequenced on an Illumina NextSeq500 (2×150 cycles) on the Mutualized Platform for Microbiology (P2M) at Institut Pasteur.

#### Genome assembly and phylogenetic analysis

Illumina adaptors, low quality reads, primer sequences were trimmed with Trimmomatic v0.36 ^9^ and reads were assembled with Megahit ^10^ as well as mapped onto reference genome Wuhan/Hu1/2019 (NCBI Nucleotide – NC_045512, GenBank – MN908947) using the CLC Genomics Suite v5.1.0 (QIAGEN). SAMtools v1.3 ^11^ was used to sort the aligned bam files and generate alignment statistics. Aligned reads were manually inspected using Geneious prime v2020.1.2 (2020) (https://www.geneious.com/), and consensus sequences were generated (> 5X read-depth coverage for a base call). SARS-CoV-2 sequences generated during this study (n=6) were combined to a global dataset of sequences available on GISAID to produce a subsampled phylogenetic tree (n=3238) with augur and auspice as implemented in the Nextstrain pipeline version (April, 2021) (https://github.com/nextstrain/ncov).

### Outcomes

Primary outcome was clinical severity defined as a score >5 in the WHO clinical progression scale ^12^. Secondary outcomes were ICU admission and 28 day-mortality.

### Statistical analysis

Statistical analysis was performed with R 3.4.3. Categorical variables were expressed as numbers (percent) and continuous variables as medians (interquartile range [IQR]). The chi-square test and the Fisher’s exact test were used to compare categorical variables and the Mann-Whitney U test or Kruskall-Wallis test was used for continuous variables, when appropriate. Kaplan-Meier survival curves were compared with the log-rank test. Multivariable analysis was made using the Cox proportional-hazard regression model. Variables associated with outcome with a *P*-value <0.2 in univariate analysis or those considered clinically relevant were included in the multivariate analysis. All tests were two-sided, and a *P*-value ≤0.05 was considered statistically significant.

### Ethics

Patients or closest relatives were informed of the retrospective collection of data and could refuse to participate. The French infectious diseases society ethics committee (Comité d’Éthique de la Recherche en Maladies Infectieuses et Tropicales, CER-MIT) approved the study (N° COVID 2021-06). Written informed consent was waived.

## RESULTS

### Genomic characteristics of B.1.616

Phylogenetic analysis revealed an original variant carrying a unique constellation of mutations, which received the B.1.616 Pango lineage designation^1^ (Figure 1). It is characterized by 9 amino acid changes and one deletion in the S protein in comparison with the original Wuhan strain, several unique amino-acid changes in the E, M, and N proteins, in ORF1ab and ORF3 as well as by a deletion and frameshift in ORF6 and replacement of the stop codon of ORF7a resulting in a 5 amino acids extension at its C-terminus (Table 1). Interestingly, some mutations (Y144-, H655Y) in the S protein have been observed for previously described VOCs, i.e B.1.1.7 and P.1, respectively. The V483A is located in the receptor binding motif next to residue 484, for which the E484K change found in several VOCs has been associated with reduced neutralization by post-infection and post-vaccination antibodies^13–15^. Also of note are the mutations in ORF6 and ORF7a, two proteins that antagonize various steps of type I interferon (IFN-I) production and signaling^16–18^.

**Table 1:**
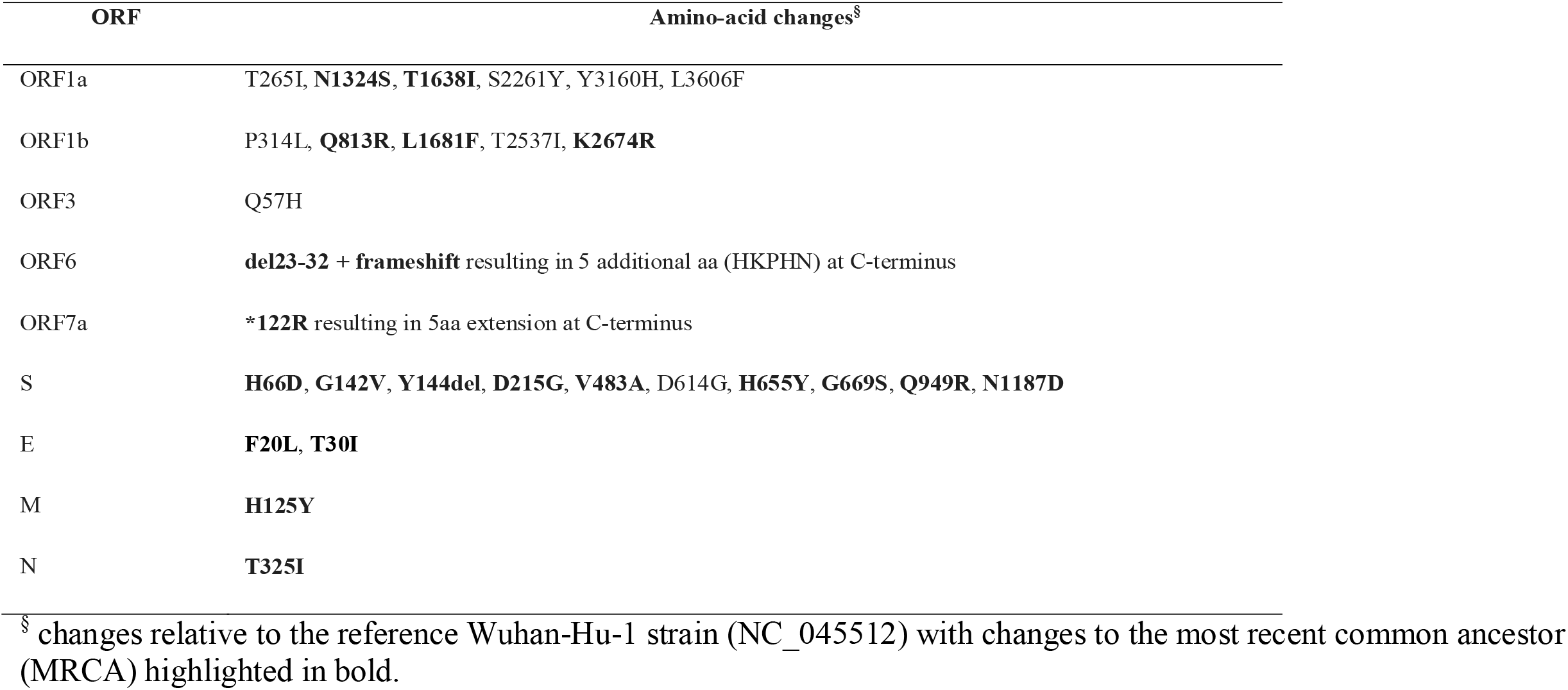
Amino-acid substitutions characteristic of the B.1.616 variant

**Figure 1:**
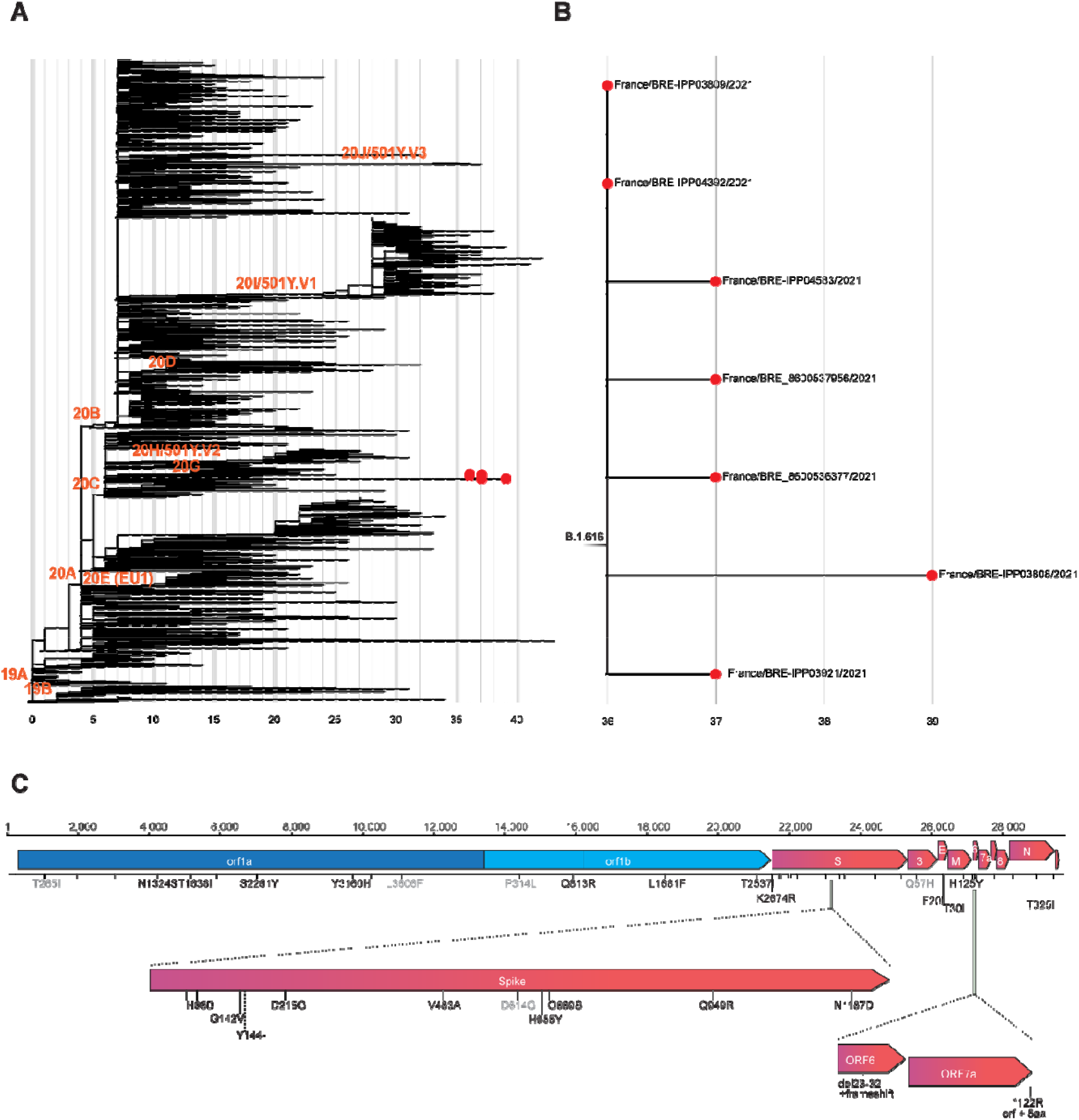
Phylogenetic analysis of the B.1.616 lineage and characteristic non-synonymous substitutions. A. Subsampled global phylogenetic maximum likelihood tree of SARS-CoV-2 with annotated Nextstrain clades next to the corresponding nodes and tips highlighted only for sequences from the Pangolin B.1.616 lineage. B. Detailed view of the B1.616 lineage. A-B. Branch lengths correspond to the number of nucleotide substitutions (shown below the tree) from the reference Wuhan-Hu-1 strain (NC_045512). C. Nucleotide and amino acid substitutions from the reference Wuhan-Hu-1 strain shared among the sequences from Lannion, France are represented as ticks along the SARS-CoV-2 genome and are annotated with text if non-synonymous. Light grey text annotated amino acid substitutions are not unique to the Lannion (B1.616) lineage.

### Population

From January 1^st^ to March 24^th^, 2021, 268 patients were hospitalized with SARS-CoV-2 infection in Lannion or Saint Brieuc hospitals, among which 86 lived in the B.1.616 epidemic area (Figure 2). CT-scan was typical of COVID-19 in 58/86 (67%) and serology was positive in 31/86 (36%). SARS-CoV-2 was detected by RT-PCR in 50/86 (58%) patients, including 10 with all B.1.616 genomic characteristic features, and 3 with viruses that could not be totally sequenced, but were classified as closely related to B.1.616 by WGS. These 13 patients were classified as confirmed B.1.616 COVID-19 cases. In addition, 21 patients developed COVID-19 not related to a VOC during or within 14 days of close contact with at least one patient with active COVID-19 classified as a confirmed B.1.616 case. Although no sample was available for WGS or PCR screening for these patients, they were classified as probable B.1.616 cases, according to our predefined criteria. Therefore, these 34 patients (13 confirmed and 21 probable) constituted the B.1.616 group.

**Figure 2:**
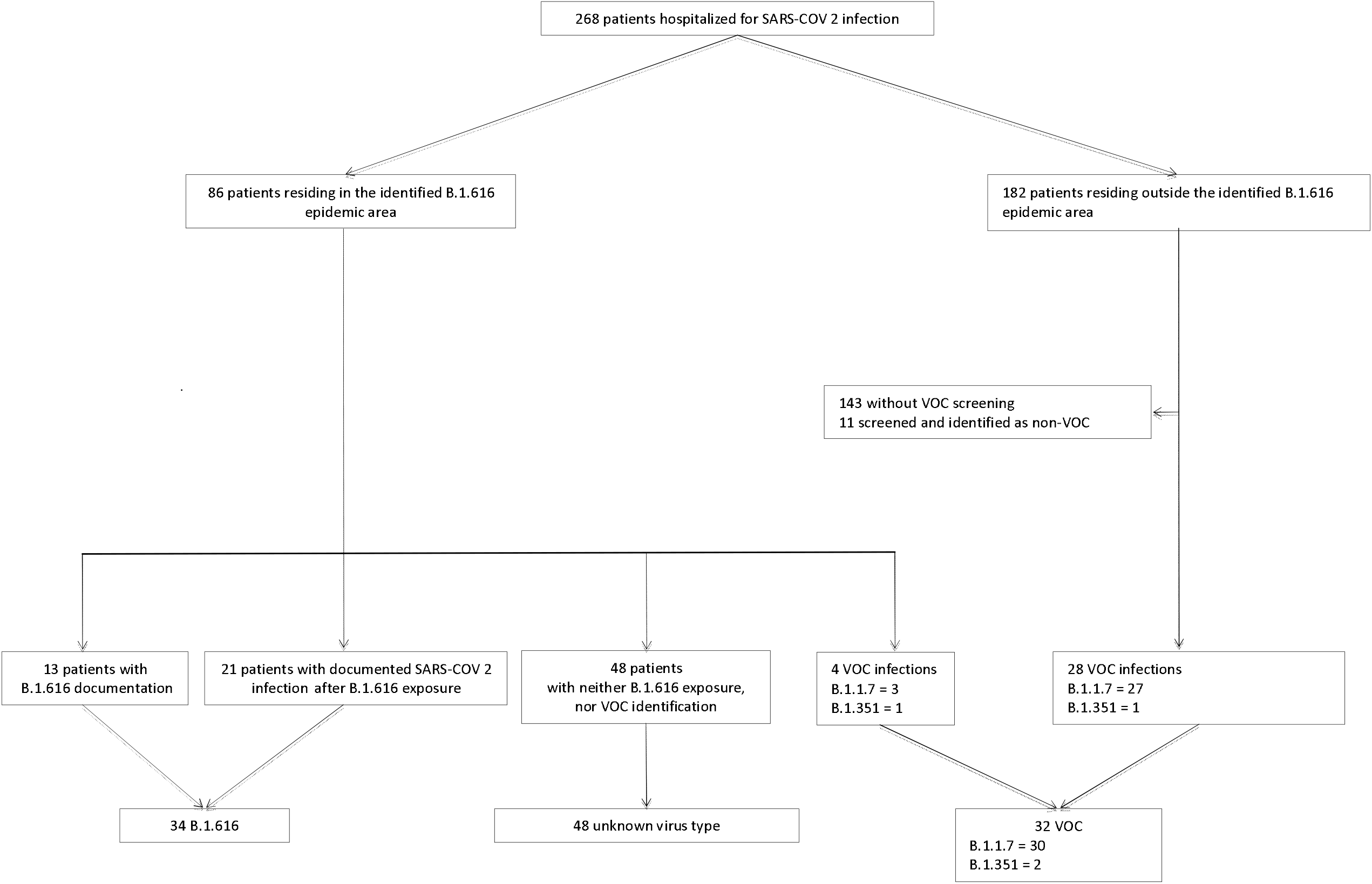
Flow chart Note. VOC, variant of concern

During the study period, 108 patients were hospitalized in a ward where at least one case of B.1.616 COVID-19 was admitted, for a total of 780 patients-days at risk. Of these 108 exposed patients, 32 (30%) developed COVID-19 symptoms after they were admitted for another reason and were categorized as nosocomial B.1.616 cases (11 confirmed B.1.616, and 21 probable). Therefore, the B.1.616 incidence rate in these wards was estimated at 41/1000 patient-days at risk. In addition, 52/86 patients living in the epidemic area were hospitalized because of COVID-19, in the absence of any contact with a B.1.616 case, among which 4 were infected by a VOC (B.1.1.7, n=3; B.1.351, n=1), and 48 were assigned to the unknown group.

Finally, among the 182 COVID-19 inpatients living outside of the B.1.616 epidemic area, 39 were screened for VOCs, among which 28 (72%) were infected with the B.1.1.7 (n=27) or the B.1.351 (n=1) variants. These 28 patients, combined with the 4 patients from the B.1.616 epidemic area infected with a VOC, were assigned to the VOC group.

Comorbidities and baseline characteristics are displayed on Table 2. Briefly, COVID-19 cases in the B.1.616 and the unknown groups were older than in the VOC group (respectively, 81 years [73-88] vs 80 years [68-86] vs 73 years [67-82], *P*=0.022). B.1.616 COVID-19 cases were less likely to be documented at first RT-PCR (15% vs 23% for unknown variant vs 97% for VOC, *P*<0.001), while time from first RT-PCR test to symptoms onset was not different (*P*=0.17), Table 3. Although B.1.616 cases had more RT-PCR tests (*P*<0.001), and more lower respiratory tract samples (*P*<0.001), only 21 (62%) had at least one positive RT-PCR. Among the 13 patients classified as B.1.616 COVID-19 in the absence of any positive RT-PCR, 12 had typical radiological findings on CT scan, and 5 had SARS-CoV-2 seroconversion, with a median time from symptoms onset to positive serology of 8 days [6-10] (supplementary Table 1). In addition, the median cycle threshold (Ct) in nasopharyngeal sample was higher in the B.1.616 and the unknown variant group, at 28 [26-33] and 35 [32-42], vs 19 [14-22] in the VOC group, *P*<0.001.

**Table 2:** Baseline characteristics

| Variables | B.1.616<br>n=34 | Undetermined<br>variant<br>n=48 | VOC<br>n=32 | p-value |
| --- | --- | --- | --- | --- |
| Age, year [IQR] | 81 [73 – 88] | 80 [68 – 86] | 73 [67 – 82] | 0.022 |
| Male, n (%) | 17 (50%) | 28 (58%) | 15 (47%) | 0.564 |
| Body mass index, kg/m<sup>2</sup> [IQR] | 25 [22 – 29] | 26 [23 – 28] | 31 [27-33] | 0.012 |
| *Comorbidities* |  |  |  |  |
| No comorbidity, n (%) | 6 (18%) | 9 (19%) | 4 (13%) | 0.812 |
| Cardiovascular disease, n (%) | 18 (53%) | 24 (50%) | 11 (34%) | 0.260 |
| Chronic respiratory disease, n (%) | 5 (15%) | 12 (25%) | 5 (16%) | 0.419 |
| Chronic kidney failure, n (%) | 9 (26%) | 10 (21%) | 5 (16%) | 0.557 |
| Cirrhosis, n (%) | 3 (9%) | 5 (10%) | 1 (3%) | 0.559 |
| Neurological disease, n (%) | 10 (29%) | 6 (12%) | 10 (31%) | 0.081 |
| Cancer, n (%) | 11 (32%) | 12 (25%) | 10 (31%) | 0.727 |
| Immunodepression, n (%) | 3 (9%) | 2 (4%) | 5 (16%) | 0.207 |
| Diabetes, n (%) | 6 (18%) | 14 (29%) | 6 (19%) | 0.384 |
| Hypertension, n (%) | 19 (56%) | 22 (46%) | 22 (69%) | 0.130 |
| Health-care associated COVID-19, n (%) | 32 (94%) | 20 (42%) | 10 (31%) | <0.001 |
| *Clinical findings* |  |  |  |  |
| Fever, n (%) | 24 (71%) | 27 (56%) | 21 (66%) | 0.392 |
| Temperature, °C | 38.3 [37.2 – 38.7] | 38 [37.4 – 38.5] | 37.8 [37.0-38.8] | 0.894 |
| Dyspnoea, n (%) | 23 (68%) | 30 (63%) | 14 (44%) | 0.113 |
| Oxygen saturation, % | 91 [90 – 94] | 93 [91 – 96] | 94 [89 - 96] | 0.098 |
| Cough, n (%) | 11 (32%) | 18 (38%) | 14 (44%) | 0.633 |
| Headache, n (%) | 2 (6%) | 3 (6%) | 1(3%) | 0.882 |
| Delirium, n (%) | 4 (12%) | 7 (15%) | 2(6%) | 0.569 |
| Fatigue, n (%) | 8 (24%) | 15 (31%) | 12 (38%) | 0.467 |
| Anosmia, n (%) | 1 (3.0%) | 2 (4%) | 0 | 0.783 |
| Digestive symptoms, n (%) | 4 (12%) | 5 (10%) | 1(3%) | 0.484 |
| Rhinorrhoea, n (%) | 0 | 4 (8%) | 4 (13%) | 0.105 |
| No symptoms – no. (%) | 0 | 4 (8%) | 4 (12%) | 0.105 |
| *Biological findings* |  |  |  |  |
| Neutrophils, x 10<sup>9</sup>/L | 4615 [3222 – 7550] | 4435 [3160 – 9298] | 4130 [1440-6700] | 0.150 |
| Lymphocytes, x 10<sup>9</sup>/L | 705 [430 – 1058] | 725 [518 – 1033] | 780 [610 - 1060] | 0.579 |
| Serum C-reactive protein, mg/mL | 81 [43 – 123] | 51 [24 – 135] | 49 [12 – 92] | 0.150 |
| D-dimeres, µg/L | 1330 [589 – 1714] | 722 [521 – 1609] | 1528 [930 – 1942] | 0.476 |
| *Thorax Computed Tomography (CT)* |  |  |  |  |
| No CT scan, n | 6 (18%) | 21 (44%) | 12 (37%) |  |
| <25%, n (%) * | 8(24%) | 10 (21%) | 5 (16%) | 0.953 |
| 25-50%, n (%) * | 12 (35%) | 11 (23%) | 9 (28%) |  |
| 50-75%, n (%) * | 7 (21%) | 6 (13%) | 5 (16%) |  |
| >75%, n (%) * | 1 (3%) | 0 | 1 (3%) |  |
| Pulmonary embolism, n (%) | 3 (12%) | 1 (2%) | 2 (6%) | 0.583 |

Note. VOC, variant of concern, \*Proportion of lung involved

**Table 3:** Virological and Radiological findings

| Variables | B.1.616<br>n=34 | Undetermined<br>variant<br>n=48 | VOC<br>n=32 | p-value |
| --- | --- | --- | --- | --- |
| Time from symptoms onset to first RT-PCR test, days [IQR] | 0 [0 – 1] | 0 [0 – 5] | 3 [1 – 5] | 0.170 |
| Time from symptoms onset to first SARS-CoV-2 detection, days [IQR] | 4 [2 – 10] | 4 [1 – 7] | 1 [0 – 4] | 0.047 |
| First RT-PCR positive for SARS-CoV-2, n (%) | 5 (15%) | 11 (23%) | 31 (97%) | <0.001 |
| At least one RT-PCR positive for SARS-CoV-2, n (%) | 21 (62%) | 25 (52%) | 32 (100%) | <0.001 |
| *Number of RT-PCR Test per patient* |  |  |  | <0.001 |
| 1 | 5 (12.9%) | 14 (27.3%) | 31 (97%) |  |
| 2 | 10 (33.3%) | 19 (38.2%) | 1 (3%) |  |
| >3 | 19 (54.5%) | 15 (34.5%) | 0 |  |
| *Site of sample (First positive RT-PCR only), n* |  |  |  | <0.001 |
| Endotracheal aspirate | 0 | 1 (2%) | 0 |  |
| Expectoration | 2 (6%) | 0 | 0 |  |
| Broncho-alveolar lavage | 6 (18%) | 0 | 0 |  |
| Nasopharyngeal swab | 13 (38%) | 23 (48%) | 32 (100%) |  |
| Stool | 0 | 1 (2%) | 0 |  |
| *CT value (Positive RT-PCR only)* |  |  |  |  |
| Nasopharyngeal sample | 28 [26 – 33] | 35 [32 – 42] | 19 [14–22] | <0.001 |
| Lower respiratory tract sample | 33 [33 – 35] | 45 [45 – 45] | 26 | 0.035 |

Note. VOC, variant of concern

As planned, we compared outcomes of B.1.616 and VOC cases, as patients in the unknown group were not considered for this analysis. Patients with B.1.616 COVID-19 had worse clinical outcomes: 22 (65%) reached a WHO score>5, 12 (35%) were admitted in the ICU, and 15 died (44%), Table 4. Variables independently associated with poor outcome in Cox proportional hazard regression were B.1.616 (HR 4.2 [1.3–13.5], *P*=0.018), and chronic respiratory diseases (HR 2.7 [1.1–6.4], *P*=0.029), Table 5. This independent association remained when the four asymptomatic patients from the VOC group were removed (HR 4.5 [1.2-16.8], *P*=0.024), and when the 13 WGS B.1.616 confirmed cases were compared with VOC cases (HR 13.7 [3.4-55.8], *P*<0.001). Moreover, case fatality rate was higher for B.1.616 COVID-19 cases than VOC cases (logrank test *P*=0.01), Figure 3, although this was not significant, after age adjustment (aHR 2.8 [0.96 – 7.9], *P*=0.057).

**Table 4:** Outcomes

| Variables | B.1.616<br>n=34 | Undetermined<br>variant<br>n=48 | VOC<br>n=32 | p-value |
| --- | --- | --- | --- | --- |
| Intensive care unit admission, n (%) | 12 (35%) | 7 (15%) | 10 (31%) | 0.071 |
| WHO score >5, n (%) | 22 (65%) | 19 (40%) | 12 (38%) | 0.039 |
| 28 day-mortality, n (%) | 15 (44%) | 13 (27%) | 5 (16%) | 0.036 |

Note. WHO, world health organization; VOC, variant of concern

**Table 5:** Risk factors for poor outcome (WHO>5) among the 34 B.1.616-related cases and the 32 VOC-related COVID-19 cases (cox proportional hazard regression)

| Variable | Univariable analysis |  |  | Multivariable analysis |  |  |
| --- | --- | --- | --- | --- | --- | --- |
|  | HR | IC | P | HR | IC | P |
| B.1.616<br>(vs VOC) | 2.6 | [1.3 – 5.6] | 0.011 | 4.2 | [1.3 – 13.5] | 0.018 |
| Age per supplementary 10 years | 1.0 | [0.8 – 1.2] | 0.699 | 0.9 | [0.7 – 1.2] | 0.438 |
| Male | 1.4 | [0.7 – 2.8] | 0.319 |  |  |  |
| Cardiovascular disease | 0.7 | [0.4 – 1.5] | 0.368 |  |  |  |
| Chronic respiratory disease | 2.2 | [1.0 – 5.1] | 0.065 | 2.7 | [1.1 – 6.4] | 0.029 |
| Chronic kidney failure | 1.0 | [0.4 – 2.3] | 0.960 |  |  |  |
| Cirrhosis | 1.3 | [0.3 – 4.4] | 0.941 |  |  |  |
| Neurological issue | 0.6 | [0.2 – 1.3] | 0.941 |  |  |  |
| Cancer | 0.9 | [0.4 – 1.9] | 0.697 |  |  |  |
| Immunodepression | 0.4 | [0.1 – 1.9] | 0.274 |  |  |  |
| Diabetes | 0.8 | [0.3 – 2.0] | 0.580 |  |  |  |
| Hypertension | 0.5 | [0.3 – 1.1] | 0.080 | 0.7 | [0.3 – 1.6] | 0.413 |
| Healthcare associated COVID | 1.3 | [0.6 – 2.8] | 0.452 | 0.7 | [0.2 – 2.3] | 0.585 |

Note. VOC, variant of concern

**Figure 3:**
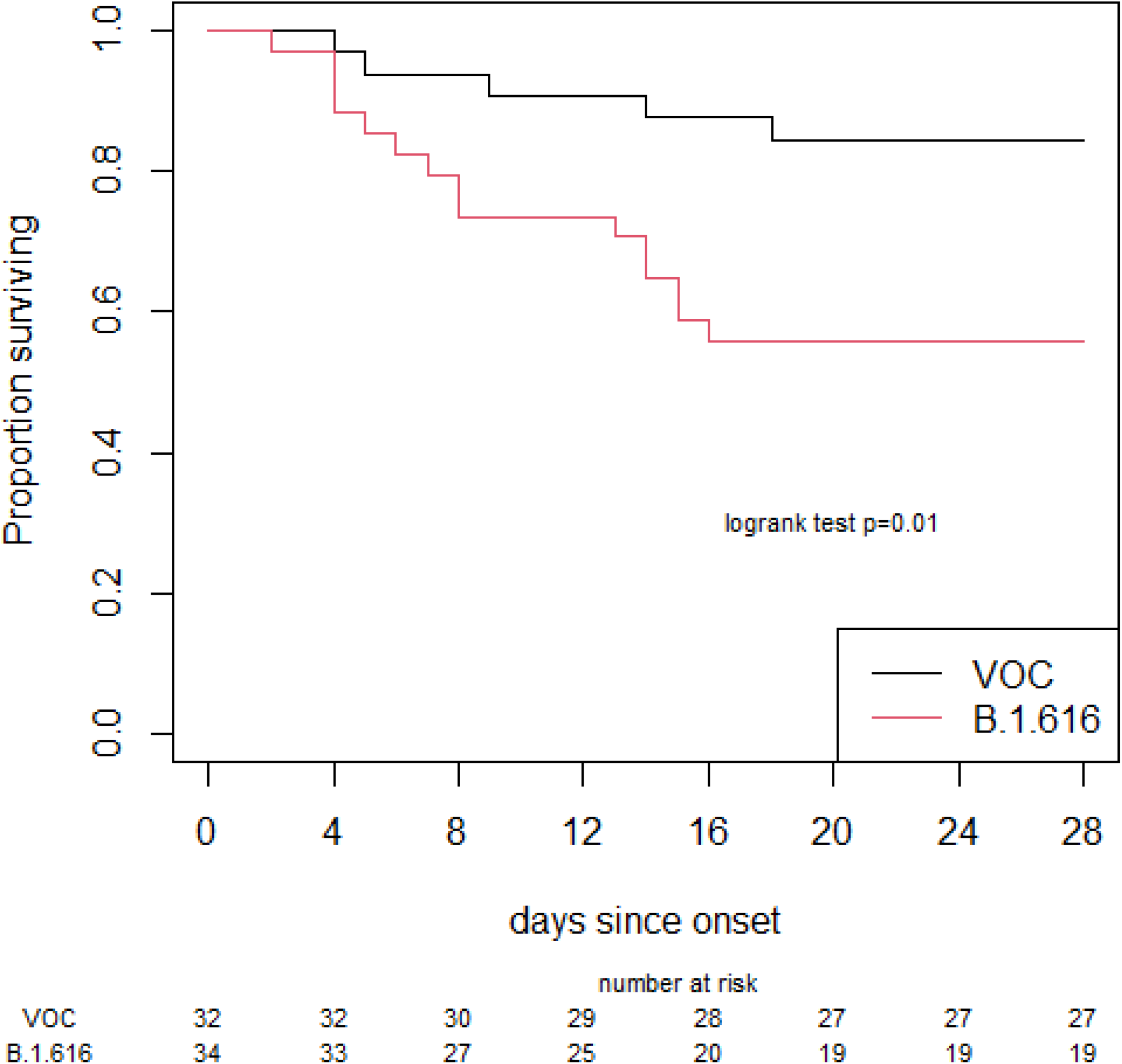
Survival curves Note. VOC, variant of concern

## DISCUSSION

The low rate of SARS-CoV-2 detection by RT-PCR tests on nasopharyngeal samples within a large cluster of COVID-19 cases led to the identification of a novel variant, B.1.616. Although COVID-19 cases due to this variant had clinical, biological and radiological findings in line with classical features of COVID-19, B.1.616 was associated with more severe disease. Recent clinical trial reported 39% of severe patients with high-flow oxygen or more invasive support at day 28, which is lower than in B.1.616 cases^19^. Similarly, B.1.616 was associated with higher 28-day mortality rate, at 15/34 (44%) vs. 5/32 (16%) for VOC, *P*=0.036, although this was not significant, after age adjustment (aHR 2.8 [0.96 – 7.9], *P*=0.057). COVID-19 in-hospital mortality ranged from 21% to 36% in previous studies, with higher mortality in nosocomial COVID-19, or in elderly patients ^20–22^. The high mortality rate among B.1.616 cases in our study must be interpreted with caution, as patients enrolled combined both pejorative factors, i.e., age (median, 81 years [73–88]), and primarily health care-acquisition (32/34, 94%). B.1.616 group included both WGS-confirmed B.1.616-related COVID-19 cases (confirmed B.1.616), and COVID-19 cases following close contact with a documented B.1.616 case (probable B.1.616), in the absence of WGS confirmation. This conservative bias implies that comparisons with controls may misjudge the actual differences, with the 21 probable cases being frails, therefore increasing morbidity. Other SARS-CoV-2 variants have been associated with an increase or a decrease in COVID-19 severity^4,23^. By the time of this writing, three variants are classified by the WHO as VOC, based on ‘evidence of an increase in transmissibility, more severe disease, significant reduction in neutralization by antibodies generated during previous infection or vaccination, reduced effectiveness of treatments or vaccines, or diagnostic detection failures’: B.1.1.7, B.1.351, and P.1 ^5^.

The second most salient features of COVID-19 cases related to B.1.616 was the high number of negative or weakly positive RT-PCR tests on nasopharyngeal samples. Previous studies estimated that 10-16% of patients with COVID-19 have negative RT-PCR tests on nasopharyngeal samples ^24,25^. A meta-analysis concluded that 1.8 to 33% of RT-PCR tests on first nasopharyngeal samples in COVID-19 patients are found negative ^26^. In our study, 16/21 (76%) of B.1.616 COVID-19 cases with at least one positive RT-PCR tested negative for RT-PCR on their first nasopharyngeal sample. Failure to detect B.1.616-related COVID-19 with the gold-standard diagnostic test most likely contributed to the emergence of several clusters, since implementation of specific infection control measures mostly relied on virological confirmation during the study period. Retrospective review of medical charts found that, in several cases, the diagnosis was suspected early in the COVID-19 course, but specific infection control measures were relieved once RT-PCR tests returned negative. Failure to detect B.1.616 on nasopharyngeal samples is even more problematic for the screening of contacts. Indeed, the incidence of nosocomial COVID-19 was much higher than reported ^7^ during the early phase of the outbreak, before we realized the low proportion of nasopharyngeal samples from which this specific circulating variant could be detected. Of note, although ‘diagnostic detection failures’ is one of the criteria to define VOC, none of the three major VOCs nor any of the current VUI meet this classification criteria. As RT-PCR assays used in France target at least two different viral genomic regions, it is unlikely that the B.1.616 strain would not be detected due to specific mismatches. Identification of this new lineage in lower respiratory tract samples confirmed the correct detection of the viral genome by commercially available assays used during the study period. Repeated failures to detect B.1.616 on nasopharyngeal samples were therefore most likely due to SARS-CoV-2 viral loads below detection threshold in this site, rather than sub-optimal sensitivity of routine RT-PCR tests.

Kim *et al*. provided original data on time from illness onset to viral clearance combining SARS-CoV-2 cultures along with RT-PCR tests ^27^. They found that viral culture was positive only in samples with a CT value of 28 or less. In our study, 75% of patients with positive nasopharyngeal RT-PCR tests in the B.1.616 group had a CT value higher than 26. This suggests that in most COVID-19 patients infected with B.1.616, viral cultures from nasopharyngeal samples would be negative, which may indicate a lower risk of secondary transmission. This assumption might explain why the VOC B.1.1.7 remains largely predominant over B.1.616 in our area, as B.1.1.7 transmission is estimated as more efficient than other SARS-CoV-2 lineages ^28^. Likewise, the duration of nasopharyngeal virus shedding might be reduced with B.1.616 as compared with other SARS-CoV-2 variants. Of note, RT-PCR was positive on nasopharyngeal samples for only 32% of patients with pneumonia due to SARS-CoV ^29^, and the Middle East respiratory syndrome-related coronavirus (MERS-CoV) is mostly detected in lower respiratory tract specimens, with high viral load, whereas nasopharyngeal specimens are poorly contributive ^30^. The mechanisms leading to unreliable detection of the B.1.616 variant, likely because of low viral load in the upper respiratory tract soon after infection, and when symptoms occur, will need to be further explored. Whether a change of tropism could explain the detection of the virus in the lower respiratory tract upon worsening of the symptoms remains to be determined. Indeed, detection of SARS-CoV-2 in the lower respiratory tract at a time when the virus is no longer detected in the upper respiratory tract has been documented in severe patients. The functional impact of the mutations noted in the SARS-CoV-2 variant identified here will require further investigation. In particular, it will be of interest to determine to what extent the changes in the two proteins involved in antagonism of the host antiviral response could result in altered innate immune responses and the unusual course of infection.

In our study, among patients with a positive RT-PCR assay in the B.1.616 group, the sensitivity of one, two, three, and four tests on nasopharyngeal samples were, respectively, 5/34 (15%), 13/34 (38%), 14/34 (45%), and 17/34 (55%). RT-PCR tests on sputum, or broncho-alveolar lavage (BAL), were positive in 8/34 (24%) B.1.616-related COVID-19 cases with previous negative nasopharyngeal RT-PCR tests. As samples from the lower respiratory tract are more difficult to obtain in frail patients, the real extent of the B.1.616-related COVID-19 outbreak in our institution has probably been underestimated. A large surveillance study, with sequencing of a representative sample of 15% of all RT-PCR-positive COVID-19 cases during the study period found no community-acquired B.1.616-related COVID-19 (Flash study#5, SpF, Paris, France, unpublished data), but the low detection in standard sampling may have contributed to this result.

Our study has limitations. First, the small sample size and the retrospective design both limit the statistical power. However, due to the fast pace of the pandemic, early communication on the characteristics of new variants is warranted, and this would be the first case series of B.1.616-related COVID-19 cases. Second, B.1.616 confirmed cases where those in whom a deep respiratory sample was obtained, mostly motivated by disease severity, hence constituting a selection bias.. Finally, the selection of controls may be an additional limitation: VOC-related cases were selected as controls mostly because these cases could be reliably classified as ‘non-B.1.616’: indeed, other COVID-19 cases managed during the study period were no longer available for virological characterization, hence we could not rule out that they could be B.1.616. Of note, our VOC control group was mostly constituted by the B.1.1.7 variant, which is largely dominant in Western Europe, and has been associated with an increased mortality^4^. Hence, as B.1.616 cases were independently associated with worse outcomes as compared to VOC, this would probably be even more pronounced if compared with COVID-19 cases non-related to VOC. In addition, all consecutive cases with available VOC screening were enrolled in the control group, which limits selection bias.

## CONCLUSION

We report a nosocomial outbreak of COVID-19 cases related to a new variant, B.1.616, characterized by poor detection by RT-PCR tests on nasopharyngeal samples despite typical clinical, radiological, and biological features of COVID-19. We also noted high case fatality rate in our sampled population. The novel variant reported here adds to the diversity of emerging SARS-CoV-2 variants with impact on early diagnosis and control. Further investigations are required to confirm those observations. This work also highlights the difficulties to manage nosocomial cases when the gold-standard test fails to confirm the diagnosis. With constantly emerging new variants, one should remain attentive to any unusual clinical situation that could be linked to such emergence.

## Data Availability

A complete de-identified patient dataset, accompanied by the original study protocol will be available to European researchers on request. Individuals wishing to access the data should send a request to.

## DECLARATIONS

### Ethics approval

This study respects the French reference method MR004.

### Contributors

PF, NM conceptualization, formal analysis, methodology, writing original draft. MJ-D, VT, CG, CP, EG, SB, virological assays, investigation, writing - review & editing. AB and AZ: phylogenetic analysis. ESL supervision, phylogenetic analysis, writing – review & editing. VE, sequencing. XL conceptualization, methodology. BG, RV, MT writing - review & editing. DC conceptualization, methodology, supervision, formal analysis, writing - review & editing. SW conceptualization, supervision, formal analysis, writing - review & editing, PT conceptualization, methodology, supervision, writing - review & editing. PR, AG, CM, SG, ED, VM, RB, MV, EG were investigators. All authors revised the final version.

## Acknowledgments

We thank Olivia DA CONCEICAO and Cylia IMEKHLAF for their help in collecting data from patients charts.

We would like to thank all of the healthcare workers, public health employees, and scientists involved in the COVID-19 response and this outbreak.

We acknowledge the authors, originating and submitting laboratories of the sequences from GISAID (Table S2). We avoided any direct analysis of genomic data not submitted as part of this paper and used this genomic data only as background. This work used the computational and storage services (Maestro cluster) provided by the IT department at Institut Pasteur, Paris.

---

**Supplementary Table 1:** Seroconversion in B 1.616 patients with negative RT-PCR.

| Case # | 1st serology<br>(DPS<sup>§</sup>) | IgG* | IgM* | 2nd serology<br>(DPS) | IgG* | IgM* |
| --- | --- | --- | --- | --- | --- | --- |
| 1 | 0 | 1.05 | 0.53 | 7 | 6.77 | 7.24 |
| 2 | 0 | 0.02 | 0.03 | 10 | 3.66 | 17.06 |
| 3 | 7 | 1.81 | 2.38 | 11 | 6.63 | 10.08 |
| 4 | 2 | 0.89 | 0.28 | 8 | 6.2 | 1.92 |
| 5 | 0 | 0.01 | 0.27 | 12 | 1.94 | 11.75 |

<sup>§</sup>DPS: day post-symptoms onset; \*IgG and IgM levels as determined with the commercial assay AdviseDx SARS-CoV-2
Alinity Abbott

**Supplementary table 2 (file supplementary material)**

