## Supplementary table 2 for "A new SARS-CoV-2 variant poorly detected by RT-PCR on nasopharyngeal samples, with high lethality"

| GISAID_accession | originating_lab | submitting_lab | authors |
|---|---|---|---|
| EPI_ISL_1167055,EPI_ISL_1167058,EPI_ISL_1167075,EPI_ISL_1167086 | Dr. Andrija Stampar" Teaching Institute of Public Health, Department of Clinical Microbiology" | Istituto di Genomica Applicata | Jasmina Vranes et al |
| EPI_ISL_993056 | Dr. Andrija Stampar“ Teaching Institute of Public Health, Department of Clinical Microbiology" | Institute of Applied Genomics | Jasmina Vranes et al |
| EPI_ISL_468145,EPI_ISL_468149,EPI_ISL_468153,EPI_ISL_468156 | [Romania, Bucharest] National Institute for Infectious Diseases “Prof. Dr. Matei Balș” | [Romania, Bucharest] National Institute for Infectious Diseases “Prof. Dr. Matei Balș” | Leontina Banica et al |
| EPI_ISL_699655,EPI_ISL_707697,EPI_ISL_707791,EPI_ISL_733500,EPI_ISL_763065,EPI_ISL_763067,EPI_ISL_794735 | 1-Laboratory of Microbiology, National Reference Lab, Charles Nicolle Hospital; 2-University of Tunis ElManar, Faculty of Medicine of Tunis, LR99ES09, Tunis, Tunisia | 1-Clinical and Experimental Pharmacology Lab, LR16SP02, National Center of Pharmacovigilance, University of Tunis El Manar, Tunis, Tunisia. 2-Neurodegenerative diseases and psychiatric troubles, LR18SP03, Razi Hospital, University of Tunis El Manar, Tunis, Tunisia. 3- Ministry of Health, National Observatory of New and Emerging Diseases, 1006, Tunis, Tunisia | Ilhem Boutiba-Ben Boubaker et al |
| EPI_ISL_455442 | 1. ViroGenetics - BSL3 Laboratory of Virology, Małopolska Centre of Biotechnology, Jagiellonian University; 2. II Department of Internal Medicine, Faculty of Medicine, Jagiellonian University Medical College; 3. Narodowy Instytut Zdrowia Publicznego – Państwowy Zakład Higieny (NIZP-PZH) | 1. ViroGenetics - BSL3 Laboratory of Virology, Małopolska Centre of Biotechnology, Jagiellonian University; 2. II Department of Internal Medicine, Faculty of Medicine, Jagiellonian University Medical College; 3. Narodowy Instytut Zdrowia Publicznego – Państwowy Zakład Higieny (NIZP-PZH). | Katarzyna Pancer et al |
| EPI_ISL_882703,EPI_ISL_954190 | 1.AO Universitaria 'S. Giovanni di Dio e Ruggi D'Aragona, Scuola Medica Salernitana' Hospital / 2.UOC di Virologia e Microbiologia, Università della Campania 'L. Vanvitelli' / 3.AO Universitaria 'Federico II' Napoli Hospital / 4.AORN 'San Giuseppe Moscati' Avellino Hospital / 5.AO 'San Pio - presidio G. Rummo' Benevento Hospital / 6.AO 'Sant'Anna e San Sebastiano' Caserta Hospital / 7.PO 'Maria Santissima Addolorata' Eboli Hospital / 8.Biogem Istituto di Ricerche Genetiche | 1. Genome Research Center for Health (CRGS) / 2. Laboratory of Molecular Medicine and Genomics(LMMGe) / 3. Center for Research in Pure and Applied Mathematics (CRMPA) | Giorgio Giurato et al |
| EPI_ISL_640056 | 2 Military Hospital wc MAA | NHLS/UCT | Arash Iranzadeh et al |
| EPI_ISL_517616,EPI_ISL_517634,EPI_ISL_517635,EPI_ISL_517642,EPI_ISL_517643,EPI_ISL_517657,EPI_ISL_517661,EPI_ISL_517662,EPI_ISL_518811,EPI_ISL_518812 | Academic Hospital Paramaribo | Erasmus Medical Center | Bas Oude Munnink et al |
| EPI_ISL_500793 | Akershus University Hospital, Department for Microbiology and Infectious Disease Control | Norwegian Institute of Public Health, Department of Virology | Kathrine Stene-Johansen et al |
| EPI_ISL_747240 | Al Islam Hospital | West Java Health Laboratory; School of Life Sciences and Technology, Institut Teknologi Bandung | Azzania Fibriani et al |
| EPI_ISL_739660 | Al-Quds Nutrition and Health Research Institute, Al-Quds University | Al-Quds Nutrition and Health Research Institute, Al-Quds University | Nasereddin et al |
| EPI_ISL_672095 | Alameda County Public Health Lab | Chan-Zuckerberg Biohub | CZB Cliahub Consortium et al |
| EPI_ISL_1287290 | Alexianer DaKS GmbH (eigenes IT-Tochterunternehmen der Alexianer Krankenhäuser) | Robert Koch Institute | et al |
| EPI_ISL_528702 | Alsafar - Khalifa University Abu Dhabi | Alsafar - Khalifa University Abu Dhabi | Andreas Henschel et al |
| EPI_ISL_458150 | ANOUAL | ANOUAL | Jouali Farah et al |
| EPI_ISL_909955,EPI_ISL_909960 | Apollo Hospitals | CSIR-Centre for Cellular and Molecular Biology | Onkar Kulkarni et al |
| EPI_ISL_682235 | AREA DE SALUD ALAJUELA NORTE - CLINICA DR. MARCIAL RODRIGUEZ | Inciensa, Instituto Costarricense de Investigación y Enseñanza en Nutrición y Salud | Francisco Duarte et al |
| EPI_ISL_770030 | Area De Salud Buenos Aires | Inciensa, Instituto Costarricense de Investigación y Enseñanza en Nutrición y Salud | Francisco Duarte et al |
| EPI_ISL_914794,EPI_ISL_1067588 | AREA DE SALUD BUENOS AIRES | Inciensa, Instituto Costarricense de Investigación y Enseñanza en Nutrición y Salud | Francisco Duarte et al |
| EPI_ISL_527751,EPI_ISL_770008 | Area De Salud Corredores | Inciensa, Instituto Costarricense de Investigación y Enseñanza en Nutrición y Salud | Francisco Duarte et al |
| EPI_ISL_1196424 | AREA DE SALUD DESAMPARADOS 1 - CLINICA DR. MARCIAL FALLAS | Inciensa, Instituto Costarricense de Investigación y Enseñanza en Nutrición y Salud | Francisco Duarte et al |
| EPI_ISL_491437 | Area de Salud Escazu (Coopesana) | Inciensa, Instituto Costarricense de Investigación y Enseñanza en Nutrición y Salud | Francisco Duarte et al |

| | | | |
|---|---|---|---|
| EPI_ISL_1067605 | AREA DE SALUD HATILLO - CLINICA DR. SOLON NUÑEZ | Inciensa, Instituto Costarricense de Investigación y Enseñanza en Nutrición y Salud | Francisco Duarte et al |
| EPI_ISL_512666,EPI_ISL_512667,EPI_ISL_770000 | Area De Salud La Cruz | Inciensa, Instituto Costarricense de Investigación y Enseñanza en Nutrición y Salud | Francisco Duarte et al |
| EPI_ISL_512660 | Area De Salud Los Chiles | Inciensa, Instituto Costarricense de Investigación y Enseñanza en Nutrición y Salud | Francisco Duarte et al |
| EPI_ISL_770005 | Area De Salud Moravia | Inciensa, Instituto Costarricense de Investigación y Enseñanza en Nutrición y Salud | Francisco Duarte et al |
| EPI_ISL_1201439 | AREA DE SALUD PAVAS (COOPESALUD) | Inciensa, Instituto Costarricense de Investigación y Enseñanza en Nutrición y Salud | Francisco Duarte et al |
| EPI_ISL_770017 | Area De Salud Perez Zeledon | Inciensa, Instituto Costarricense de Investigación y Enseñanza en Nutrición y Salud | Francisco Duarte et al |
| EPI_ISL_769989,EPI_ISL_770023 | Area De Salud San Francisco-San Antonio (Coopesana) | Inciensa, Instituto Costarricense de Investigación y Enseñanza en Nutrición y Salud | Francisco Duarte et al |
| EPI_ISL_770007 | Area De Salud San Rafael | Inciensa, Instituto Costarricense de Investigación y Enseñanza en Nutrición y Salud | Francisco Duarte et al |
| EPI_ISL_509505 | Area of Virology, Serology and Virology Division (SAViD), New South Wales Health Pathology Randwick | Area of Virology, Serology and Virology Division (SAViD), New South Wales Health Pathology Randwick | Rawlinson et al |
| EPI_ISL_678350,EPI_ISL_717711 | Area of Virology, Serology and Virology Division (SAViD), New South Wales Health Pathology Randwick | Virology Research Laboratory; Area of Virology, Serology and Virology Division (SAViD), New South Wales Health Pathology Randwick | Foster et al |
| EPI_ISL_542237 | ASST GOM Niguarda | Dep. Of Oncology and Hemato-Oncology University of Milan | Claudia Alteri et al |
| EPI_ISL_413490 | Auckland Hospital | Institute of Environmental Science and Research (ESR) | Matt Storey et al |
| EPI_ISL_583573 | Austrian Agency for Health and Food Safety (AGES) | Bergthaler laboratory, CeMM Research Center for Molecular Medicine of the Austrian Academy of Sciences | Alexandra Popa et al |
| EPI_ISL_853971,EPI_ISL_853977 | Austrian Agency for Health and Food Safety (AGES) | Bergthaler laboratory, CeMM Research Center for Molecular Medicine of the Austrian Academy of Sciences | Lukas Endler et al |
| EPI_ISL_1081798,EPI_ISL_1081802,EPI_ISL_1081804,EPI_ISL_1081805,EPI_ISL_1081807,EPI_ISL_1081808 | Baguio General Hospital and Medical Center | Philippine Genome Center | Francis A. Tablizo et al |
| EPI_ISL_1159375 | Bahman Hospital | Laboratory of Molecular Biology and Cancer Immunology,Lebanese University Public Health England | Fadi Abdel Sater et al |
| EPI_ISL_791978 | Balai Labkes Lampung | National Institute of Health Research and Development | Nugraha et al |
| EPI_ISL_434693 | Bamrasnaradura hospital | National Institute of Health. Department of medical Sciences, Ministry of Public Health, Thailand | Pilailuk et al |
| EPI_ISL_403962,EPI_ISL_403963 | Bamrasnaradura Hospital | 1. Department of Medical Sciences, Ministry of Public Health, Thailand 2. Thai Red Cross Emerging Infectious Diseases - Health Science Centre 3. Department of Disease Control, Ministry of Public Health, Thailand | Pilailuk et al |
| EPI_ISL_860186 | Bangalore Medical College and Research Institute | Department of Neurovirology, National Institute of Mental Health and Neurosciences (NIMHANS) | Chitra Pattabiraman et al |
| EPI_ISL_995709,EPI_ISL_995710,EPI_ISL_995724,EPI_ISL_995763 | BANGALORE MEDICAL COLLEGE AND RESEARCH INSTITUTE | Department of Neurovirology, National Institute of Mental Health and Neurosciences (NIMHANS) | Chitra Pattabiraman et al |
| EPI_ISL_843128 | Barts Health NHS Trust | COVID-19 Genomics UK (COG-UK) Consortium | CUTINO-MOGUEL et al |
| EPI_ISL_833333,EPI_ISL_833334 | Batangas City Health Office | Research Institute for Tropical Medicine | Hannah Leah Morito et al |
| EPI_ISL_1144010 | Bayerisches Landesamt für Gesundheit und Lebensmittelsicherheit (LGL) | Robert Koch Institute | et al |
| EPI_ISL_747244,EPI_ISL_803912,EPI_ISL_1250853,EPI_ISL_1250855,EPI_ISL_1250856,EPI_ISL_1250858 | BBMP Urban PHC | Department of Neurovirology, National Institute of Mental Health and Neurosciences (NIMHANS) | Chitra Pattabiraman et al |
| EPI_ISL_415580,EPI_ISL_415577,EPI_ISL_415578,EPI_ISL_412965 | BCCDC Public Health Laboratory | BCCDC Public Health Laboratory | Harrigan et al |
| EPI_ISL_973567,EPI_ISL_974478,EPI_ISL_974493,EPI_ISL_974721,EPI_ISL_975486,EPI_ISL_976529 | BCCDC Public Health Laboratory | BCCDC Public Health Laboratory | Prystajecky Natalie et al |
| EPI_ISL_1015370 | BCRM Cherbourg | IRBA, 2MI | GORGE O. et al |

| | | | |
|---|---|---|---|
| EPI_ISL_1117376,EPI_ISL_1117378,EPI_ISL_1168770 | Beijing Center for Disease Prevention and Control | Beijing Center for Disease Prevention and Control | Yang Pan et al |
| EPI_ISL_509711,EPI_ISL_509712,EPI_ISL_509713,EPI_ISL_509714 | Belize Ministry of Health | Pathogen Discovery, Respiratory Viruses Branch, Division of Viral Diseases, Centers for Disease Control and Prevention | Jing Zhang et al |
| EPI_ISL_780384,EPI_ISL_780385,EPI_ISL_780386,EPI_ISL_780387,EPI_ISL_780390,EPI_ISL_780392,EPI_ISL_780393,EPI_ISL_780395,EPI_ISL_780396,EPI_ISL_780404 | Bermuda Government Molecular Diagnostics Laboratory (MDL) | Respiratory Virus Unit, National Infection Service, Public Health England | PHE Covid Sequencing Team et al |
| EPI_ISL_430843 | Bethany Hospital | Research Institute for Tropical Medicine | Medado et al |
| EPI_ISL_1273093,EPI_ISL_1273088,EPI_ISL_1273089 | Biochemistry and Molecular Biology Department-Faculty of Medicine, Al-Quds University | Biochemistry and Molecular Biology Department-Faculty of Medicine, Al-Quds University | Ereqat et al |
| EPI_ISL_429999,EPI_ISL_430003,EPI_ISL_430013,EPI_ISL_430012,EPI_ISL_450187,EPI_ISL_450188,EPI_ISL_450189,EPI_ISL_635777,EPI_ISL_635778,EPI_ISL_635779,EPI_ISL_635780,EPI_ISL_635781,EPI_ISL_635782,EPI_ISL_730263,EPI_ISL_730461,EPI_ISL_730479,EPI_ISL_730523,EPI_ISL_730530,EPI_ISL_730534,EPI_ISL_730543,EPI_ISL_730545,EPI_ISL_730546,EPI_ISL_755127,EPI_ISL_755129,EPI_ISL_755145,EPI_ISL_755267,EPI_ISL_878516,EPI_ISL_878571,EPI_ISL_878574,EPI_ISL_878576 | Biolab Diagnostic Laboratories | Andersen lab at Scripps Research | Issa Abu-Dayyeh et al |
| EPI_ISL_526988,EPI_ISL_526985,EPI_ISL_526990,EPI_ISL_527002 | Biological prevention, army | Biological prevention, army | Seadawy et al |
| EPI_ISL_907080 | Biology, MCL | Biology, MCL | Seadawy et al |
| EPI_ISL_985084,EPI_ISL_985091,EPI_ISL_985096,EPI_ISL_985098 | Biorepository and Clinical Virology Laboratory | Ozer Lab | Ramon Lorenzo-Redondo et al |
| EPI_ISL_1145181 | Bioscientia Labor Wermsdorf | Robert Koch Institute | et al |
| EPI_ISL_747238 | Bogor Public Health | West Java Health Laboratory; School of Life Sciences and Technology, Institut Teknologi Bandung | Azzania Fibriani et al |
| EPI_ISL_871872,EPI_ISL_935040,EPI_ISL_935042,EPI_ISL_935045,EPI_ISL_933724,EPI_ISL_962886,EPI_ISL_965277,EPI_ISL_1287349,EPI_ISL_1296316,EPI_ISL_1287760 | Botswana Harvard HIV Reference Laboratory | Botswana Harvard HIV Reference Laboratory | Sikhulile Moyo et al |
| EPI_ISL_859563,EPI_ISL_859601,EPI_ISL_859622,EPI_ISL_859629,EPI_ISL_859657,EPI_ISL_859769,EPI_ISL_859792,EPI_ISL_859819,EPI_ISL_859852,EPI_ISL_859878,EPI_ISL_859996,EPI_ISL_860021,EPI_ISL_860028,EPI_ISL_860067,EPI_ISL_860084 | BTC, Khalifa University | BTC, Khalifa University | Al Safar et al et al |
| EPI_ISL_1001001,EPI_ISL_1001002,EPI_ISL_1001003,EPI_ISL_1034755,EPI_ISL_1034756,EPI_ISL_1034757,EPI_ISL_1034758,EPI_ISL_1034759,EPI_ISL_1046791,EPI_ISL_1046792,EPI_ISL_1046793 | Bundeswehr Institute of Microbiology | Bundeswehr Institute of Microbiology | Markus Antwerpen et al |

| | | | |
|---|---|---|---|
| EPI_ISL_833336 | Bureau of Quarantine | Research Institute for Tropical Medicine | Hannah Leah Morito et al |
| EPI_ISL_1135056 | C.H. PRINCESSE GRACE | CNR Virus des Infections Respiratoires - France SUD | Antonin Bal et al |
| EPI_ISL_935793 | Cadham Provincial laboratory | National Microbiology Laboratory (NML) | Anna Majer et al |
| EPI_ISL_755016 | California Department of Public Health | California Department of Public Health | CDPH IDLB COVIDNet et al |
| EPI_ISL_410044 | California Department of Public Health | Pathogen Discovery, Respiratory Viruses Branch, Division of Viral Diseases, Centers for Disease Control and Prevention | Jing Zhang et al |
| EPI_ISL_1096139,EPI_ISL_1098603,EPI_ISL_1098604,EPI_ISL_1098605 | Cambodian National Public Health Laboratory, National Institute of Public Health | Virology Unit, Institut Pasteur du Cambodge | Sokhoun Yann et al |
| EPI_ISL_576371,EPI_ISL_857327,EPI_ISL_857326,EPI_ISL_857323,EPI_ISL_857335,EPI_ISL_890221,EPI_ISL_890205,EPI_ISL_890215,EPI_ISL_907098 | Cancer Biology Department, National Cancer Institute | Cancer Biology Department, National Cancer Institute | Zekri et al |
| EPI_ISL_548138,EPI_ISL_548139,EPI_ISL_579479,EPI_ISL_579490,EPI_ISL_579496,EPI_ISL_622789,EPI_ISL_622791,EPI_ISL_755619 | Canterbury Health Laboratories | Institute of Environmental Science and Research (ESR) | Xiaoyun Ren et al |
| EPI_ISL_977539,EPI_ISL_977538,EPI_ISL_977584,EPI_ISL_977594,EPI_ISL_977583,EPI_ISL_977581 | Caribbean Public Health Agency | Carrington Lab, Department of PreClinical Sciences | Nikita S. D. Sahadeo et al |
| EPI_ISL_977659,EPI_ISL_977658,EPI_ISL_977656,EPI_ISL_977655 | Caribbean Public Health Agency | Carrington Lab, Department of PreClinical Sciences, Building 36, First Floor Biochemistry Unit, Faculty of Medical Sciences, The University of the West Indies | Nikita S. D. Sahadeo et al |
| EPI_ISL_872196,EPI_ISL_872194 | Caribbean Public Health Agency | Carrington Lab, Department of PreClinical Sciences, Faculty of Medical Sciences, The University of the West Indies | Nikita S. D. Sahadeo et al |
| EPI_ISL_872193 | Caribbean Public Health Agency | Carrington Lab, Department ofBuilding 36, First Floor Biochemistry Unit, Faculty of Medical Sciences, The University of the West Indies | Nikita S. D. Sahadeo et al |
| EPI_ISL_1060885,EPI_ISL_1060901,EPI_ISL_1060958 | CDL Laboratorio Santos e Vidal LTDA. | Instituto de Medicina Tropical de Sao Paulo | Brazil-UK Centre for Arbovirus Discovery Diagnosis Genomics et al |
| EPI_ISL_1220348 | CDP PIERRE OUDOT | CNR Virus des Infections Respiratoires - France SUD | Antonin Bal et al |
| EPI_ISL_1122433,EPI_ISL_1122443,EPI_ISL_1122446,EPI_ISL_1122452,EPI_ISL_1213539,EPI_ISL_1213552,EPI_ISL_1213556 | Cebu TB Reference Laboratory | Philippine Genome Center | Francis A. Tablizo et al |
| EPI_ISL_971451 | Cell culture Unit at CV-MIT belonging to HIMMV | Functional Genomic Platform UATRS-biology, CNRST | Nadia Touil et al |
| EPI_ISL_459864 | Center for Genome Regulation (CRG) | Center for Mathematical Modeling and Center for Genome Regulation. Santiago, Chile | Gaete A et al |
| EPI_ISL_522446,EPI_ISL_522448,EPI_ISL_522451,EPI_ISL_522457,EPI_ISL_522459,EPI_ISL_522463,EPI_ISL_522467 | Center for Laboratory Control of Infectious Diseases, Korea Centers for Diseases Control and Prevention | Center for Laboratory Control of Infectious Diseases, Korea Centers for Diseases Control and Prevention | Junyoung Kim et al |
| EPI_ISL_493137 | Center for Research and Innovation, Faculty of Medical Technology, Mahidol University | Center for Research and Innovation, Faculty of Medical Technology, Mahidol University | Kantima Sangsiriwut et al |
| EPI_ISL_913082 | Center for Virology | Center for Virology | Jeremy V. Camp et al |
| EPI_ISL_475807,EPI_ISL_583708 | Center for Virology, Medical University of Vienna | Bergthaler laboratory, CeMM Research Center for Molecular Medicine of the Austrian Academy of Sciences | Alexandra Popa et al |
| EPI_ISL_853891 | Center for Virology, Medical University of Vienna | Bergthaler laboratory, CeMM Research Center for Molecular Medicine of the Austrian Academy of Sciences | Lukas Endler et al |
| EPI_ISL_1160167 | Center of Medical Microbiology, Virology, and Hospital Hygiene, University of Duesseldorf | Center of Medical Microbiology, Virology, and Hospital Hygiene, University of Duesseldorf | Maximilian Damagnez et al |
| EPI_ISL_428488 | Centers for Disease Control, R.O.C. (Taiwan) | Centers for Disease Control, R.O.C. (Taiwan) | Ji-Rong Yang et al |
| EPI_ISL_815257,EPI_ISL_815390,EPI_ISL_815398,EPI_ISL_815255 | Centogene | Centogene | Peter Bauer et al |

| | | | |
|---|---|---|---|
| EPI_ISL_794602 | Central Laboratories, Egyptian Ministry of Health and Population | Central Laboratories, Egyptian Ministry of Health and Population | Kayed et al |
| EPI_ISL_794599,EPI_ISL_794593 | Central Laboratories, Egyptian Ministry of Health and Population | Central Laboratories, Egyptian Ministry of Health and Population | Roshdy et al |
| EPI_ISL_943587 | Central Laboratory of Public Health of Rio Grande do Sul (Lacen-RS) | State Center for Health Surveillance of the Health Department of the State of Rio Grande do Sul (CEVS/SES-RS) | Aline Campos et al |
| EPI_ISL_794606,EPI_ISL_794608 | Central Public Health Laboratories, Egyptian Ministry of Health and Population | Central Public Health Laboratories, Egyptian Ministry of Health and Population | El-Shesheny et al |
| EPI_ISL_885143 | Central public health laboratory | Molecular Diagnostics Department, Central public health laboratory | Dler et al |
| EPI_ISL_693471,EPI_ISL_693472,EPI_ISL_693474,EPI_ISL_693475,EPI_ISL_693476,EPI_ISL_693478,EPI_ISL_693479,EPI_ISL_693480,EPI_ISL_693481,EPI_ISL_693482,EPI_ISL_693483 | Central Public Health Laboratory | National Public Health Laboratory, National Centre for Infectious Diseases | Tze Minn Mak et al |
| EPI_ISL_978512,EPI_ISL_1068341 | Central Public Health Laboratory - LACEN - Bahia, Salvador, Brazil | Central Public Health Laboratory - LACEN -Bahia, Salvador, Brazil | Stephane Tosta et al |
| EPI_ISL_419211 | Central Virology Laboratory | Israel Institute for Biological Research | Inbar Cohen-Gihon et al |
| EPI_ISL_486437,EPI_ISL_486411,EPI_ISL_501896,EPI_ISL_534201 | Centrālā laboratorija | Latvian Biomedical Research and Study Centre | Ivars Silamiķelis et al |
| EPI_ISL_788934,EPI_ISL_788935 | Centre de Recherche et de Formation en Infectiologie Guinée | TransVIHMI, IRD/INSERM/Monpellier University | Alpha Kabinet KEITA et al |
| EPI_ISL_539574,EPI_ISL_539575,EPI_ISL_539576 | Centre de Recherches Medicales de Lambarene (CERMEL) | Department of Emerging Infectious Diseases, Institute of Tropical Medicine, Nagasaki University | Haruka Abe et al |
| EPI_ISL_428670,EPI_ISL_428672,EPI_ISL_428671,EPI_ISL_428673,EPI_ISL_525476,EPI_ISL_525478,EPI_ISL_525486,EPI_ISL_525488 | Centre for Dengue Research | Centre for Dengue Research | Chandima Jeewandara et al |
| EPI_ISL_525474 | Centre for Dengue Research | Centre for Dengue Research, USJ, SL | Chandima Jeewandara et al |
| EPI_ISL_792548,EPI_ISL_792555,EPI_ISL_862725,EPI_ISL_872568,EPI_ISL_978920,EPI_ISL_978922,EPI_ISL_978923,EPI_ISL_978987,EPI_ISL_1233056,EPI_ISL_1233058,EPI_ISL_1233103,EPI_ISL_1233109,EPI_ISL_1233110,EPI_ISL_1233113,EPI_ISL_1233115,EPI_ISL_1233119,EPI_ISL_1233134 | Centre for Dengue Research and AICBU, Department of Immunology and Molecular Medicine | Centre for Dengue Research and AICBU, Department of Immunology and Molecular Medicine | Chandima Jeewandara et al |
| EPI_ISL_602574,EPI_ISL_602566,EPI_ISL_602576,EPI_ISL_668451,EPI_ISL_668455,EPI_ISL_668453,EPI_ISL_668449 | Centre for Dengue Research, Department of Immunology and Molecular Medicine | Centre for Dengue Research, Department of Immunology and Molecular Medicine | Chandima Jeewandara et al |
| EPI_ISL_602565 | Centre for Dengue Research, Department of Immunology and Molecular Medicine | Centre for Dengue Research, Department of Immunology and Molecular Medicine, | Chandima Jeewandara et al |
| EPI_ISL_413550 | Centre for Human and Zoonotic Virology (CHAZVY), College of Medicine University of Lagos/Lagos University Teaching Hospital (LUTH), part of the Laboratory Network of the Nigeria Centre for Disease Control (NCDC) | African Centre of Excellence for Genomics of Infectious Diseases (ACEGID), Redeemer's University, Ede, Osun State, Nigeria | Oluniyi P.E. et al |
| EPI_ISL_654794 | Centre for Human Virology & Genomics, Nigerian Institute of Medical Research | Centre for Human Virology & Genomics, Nigerian Institute of Medical Research | Shaibu et al |
| EPI_ISL_413596 | Centre for Infectious Diseases and Microbiology - Public Health | NSW Health Pathology - Institute of Clinical Pathology and Medical Research; Westmead Hospital; University of Sydney | Rockett R et al |
| EPI_ISL_407893 | Centre for Infectious Diseases and Microbiology Laboratory Services | NSW Health Pathology - Institute of Clinical Pathology and Medical Research; Westmead Hospital; University of Sydney | Eden J-S et al |

| | | | |
|---|---|---|---|
| EPI_ISL_413594 | Centre for Infectious Diseases and Microbiology Laboratory Services | NSW Health Pathology - Institute of Clinical Pathology and Medical Research; Westmead Hospital; University of Sydney | Rockett R et al |
| EPI_ISL_427643 | Centre for Infectious Diseases and Microbiology Public Health | NSW Health Pathology - Institute of Clinical Pathology and Medical Research; Westmead Hospital; University of Sydney | Timms V et al |
| EPI_ISL_414624 | Centre Hositalier Universitaire de Rouen Laboratoire de Virologie | National Reference Center for Viruses of Respiratory Infections, Institut Pasteur, Paris | Mélnie Albert et al |
| EPI_ISL_900573 | Centre Hospitalier de Guéret | CNR Virus des Infections Respiratoires - France SUD | Antonin Bal et al |
| EPI_ISL_1250627 | Centre Hospitalier de l'Austreberthe | Centre Hospitalier Universitaire de Rouen Laboratoire de Virologie | Alice Moisan et al |
| EPI_ISL_508946 | Centre Hospitalier de Macon | CNR Virus des Infections Respiratoires - France SUD | Antonin Bal et al |
| EPI_ISL_414625 | Centre Hospitalier Régional Universitaire de Nantes Laboratoire de Virologie | National Reference Center for Viruses of Respiratory Infections, Institut Pasteur, Paris | Mélnie Albert et al |
| EPI_ISL_1299492 | Centre Hospitalo Universitaire Souro Sanou | Centre Muraz | Arsène Zongo et al |
| EPI_ISL_1015374 | Centre Marine La Villeneuve | IRBA, 2MI | GORGE O. et al |
| EPI_ISL_660442 | Centre Muraz | Project group Epidemology of Highly Pathogenic Microorganisms, Robert Koch Institut | Soumeya Ouangraoua et al |
| EPI_ISL_1279266 | Centro de Investigación Biomédica del Noreste (CIBIN) | Instituto Nacional de Enfermedades Respiratorias (INER): Centro de Investigación en Enfermedades Infecciosas (CIENI) | Consorcio Mexicano de Vigilancia Genómica (CoViGen-Mex). Authors (in alphabetical order): Julio Elias Alvarado-Yaah et al |
| EPI_ISL_941130 | Centro de Investigaciones en Microbiología y Biotecnología-UR (CIMBIUR), Facultad de Ciencias Naturales, Universidad del Rosario, Bogotá, Colombia | Centro de Investigaciones en Microbiología y Biotecnología-UR (CIMBIUR), Facultad de Ciencias Naturales, Universidad del Rosario, Bogotá, Colombia Icahn School of Medicine at Mount Sinai, New York, USA | Nathalia Ballesteros et al |
| EPI_ISL_941944,EPI_ISL_941945,EPI_ISL_941947,EPI_ISL_941995 | Centro de Investigaciones en Microbiología y Biotecnología-UR (CIMBIUR), Facultad de Ciencias Naturales, Universidad del Rosario, Bogotá, Colombia | Centro de Investigaciones en Microbiología y Biotecnología-UR (CIMBIUR), Facultad de Ciencias Naturales, Universidad del Rosario, Bogotá, Colombia Instituto Nacional de Salud, Bogotá, Colombia Icahn School of Medicine at Mount Sinai, New York, USA | Luz Helena Patiño et al |
| EPI_ISL_527819,EPI_ISL_697797 | Centro de Investigaciones, Universidad de Especialidades Espíritu Santo | Institute of Microbiology, Universidad San Francisco de Quito | Derly Andrade et al |
| EPI_ISL_837550,EPI_ISL_837554,EPI_ISL_837555,EPI_ISL_837556,EPI_ISL_837557,EPI_ISL_837558,EPI_ISL_837559,EPI_ISL_837560,EPI_ISL_837561,EPI_ISL_837562,EPI_ISL_837566,EPI_ISL_837567,EPI_ISL_837568,EPI_ISL_837571,EPI_ISL_837572,EPI_ISL_837574,EPI_ISL_837575,EPI_ISL_837576 | Centro Nacional de Enfermedades Tropicales (CENETROP) | Laboratory of Respiratory Viruses and Measles, Oswaldo Cruz Institute, FIOCRUZ | Paola Resende et al |
| EPI_ISL_512670,EPI_ISL_527755 | Centro Nacional De Rehabilitacion Humberto Araya Rojas (Cenare) | Inciensa, Instituto Costarricense de Investigación y Enseñanza en Nutrición y Salud | Francisco Duarte et al |
| EPI_ISL_1239537 | Centrum Medycyny Klinicznej i Estetycznej DiMedical Sp. z o.o. | 1. National Institute of Public Health - National Institute of Hygiene; 2. Eurofins Genomics Europe Sequencing GmbH | Wołkowicz Tomasz et al |
| EPI_ISL_751184 | CENUR Litoral Norte – UdelaR, Salto, Uruguay | Institut Pasteur de Montevideo | Daiana Mir et al |
| EPI_ISL_1229000 | Cerballiance | CNR Virus des Infections Respiratoires - France SUD | Antonin Bal et al |
| EPI_ISL_1265787,EPI_ISL_1265789 | CH BASTIA | CNR Virus des Infections Respiratoires - France SUD | Antonin Bal et al |
| EPI_ISL_1013406 | CH de Mayotte | National Reference Center for Viruses of Respiratory Infections, Institut Pasteur, Paris | Marion Barbet et al |
| EPI_ISL_860901 | CH du MANS - Lab. Bio. Moléculaire | National Reference Center for Viruses of Respiratory Infections, Institut Pasteur, Paris | Marion Barbet et al |
| EPI_ISL_416493 | CH Jean de Navarre Laboratoire de Biologie | National Reference Center for Viruses of Respiratory Infections, Institut Pasteur, Paris | Mélnie Albert et al |
| EPI_ISL_1209382 | CH ROUBAIX | CHU Lille - Laboratoire de Virologie | AIT YAHYA Emilie et al |
| EPI_ISL_1228999 | CH ROYAN | CNR Virus des Infections Respiratoires - France SUD | Antonin Bal et al |

| | | | |
|---|---|---|---|
| EPI_ISL_802995 | Charité Universitätsmedizin Berlin, Institut für Virologie, Charitéplatz 1, 10117 Berlin, Germany | Charité Universitätsmedizin Berlin, Institut für Virologie, Charitéplatz 1, 10117 Berlin, Germany | Victor M Corman et al |
| EPI_ISL_1164535,EPI_ISL_729475 | Charité Universitätsmedizin Berlin, Institut für Virologie/Labor Berlin | Charité Universitätsmedizin Berlin, Institut für Virologie | Victor M Corman et al |
| EPI_ISL_406862 | Charité Universitätsmedizin Berlin, Institute of Virology; Institut für Mikrobiologie der Bundeswehr, Munich | Charité Universitätsmedizin Berlin, Institute of Virology | Victor M Corman et al |
| EPI_ISL_1239284 | Charlotte Maxeke Johannesburg Academic Hospital, National Health Laboratory Services, Gauteng, South Africa | National Institute for Communicable Diseases of the National Health Laboratory Service | Amoako DG et al |
| EPI_ISL_1085538 | CHI VILLENEUVE ST GEORGES | Department of Virology, Henri Mondor University Hospital, Assistance Publique Hôpitaux de Paris, Université Paris-Est Créteil, INSERM U955 | Christophe Rodriguez et al |
| EPI_ISL_419310,EPI_ISL_901719 | Chiba Prefectural Institute of Public Health | Pathogen Genomics Center, National Institute of Infectious Diseases | Tsuyoshi Sekizuka et al |
| EPI_ISL_492028,EPI_ISL_625457,EPI_ISL_625458,EPI_ISL_625472,EPI_ISL_700328,EPI_ISL_700329,EPI_ISL_700331,EPI_ISL_700337,EPI_ISL_700338,EPI_ISL_700342,EPI_ISL_768731,EPI_ISL_906083,EPI_ISL_906092 | Child Health Research Foundation | Child Health Research Foundation | Senjuti Saha et al |
| EPI_ISL_1190785 | CHR LA REUNION FELIX GUYON | CNR Virus des Infections Respiratoires - France SUD | Antonin Bal et al |
| EPI_ISL_1085370 | CHR METZ THIONVILLE Hôpital de Mercy | Department of Virology, Henri Mondor University Hospital, Assistance Publique Hôpitaux de Paris, Université Paris-Est Créteil, INSERM U955 | Christophe Rodriguez et al |
| EPI_ISL_1013408,EPI_ISL_1013409,EPI_ISL_1013410 | CHU - Hôpital Cavale Blanche | National Reference Center for Viruses of Respiratory Infections, Institut Pasteur, Paris | Marion Barbet et al |
| EPI_ISL_779838,EPI_ISL_779844 | CHU Bordeaux | CNR Virus des Infections Respiratoires - France SUD | Antonin Bal et al |
| EPI_ISL_644680,EPI_ISL_663214,EPI_ISL_666714 | CHU de Limoges | CNR Virus des Infections Respiratoires - France SUD | Antonin Bal et al |
| EPI_ISL_644681,EPI_ISL_660693,EPI_ISL_660709 | CHU Montpellier | CNR Virus des Infections Respiratoires - France SUD | Antonin Bal et al |
| EPI_ISL_663237,EPI_ISL_663242,EPI_ISL_900531 | CHU Poitiers | CNR Virus des Infections Respiratoires - France SUD | Antonin Bal et al |
| EPI_ISL_1292807,EPI_ISL_1292808 | CHU Pontchaillou | CHU Pontchaillou | GROLHIER Claire et al |
| EPI_ISL_593874,EPI_ISL_671967,EPI_ISL_754137 | CHU Purpan - Laboratoire de Virologie - Institut Fédératif de Biologie | CHU Purpan - Laboratoire de Virologie - Institut Fédératif de Biologie | Latour J. et al |
| EPI_ISL_482888 | CHU Purpan - Laboratoire de Virologie - Institut Fédératif de Biologie | Laboratoire de virologie - École Nationale Vétérinaire de Toulouse | Guillaume Croville et al |
| EPI_ISL_660671 | CHU Toulouse | CNR Virus des Infections Respiratoires - France SUD | Antonin Bal et al |
| EPI_ISL_735391 | CHU Tours | CNR Virus des Infections Respiratoires - France SUD | Antonin Bal et al |
| EPI_ISL_1265619 | CHUGA-IBP-laboratoire de Virologie | IBP-laboratoire de virologie | Sylvie Larrat et al |
| EPI_ISL_745260 | CHUM-Site Glen-LAB Microbiologie | Laboratoire de santé publique du Québec | Sandrine Moreira et al |
| EPI_ISL_1299248 | CHWAPI - SITE NOTRE DAME | Institut de Pathologie et Genetique (IPG) | Pascale Hilbert et al |
| EPI_ISL_683835 | CICM | Malaria Research and Training Center (MRTC-Parasito) | Antoine Dara et al |
| EPI_ISL_487448,EPI_ISL_487453,EPI_ISL_487455,EPI_ISL_487457,EPI_ISL_487459,EPI_ISL_487460,EPI_ISL_487462,EPI_ISL_487464 | CICM-Mali | Bundeswehr Institut of Microbiology | Kouriba et al |
| EPI_ISL_1278404,EPI_ISL_1278453,EPI_ISL_1278520,EPI_ISL_1278521,EPI_ISL_1278522,EPI_ISL_1278535,EPI_ISL_1278849 | Clalit Health Services Laboratories, Israel | Stern Lab | Stern Lab et al |
| EPI_ISL_1250842,EPI_ISL_1278275,EPI_ISL_1278276 | Clin & Gen Lab | Molecular Genetics Laboratory, Instituto de Investigaciones Químicas, Universidad Mayor de San Andrés | Oscar M. Rollano-Peñaloza et al |
| EPI_ISL_462464,EPI_ISL_462470 | Clinical Center, University of Sarajevo | Charite Universitatsmedizin Berlin, Institute of Virology | Victor M Corman et al |
| EPI_ISL_1300657 | Clinical Center, University of Sarajevo; Unit for Clinical Microbiology | Clinical Center, University of Sarajevo; Unit for Clinical Microbiology | Irma Salimović-Bešić et al |

| | | | |
|---|---|---|---|
| EPI_ISL_747194,EPI_ISL_1122422,EPI_ISL_1122423 | Clinical Microbiology Laboratory, Faculty of Medicine, Universitas Indonesia | Clinical Microbiology Laboratory, Faculty of Medicine, Universitas Indonesia | Fera Ibrahim et al |
| EPI_ISL_1196798,EPI_ISL_1196795 | Clinical Microbiology Laboratory, Faculty of Medicine, Universitas Indonesia | Faculty of Medicine, Universitas Indonesia | Fadilah et al |
| EPI_ISL_447428,EPI_ISL_447446 | Clinical Microbiology Laboratory, Sheba Medical Center | Stern Lab | Stern Lab et al |
| EPI_ISL_447317,EPI_ISL_447325 | Clinical Virology Laboratory, Soroka Medical Center and the Faculty of Health Sciences, Ben-Gurion University of the Negev | Stern Lab | Stern Lab et al |
| EPI_ISL_1299497 | CMA Dano | Centre Muraz | Arsène Zongo et al |
| EPI_ISL_640001,EPI_ISL_640003,EPI_ISL_640009,EPI_ISL_683337,EPI_ISL_683391,EPI_ISL_683396,EPI_ISL_692757,EPI_ISL_730646,EPI_ISL_745375 | CNR Virus des Infections Respiratoires - France SUD | CNR Virus des Infections Respiratoires - France SUD | Antonin Bal et al |
| EPI_ISL_410486 | CNR Virus des Infections Respiratoires - France SUD | CNR Virus des Infections Respiratoires - France SUD | Bal et al |
| EPI_ISL_632903 | Communicable Disease Laboratory, Public Health Directorate | Communicable Disease Laboratory, Public Health Directorate | AlAbbas et al |
| EPI_ISL_681309,EPI_ISL_681301,EPI_ISL_681312,EPI_ISL_682303,EPI_ISL_682304,EPI_ISL_682305,EPI_ISL_682315,EPI_ISL_682316,EPI_ISL_682312,EPI_ISL_684035 | Communicable Disease Laboratory, Public Health Directorate | Communicable Disease Laboratory, Public Health Directorate | Alwasti et al |
| EPI_ISL_632255,EPI_ISL_632261,EPI_ISL_632283,EPI_ISL_632284,EPI_ISL_632262,EPI_ISL_632267,EPI_ISL_632263,EPI_ISL_632285 | Communicable Disease Laboratory, Public Health Directorate | Communicable Disease Laboratory, Public Health Directorate | AlWasti et al |
| EPI_ISL_510531 | Communicable Disease Laboratory, Public Health Directorate | Communicable Disease Laboratory, Public Health Directorate | Shehab et al |
| EPI_ISL_729778 | Connecticut Department of Health | Grubaugh Lab - Yale School of Public Health | Joseph Fauver et al |
| EPI_ISL_1040819,EPI_ISL_1040820,EPI_ISL_1040821 | CoVid WC Cape Town Metro | NHLS/UCT | Arash Iranzadeh et al |
| EPI_ISL_847827 | COVID-19 National Reference Laboratory | COVID-19 National Reference Laboratory | Tahmineh Jalali et al |
| EPI_ISL_1190754,EPI_ISL_1190757,EPI_ISL_1190758,EPI_ISL_1190763,EPI_ISL_1190764,EPI_ISL_1190765,EPI_ISL_1190767,EPI_ISL_1190768 | CREMER(Centre de Rechercherches sur les Maladies Emergentes et Ré-émergentes) | TransVIHMI(Recherches Translationnelles sur le VIH et les Maladies Infectieuses) | Celestin Godwe et al |
| EPI_ISL_1085405 | CRICQUEBOEUF Cerballiance Normandie | Department of Virology, Henri Mondor University Hospital, Assistance Publique Hôpitaux de Paris, Université Paris-Est Créteil, INSERM U955 | Christophe Rodriguez et al |
| EPI_ISL_1195673 | Croatian Institute of Public Health | Croatian Institute of Public Health | Irena Tabain et al |
| EPI_ISL_528850 | CSIR-Centre for Cellular and Molecular Biology | CSIR-Centre for Cellular and Molecular Biology | Lamuk Zaveri et al |
| EPI_ISL_910199,EPI_ISL_910242,EPI_ISL_910248,EPI_ISL_910258,EPI_ISL_910276,EPI_ISL_910278,EPI_ISL_910281 | CSIR-Centre for Cellular and Molecular Biology | CSIR-Centre for Cellular and Molecular Biology | Payel Mukherjee et al |
| EPI_ISL_661304,EPI_ISL_661309 | CSIR-Indian Institute of Chemical Biology, MEDICA Supercpecialty Hospital Kolkata | CSIR-Indian Institute of Chemical Biology, MEDICA Supercpecialty Hospital Kolkata | Sujay Krishna Maity et al |
| EPI_ISL_416542,EPI_ISL_421652 | Dasman Diabetes Institute | Dasman Diabetes Institute | Fahd Al-Mulla et al |
| EPI_ISL_1060924 | DB Diagnosticos do Brasil | Instituto de Medicina Tropical de Sao Paulo | Brazil-UK Centre for Arbovirus Discovery Diagnosis Genomics et al |

| | | | |
|---|---|---|---|
| EPI_ISL_722144 | DB Diagnosticos do Brasil | Laboratório de Parasitologia Médica - Instituto de Medicina Tropical - Universidade de São Paulo | Brazil-UK Centre for Arbovirus Discovery Diagnosis Genomics et al |
| EPI_ISL_804826 | DB Diagnosticos do Brasil | Laboratório de Parasitologia Médica - Instituto de Medicina Tropical - Universidade de São Paulo | Nuno Faria et al |
| EPI_ISL_476195,EPI_ISL_476279,EPI_ISL_476328 | DB Diagnósticos do Brasil | Instituto de Medicina Tropical da Univesidade de São Paulo | Samples: Nelson Gaburo Jr et al |
| EPI_ISL_828182,EPI_ISL_827757,EPI_ISL_829158,EPI_ISL_829968 | deCODE genetics | deCODE genetics | Daniel F Gudbjartsson et al |
| EPI_ISL_833041 | Defence Services Medical Research Center, Biological Research Laboratory | Defence Services Medical Research Center, Biological Research Laboratory | Khine Zaw Oo et al |
| EPI_ISL_792301 | Departamento de Biología y genética molecular, IACA Laboratorios. | Área de Secuenciación del Laboratorio de Virología del Hospital de Niños Dr. Ricardo Gutierrez on behalf of 'Proyecto Argentino Interinstitucional de genomica de SARS-CoV-2' (PAIS Consortium) | Nabaes Jodar et al |
| EPI_ISL_508666 | Departamento de Microbiología, CDB, Hospital Clínic, Barcelona | SeqCOVID-SPAIN consortium/IBV(CSIC) | Andrea Vergara et al |
| EPI_ISL_1137609 | Département de Maladies Infectieuses, CHU Farhat Hached Sousse, Tunisie | Laboratoire des Procédés de Criblage Moléculaire et Cellulaire-Centre de Biotechnologie de Sfax | Souissi et al |
| EPI_ISL_516922,EPI_ISL_516923,EPI_ISL_516925 | Department for Molecular Diagnostics, Centre for Medical Microbiology, Institute of Public Health of Montenegro | Charité Universitätsmedizin Berlin, Institut für Virologie | Victor M Corman et al |
| EPI_ISL_1013596,EPI_ISL_1013610 | Department for Molecular Diagnostics, Centre for Medical Microbiology, Institute of Public Health, Montenegro | Charité Universitätsmedizin Berlin, Institut für Virologie | Victor M Corman et al |
| EPI_ISL_481380 | Department for Virology, Molecular Biology and Genome Research, R. G. Lugar Center for Public Health Research, National Center for Disease Control and Public Health (NCDC) of Georgia. | Department for Virology, Molecular Biology and Genome Research, R. G. Lugar Center for Public Health Research, National Center for Disease Control and Public Health (NCDC) of Georgia. | Ana Papkiauri et al |
| EPI_ISL_470876,EPI_ISL_763062 | Department for Virology, Molecular Biology and Genome Research, R. G. Lugar Center for Public Health Research, National Center for Disease Control and Public Health (NCDC) of Georgia. | Department for Virology, Molecular Biology and Genome Research, R. G. Lugar Center for Public Health Research, National Center for Disease Control and Public Health (NCDC) of Georgia. | Giorgi Tomashvili et al |
| EPI_ISL_447056,EPI_ISL_470877,EPI_ISL_1048367 | Department for Virology, Molecular Biology and Genome Research, R. G. Lugar Center for Public Health Research, National Center for Disease Control and Public Health (NCDC) of Georgia. | Department for Virology, Molecular Biology and Genome Research, R. G. Lugar Center for Public Health Research, National Center for Disease Control and Public Health (NCDC) of Georgia. | Gvantsa Brachveli et al |
| EPI_ISL_420144 | Department for Virology, Molecular Biology and Genome Research, R. G. Lugar Center for Public Health Research, National Center for Disease Control and Public Health (NCDC) of Georgia. | Department for Virology, Molecular Biology and Genome Research, R. G. Lugar Center for Public Health Research, National Center for Disease Control and Public Health (NCDC) of Georgia. | Gvantsa Chanturia et al |
| EPI_ISL_447055,EPI_ISL_471529,EPI_ISL_754181 | Department for Virology, Molecular Biology and Genome Research, R. G. Lugar Center for Public Health Research, National Center for Disease Control and Public Health (NCDC) of Georgia. | Department for Virology, Molecular Biology and Genome Research, R. G. Lugar Center for Public Health Research, National Center for Disease Control and Public Health (NCDC) of Georgia. | Meri Pantsulaia et al |
| EPI_ISL_420140 | Department for Virology, Molecular Biology and Genome Research, R. G. Lugar Center for Public Health Research, National Center for Disease Control and Public Health (NCDC) of Georgia. | Department for Virology, Molecular Biology and Genome Research, R. G. Lugar Center for Public Health Research, National Center for Disease Control and Public Health (NCDC) of Georgia. | Nato Kotaria et al |
| EPI_ISL_754180 | Department for Virology, Molecular Biology and Genome Research, R. G. Lugar Center for Public Health Research, National Center for Disease Control and Public Health (NCDC) of Georgia. | Department for Virology, Molecular Biology and Genome Research, R. G. Lugar Center for Public Health Research, National Center for Disease Control and Public Health (NCDC) of Georgia. | Salome Javashvili et al |
| EPI_ISL_477169 | Department for Virology, Molecular Biology and Genome Research, R. G. Lugar Center for Public Health Research, National Center for Disease Control and Public Health (NCDC) of Georgia. | Department for Virology, Molecular Biology and Genome Research, R. G. Lugar Center for Public Health Research, National Center for Disease Control and Public Health (NCDC) of Georgia. | Tata Imnadze et al |

| | | | |
|---|---|---|---|
| EPI_ISL_515083,EPI_ISL_515084,EPI_ISL_515088,EPI_ISL_515091,EPI_ISL_515094,EPI_ISL_515099,EPI_ISL_515103,EPI_ISL_515110 | Department of Biochemistry, Cell and Molecular Biology | WACCBIP, University of Ghana | Ngoi et al |
| EPI_ISL_944717,EPI_ISL_944720 | Department of Biochemistry, Cell and Molecular Biology, West African Centre for Cell Biology of Infectious Pathogens (WACCBIP), University of Ghana | Department of Biochemistry, Cell and Molecular Biology, West African Centre for Cell Biology of Infectious Pathogens (WACCBIP), University of Ghana | Morang'a et al |
| EPI_ISL_884845,EPI_ISL_884843,EPI_ISL_884838,EPI_ISL_884856,EPI_ISL_884841 | Department of Biochemistry, Cell and Molecular Biology, West African Centre for Cell Biology of Infectious Pathogens (WACCBIP), University of Ghana | Department of Biochemistry, Cell and Molecular Biology, West African Centre for Cell Biology of Infectious Pathogens (WACCBIP), University of Ghana | Ngoi et al |
| EPI_ISL_907075,EPI_ISL_956332 | Department of Biology, University of Basrah | Department of Biology, University of Basrah | Abu-Ali et al |
| EPI_ISL_626299,EPI_ISL_424648,EPI_ISL_484707,EPI_ISL_707745,EPI_ISL_707746 | Department of Clinical Microbiology | GIGA Medical Genomics | Keith Durkin et al |
| EPI_ISL_419228,EPI_ISL_419227 | Department of Clinical Pathology, Pamela Youde Nethersole Eastern Hospital | Department of Health Technology and Informatics, Faculty of Health and Social Science, The Hong Kong Polytechnic University | Kenneth Siu-Sing LEUNG et al |
| EPI_ISL_419242 | Department of Clinical Pathology, Tuen Mun Hospital | Department of Health Technology and Informatics, Faculty of Health and Social Science, The Hong Kong Polytechnic University | Kenneth Siu-Sing LEUNG et al |
| EPI_ISL_419231 | Department of Clinical Pathology, Tuen Mun Hospital, 23 Tsing Chung Koon Road, Tuen Mun, N.T. | Department of Health Technology and Informatics, Faculty of Health and Social Science, The Hong Kong Polytechnic University | Kenneth Siu-Sing LEUNG et al |
| EPI_ISL_1018120,EPI_ISL_1018121,EPI_ISL_1018258,EPI_ISL_1018285,EPI_ISL_1018304,EPI_ISL_1018317,EPI_ISL_1019640,EPI_ISL_1019707,EPI_ISL_1019710,EPI_ISL_1020103,EPI_ISL_1020214 | Department of Health Technology and Informatics, The Hong Kong Polytechnic University | Department of Health Technology and Informatics, The Hong Kong Polytechnic University | Gilman Kit-Hang Siu et al |
| EPI_ISL_610211 | Department of Health Technology and Informatics, The Hong Kong Polytechnic University | Department of Health Technology and Informatics, The Hong Kong Polytechnic University | Siu et al |
| EPI_ISL_417444 | Department of Healthcare Biotechnology, National University of Sciences and Technology (NUST) | Department of Healthcare Biotechnology, National University of Sciences and Technology (NUST) | Javed et al |
| EPI_ISL_411220 | Department of Infectious and Tropical Diseases, Bichat Claude Bernard Hospital, Paris | Laboratoire Virpath, CIRI U111, UCBL1, INSERM, CNRS, ENS Lyon | Olivier Terrier et al |
| EPI_ISL_1255104 | Department of Infectious Disease Prevention and Control, Henan Provincial Center for Disease Control and Prevention | Department of Infectious Disease Prevention and Control, Henan Provincial Center for Disease Control and Prevention | Li et al |
| EPI_ISL_477180 | Department of Laboratory Medicine Tan Tock Seng Hospital | Department of Laboratory Medicine Tan Tock Seng Hospital | Chen YYC et al |
| EPI_ISL_934554,EPI_ISL_934569 | Department of Laboratory Medicine, Division of Clinical Virology, University of Medicine, Vienna | Bergthaler laboratory, CeMM Research Center for Molecular Medicine of the Austrian Academy of Sciences | Lukas Endler et al |
| EPI_ISL_410218,EPI_ISL_408489,EPI_ISL_447621,EPI_ISL_534336,EPI_ISL_693303,EPI_ISL_693304,EPI_ISL_738065,EPI_ISL_1010728,EPI_ISL_1039160,EPI_ISL_1041957 | Department of Laboratory Medicine, National Taiwan University Hospital | Microbial Genomics Core Lab, National Taiwan University Centers of Genomic and Precision Medicine | Shiou-Hwei Yeh et al |
| EPI_ISL_648043,EPI_ISL_648071 | Department of Laboratory Medicine, Tan Tock Seng Hospital | Department of Laboratory Medicine, Tan Tock Seng Hospital | Chen YYC et al |
| EPI_ISL_977590 | Department of Medical Microbiology, Hospital Pengajar Universiti Putra Malaysia | Malaysia Genome Institute | Mohd Noor Mat Isa et al |
| EPI_ISL_775346 | Department of medical microbiology, section Aalesund, Aalesund Hospital | Norwegian Institute of Public Health, Department of Virology | Kathrine Stene-Johansen et al |
| EPI_ISL_501176,EPI_ISL_501177,EPI_ISL_501185,EPI_ISL_501204,EPI_ISL_501207,EPI_ISL_501220 | Department of Medical Microbiology, University Malaya Medical Centre | Department of Medical Microbiology, Faculty of Medicine, University of Malaya | Yoong Min CHONG et al |
| EPI_ISL_512844 | Department of Medical Research | DMR_Myanmar | Myat Htut Nyunt et al |

| | | | |
|---|---|---|---|
| EPI_ISL_431117 | Department of Microbiology, Gandhi Medical College and Hospital, Secendrabad, Hyderabad, India | Department of Microbiology, Gandhi Medical College and Hospital, Secendrabad, Hyderabad, India | Thrilok Chander B et al |
| EPI_ISL_1027646,EPI_ISL_1027645,EPI_ISL_1027649 | Department of Microbiology, National Institute for Public Health of Kosova | Charité Universitätsmedizin Berlin, Institut für Virologie | Victor M Corman et al |
| EPI_ISL_498271,EPI_ISL_498270,EPI_ISL_497865,EPI_ISL_497818,EPI_ISL_497805,EPI_ISL_497797,EPI_ISL_497788,EPI_ISL_497823,EPI_ISL_497827,EPI_ISL_1034417,EPI_ISL_1034434,EPI_ISL_1034435,EPI_ISL_1034436,EPI_ISL_1034437,EPI_ISL_1034440,EPI_ISL_1034446,EPI_ISL_1034604,EPI_ISL_1034710,EPI_ISL_1197084,EPI_ISL_1197100,EPI_ISL_1197102,EPI_ISL_1197106,EPI_ISL_1197109 | Department of Microbiology, The University of Hong Kong | Department of Microbiology, The University of Hong Kong | Kelvin K.W. To et al |
| EPI_ISL_1008286,EPI_ISL_1180897 | Department of Microbiology, University Innsbruck | Bergthaler laboratory, CeMM Research Center for Molecular Medicine of the Austrian Academy of Sciences | Lukas Endler et al |
| EPI_ISL_463741,EPI_ISL_1164627,EPI_ISL_1164630,EPI_ISL_1164631,EPI_ISL_1164639,EPI_ISL_1164641,EPI_ISL_1164643,EPI_ISL_1164647,EPI_ISL_1164653,EPI_ISL_1164670,EPI_ISL_1164688,EPI_ISL_1164702,EPI_ISL_1164733,EPI_ISL_1164734 | Department of Molecular Virology, Cyprus Institute of Neurology and Genetics | Department of Molecular Virology, Cyprus Institute of Neurology and Genetics | Jan Richter et al |
| EPI_ISL_853819 | Department of Pathology, Landeskrankenhaus Graz II, Medical University Graz | Bergthaler laboratory, CeMM Research Center for Molecular Medicine of the Austrian Academy of Sciences | Lukas Endler et al |
| EPI_ISL_594185,EPI_ISL_594186,EPI_ISL_594187,EPI_ISL_594188,EPI_ISL_596452,EPI_ISL_596451,EPI_ISL_596455 | Department of Pathology, School of Medicine, Imam Khomeini Hospital, Tehran University of Medical Sciences | Genetics Research Center, University of Social Welfare and Rehabilitation Sciences | Zohreh Fattahi et al |
| EPI_ISL_582033 | Department of Pathology, School of Medicine, Imam Khomeini Hospital, Tehran University of Medical Sciences | Genetics Research Center. University Of Social Welfare And Rehabilitation Sciences | Zohreh Fattahi et al |
| EPI_ISL_425340 | Department of Pathology, University of Cambridge | COVID-19 Genomics UK (COG-UK) Consortium | Luke W Meredith et al |
| EPI_ISL_1289914 | Department of Public Health Microbiology Ljubljana, National Laboratory for Health, Environment and Food | Department of Public Health Microbiology Ljubljana, National Laboratory for Health, Environment and Food | José Gonçalves et al |
| EPI_ISL_576147 | Department of Respiratory & Other Viral Infections of L.V. Gromashevsky Institute of Epidemiology & Infectious Diseases NAMS of Ukraine | Department of Respiratory & Other Viral Infections of L.V. Gromashevsky Institute of Epidemiology & Infectious Diseases NAMS of Ukraine, JSC Farmak"" | Alla Mironenko et al |
| EPI_ISL_582512,EPI_ISL_654819,EPI_ISL_1122014 | Department of Respiratory and other Viral Infections of L.V.Gromashevsky Institute of Epidemiology & Infectious Diseases NAMS of Ukraine | Department of Respiratory and other Viral Infections of L.V.Gromashevsky Institute of Epidemiology & Infectious Diseases NAMS of Ukraine, JSC Farmak"" | Alla Mironenko et al |
| EPI_ISL_737035,EPI_ISL_737211,EPI_ISL_738134,EPI_ISL_862054,EPI_ISL_862066,EPI_ISL_862102,EPI_ISL_1240105,EPI_ISL_1240166,EPI_ISL_1240690,EPI_ISL_1240691 | Department of Virology and Immunology, University of Helsinki and Helsinki University Hospital, Huslab Finland | Department of Virology, Faculty of Medicine, University of Helsinki, Helsinki, Finland | Teemu Smura et al |
| EPI_ISL_407084 | Department of Virology III, National Institute of Infectious Diseases | Pathogen Genomics Center, National Institute of Infectious Diseases | Tsuyoshi Sekizuka et al |
| EPI_ISL_1273443 | Department of Virology, Bangabandhu Sheikh Mujib Medical University | Bioinformatics Division, National Institute of Biotechnology (NIB) | Munira Jahan et al |

| | | | |
|---|---|---|---|
| EPI_ISL_1015001,EPI_ISL_1015153,EPI_ISL_1015272,EPI_ISL_1015275,EPI_ISL_1015304,EPI_ISL_1015324 | Department of Virology, Pitié-Salpêtrière hospital | Department of Virology, Pitié-Salpêtrière hospital | Valentin Leducq et al |
| EPI_ISL_855561,EPI_ISL_855568 | Department of Virology, Principal Military Hospital of Instruction of Tunis | Bundeswehr Institute of Microbiology | Susann Handrick et al |
| EPI_ISL_793253 | Department of Virus and Microbiological Special Diagnostics, Statens Serum Institut, Copenhagen, Denmark | Albertsen Lab, Department of Chemistry and Bioscience, Aalborg University, Denmark | Danish Covid-19 Genome Consortium et al |
| EPI_ISL_429413 | Department of Virus and Microbiological Special Diagnostics, Statens Serum Institut, Copenhagen, Denmark, Artillerivej 5, 2300 Copenahgen S | Albertsen lab, Department of Chemistry and Bioscience, Aalborg University, Denmark | Rasmus Kirkegaard et al |
| EPI_ISL_416142 | Department of Virus and Microbiological Special diagnostics, Statens Serum Institut, Copenhagen, Denmark. | Statens Serum Institute | Morten Rasmussen et al |
| EPI_ISL_416143 | Department of Virus and Microbiological Special diagnostics, Statens Serum Institut, Copenhagen, Denmark. | ViFU | Morten Rasmussen et al |
| EPI_ISL_615335,EPI_ISL_619762,EPI_ISL_621148,EPI_ISL_622417 | Department of Virus and Microbiological Special Diagnostics, Statens Serum Institut, Denmark | Albertsen lab, Department of Chemistry and Bioscience, Aalborg University, Denmark | Danish Covid-19 Genome Consortia et al |
| EPI_ISL_664103 | Dept. of Microbiology and Infection Control, Akershus University Hospital HF | Dept. of Microbiology and Infection Control, Akershus University Hospital HF | Hege Vangstein Aamot et al |
| EPI_ISL_410532,EPI_ISL_410531 | Dept. of Pathology, National Institute of Infectious Diseases | Pathogen Genomics Center, National Institute of Infectious Diseases | Tsuyoshi Sekizuka et al |
| EPI_ISL_774874 | Designated Reference Institute for Chemical Measurements (DRICM) | DNA SOLUTION LTD. | Md. Imran Khan et al |
| EPI_ISL_1020197 | Dinkes, Bogor, West Java | Biosafety Level-3 Laboratory, Indonesian Institute of Sciences (LIPI) | Anik Budhi Dharmayanthi et al |
| EPI_ISL_1137618 | Dirección de Sanidad Policia Nacional | Instituto Nacional de Salud- Dirección de Investigación en Salud Pública | Katherine Laiton-Donato et al |
| EPI_ISL_428479 | District Surveillence Unit | Department of Neurovirology, National Institute of Mental Health and Neuroscience (NIMHANS) | Chitra Pattabiraman et al |
| EPI_ISL_747261,EPI_ISL_747315,EPI_ISL_747316,EPI_ISL_747338,EPI_ISL_747344,EPI_ISL_747349,EPI_ISL_747353,EPI_ISL_747354,EPI_ISL_760063,EPI_ISL_760066,EPI_ISL_760115,EPI_ISL_760131,EPI_ISL_760133,EPI_ISL_850192,EPI_ISL_850212,EPI_ISL_850228,EPI_ISL_850654,EPI_ISL_850656,EPI_ISL_850657,EPI_ISL_955947,EPI_ISL_1063606,EPI_ISL_1063633,EPI_ISL_1063668,EPI_ISL_1165006,EPI_ISL_1165030,EPI_ISL_1165038,EPI_ISL_1165050,EPI_ISL_1165053,EPI_ISL_1165054,EPI_ISL_1165063,EPI_ISL_1252405,EPI_ISL_1252409,EPI_ISL_1252418 | Division of Emerging Infectious Diseases, Bureau of Infectious Diseases Diagnosis Control, Korea Disease Control and Prevention Agency | Division of Emerging Infectious Diseases, Bureau of Infectious Diseases Diagnosis Control, Korea Disease Control and Prevention Agency | Ae Kyung Park et al |
| EPI_ISL_812963 | Division of Pathogen Resource Management, Korea National Institute of Health, Korea Disease Control and Prevention Agency | Division of Pathogen Resource Management, Korea National Institute of Health, Korea Disease Control and Prevention Agency | Kim et al |
| EPI_ISL_412872,EPI_ISL_426164,EPI_ISL_426166,EPI_ISL_498034,EPI_ISL_498035,EPI_ISL_506972,EPI_ISL_506980,EPI_ISL_506994,EPI_ISL_514961,EPI_ISL_522474 | Division of Viral Diseases, Center for Laboratory Control of Infectious Diseases, Korea Centers for Diseases Control and Prevention | Division of Viral Diseases, Center for Laboratory Control of Infectious Diseases, Korea Centers for Diseases Control and Prevention | Jeong-Min Kim et al |
| EPI_ISL_583734,EPI_ISL_583846 | Dr. Gernot Walder GmbH | Bergthaler laboratory, CeMM Research Center for Molecular Medicine of the Austrian Academy of Sciences | Alexandra Popa et al |

| | | | |
|---|---|---|---|
| EPI_ISL_722508,EPI_ISL_722538,EPI_ISL_722539,EPI_ISL_722541,EPI_ISL_722706,EPI_ISL_763161,EPI_ISL_422933,EPI_ISL_460858,EPI_ISL_460991,EPI_ISL_523165,EPI_ISL_523535,EPI_ISL_577876,EPI_ISL_577928 | Dutch COVID-19 response team | Erasmus Medical Center | Bas Oude Munnink et al |
| EPI_ISL_1120177,EPI_ISL_1120178,EPI_ISL_1120495,EPI_ISL_1120517,EPI_ISL_1158727,EPI_ISL_1210393 | Dutch COVID-19 response team | Medical Microbiology, Maastricht University Medical Centre | Jozef Dingemans* et al |
| EPI_ISL_905532,EPI_ISL_1288926,EPI_ISL_547445,EPI_ISL_547446,EPI_ISL_547447,EPI_ISL_547448,EPI_ISL_547449,EPI_ISL_547450,EPI_ISL_547452,EPI_ISL_636493,EPI_ISL_636513,EPI_ISL_636514,EPI_ISL_636515,EPI_ISL_636516,EPI_ISL_636517,EPI_ISL_636518,EPI_ISL_636521,EPI_ISL_904762,EPI_ISL_1014269,EPI_ISL_1014280,EPI_ISL_1014281,EPI_ISL_1014282,EPI_ISL_1014552,EPI_ISL_1014557,EPI_ISL_1014558,EPI_ISL_1014561,EPI_ISL_1014571,EPI_ISL_1014572,EPI_ISL_1014573,EPI_ISL_1014574,EPI_ISL_1014575,EPI_ISL_1014576,EPI_ISL_1014588,EPI_ISL_1014594,EPI_ISL_1035061,EPI_ISL_1035238,EPI_ISL_1035240,EPI_ISL_1035241,EPI_ISL_1035245,EPI_ISL_1035247,EPI_ISL_1035764,EPI_ISL_1035766,EPI_ISL_1035769,EPI_ISL_1059905,EPI_ISL_1165526,EPI_ISL_1165527,EPI_ISL_1165529,EPI_ISL_1165530,EPI_ISL_1232247,EPI_I | Dutch COVID-19 response team | National Institute for Public Health and the Environment (RIVM) | Adam Meijer et al |
| EPI_ISL_961803 | E. Gulbja laboratorija | Latvian Biomedical Research and Study Centre | Janis Pjalkovskis et al |
| EPI_ISL_492993 | E. Gulbja Laboratorija | Latvian Biomedical Research and Study Centre | Ivars Silamiķelis et al |
| EPI_ISL_593534,EPI_ISL_770826 | Eastern Ontario Regional Laboratory Association | McMaster University | Leanne Mortimer et al |
| EPI_ISL_1238792 | Ecological and Evolutionary Genomics. UGA-CINVESTAV | Unidad Universitaria de Secuenciación Masiva y Bioinformática (UUSMB). IBT-UNAM | Angelica Cibrian et al |
| EPI_ISL_475724 | Egyptian National Cancer Institute (ENCI) | Egyptian National Cancer Institute (ENCI) | Zekri et al |
| EPI_ISL_1122015,EPI_ISL_1121993 | Emergency department of COVID-19, Beijing Ditan Hospital | National Institute for Viral Disease Control and Prevention | Yang Song2* et al |
| EPI_ISL_708826,EPI_ISL_708822 | Emergency Operation Center, (EOC) | National Institute of Health, Department of Medical Sciences, Ministry of Public Health, Thailand | Pilailuk Okada et al |
| EPI_ISL_1273097 | Faculty of Medicine, Al-Quds University | Faculty of Medicine, Al-Quds University | Ereqat et al |
| EPI_ISL_451970 | Federal Budget Institution of Science, State Research Center for Applied Microbiology & Biotechnology | Federal Budget Institution of Science, State Research Center for Applied Microbiology & Biotechnology | Dyatlov I et al |
| EPI_ISL_419559 | FL Bureau of Public Health Laboratories-Tampa | Pathogen Discovery, Respiratory Viruses Branch, Division of Viral Diseases, Centers for Disease Control and Prevention | Anna Uehara et al |
| EPI_ISL_508768,EPI_ISL_549256 | Florida Bureau of Public Health Laboratories | Florida Bureau of Public Health Laboratories | Sarah Schmedes et al |
| EPI_ISL_541194 | Florida Bureau of Public Health Laboratories, Florida Department of Health | Florida Bureau of Public Health Laboratories, Florida Department of Health | Schmedes et al |
| EPI_ISL_476139 | Folkhalsomyndigheten | The Public Health Agency of Sweden | Oskar Karlsson Lindsjo et al |

| | | | |
|---|---|---|---|
| EPI_ISL_581455,EPI_ISL_581487,EPI_ISL_581489,EPI_ISL_581490,EPI_ISL_581491,EPI_ISL_581492,EPI_ISL_581493 | Fondation Congolaise pour la recherche medicale (FCRM) | NGS Competence Center Tübingen, Institut für Medizinische Mikrobiologie und Hygiene, Universitätsklinikum Tübingen | Angel Angelov et al |
| EPI_ISL_912361,EPI_ISL_912376,EPI_ISL_912377,EPI_ISL_912378,EPI_ISL_912379,EPI_ISL_912380,EPI_ISL_912390,EPI_ISL_912391 | Fondation Congolaise pour la recherche medicale (FCRM), Francine Ntoumi | NGS Competence Center Tuebingen, Institut für Medizinische Mikrobiologie und Hygiene, Universitaetsklinikum Tübingen | Angel Angelov et al |
| EPI_ISL_1210559,EPI_ISL_1210584,EPI_ISL_1210589 | FSBI «NATIONAL MEDICAL RESEARCH CENTER FOR OBSTETRICS, GYNECOLOGY AND PERINATOLOGY NAMED AFTER ACADEMICIAN V.I.KULAKOV» MINISTRY OF HEALTHCARE OF THE RUSSIAN FEDERATION | Center for Precision Genome Editing and Genetic Technologies for Biomedicine, Pirogov Medical University, Moscow, Russian Federation | Yegor Botsmanov et al |
| EPI_ISL_428851 | FSBSI Chumakov Federal Scientific Center for Research and Development of Immune-and-Biological Products of Russian Academy of Sciences"" | FSBSI Chumakov Federal Scientific Center for Research and Development of Immune-and-Biological Products of Russian Academy of Sciences" & NRC "Kurchatov institute"" | Liubov Kozlovskaya et al |
| EPI_ISL_431240,EPI_ISL_431180 | Fujian Center for Disease Control and Prevention | Fujian Center for Disease Control and Prevention | Lin Qi et al |
| EPI_ISL_1239137,EPI_ISL_1239138 | Fundação Ezequiel Dias | Coordenação Geral de Laboratórios de Saúde Pública (CGLAB) | Vagner Fonseca et al et al |
| EPI_ISL_1186078,EPI_ISL_1251297 | Furst Medical Laboratory | Norwegian Institute of Public Health, Department of Virology | Kathrine Stene-Johansen et al |
| EPI_ISL_661287 | Gavle klinisk mikrobiologi | The Public Health Agency of Sweden | Department of Microbiology et al |
| EPI_ISL_730567,EPI_ISL_730620,EPI_ISL_984987 | Gazi University Faculty of Medicine, Medical Virology Laboratory | Gazi University Faculty of Medicine, Medical Virology Laboratory | Erdem Şahin et al |
| EPI_ISL_1185245,EPI_ISL_1184582 | Genelabs Medical (Pvt) Ltd | Genelabs Medical (Pvt) Ltd | Chandanamali Punchihewa et al |
| EPI_ISL_1014934,EPI_ISL_1018115,EPI_ISL_1138410 | General Hospital - Ohrid | Research Center for Genetic Engineering and Biotechnology Georgi D. Efremov" , Macedonian Academy of Sciences and Arts" | Aleksandar J. Dimovski et al |
| EPI_ISL_1014936 | General Hospital - Prilep | Research Center for Genetic Engineering and Biotechnology Georgi D. Efremov" , Macedonian Academy of Sciences and Arts" | Aleksandar J. Dimovski et al |
| EPI_ISL_677674 | General Hospital - Prilep | Research Center for Genetic Engineering and Biotechnology Georgi D. Efremov" , Macedonian Academy of Sciences and Arts" | RCGEB - MASA et al |
| EPI_ISL_678254 | General Hospital - Strumica | Research Center for Genetic Engineering and Biotechnology Georgi D. Efremov" , Macedonian Academy of Sciences and Arts" | RCGEB - MASA et al |
| EPI_ISL_406801,EPI_ISL_406798 | General Hospital of Central Theater Command of People's Liberation Army of China | BGI & Institute of Microbiology, Chinese Academy of Sciences & Shandong First Medical University & Shandong Academy of Medical Sciences & General Hospital of Central Theater Command of People's Liberation Army of China | Weijun Chen et al |
| EPI_ISL_614282,EPI_ISL_614283,EPI_ISL_623097,EPI_ISL_693774 | General practitioner | National Reference Center for Viruses of Respiratory Infections, Institut Pasteur, Paris | Marion Barbet et al |
| EPI_ISL_888828 | General practitioner office | National Reference Center for Viruses of Respiratory Infections, Institut Pasteur, Paris | Marion Barbet et al |

| | | | |
|---|---|---|---|
| EPI_ISL_746504,EPI_ISL_746598,EPI_ISL_746601,EPI_ISL_746639,EPI_ISL_746728,EPI_ISL_746729,EPI_ISL_746744,EPI_ISL_746771,EPI_ISL_1167671,EPI_ISL_1167691,EPI_ISL_1167765,EPI_ISL_1167799,EPI_ISL_1167804,EPI_ISL_1167812,EPI_ISL_1167830,EPI_ISL_1167832,EPI_ISL_1167925,EPI_ISL_1167927,EPI_ISL_1167931,EPI_ISL_1167932,EPI_ISL_1300469,EPI_ISL_1300470,EPI_ISL_1300479,EPI_ISL_1300504,EPI_ISL_1300510,EPI_ISL_1300519 | Genetica Molecular and Subdepartamento de Virologia ISP Chile | Instituto de Salud Publica de Chile | Javier Tognarelli et al |
| EPI_ISL_480243,EPI_ISL_480267,EPI_ISL_480283,EPI_ISL_480290,EPI_ISL_735259,EPI_ISL_735276 | Genomic Laboratory (GLAB) (Conjoint lab of Health Directorate of Istanbul and Istanbul Technical University) | Genomic Laboratory (GLAB), Istanbul Technical University | Ilker Karacan et al |
| EPI_ISL_632908 | Genomic Sciences, Rehman Medical Institute | Genomic Sciences, Rehman Medical Institute | Ali et al |
| EPI_ISL_812837,EPI_ISL_812809,EPI_ISL_812814,EPI_ISL_812858,EPI_ISL_812786,EPI_ISL_812868,EPI_ISL_812869 | Genomics Program, Children Cancer Hospital | Genomics Program, Children Cancer Hospital | Hatem et al |
| EPI_ISL_1220317,EPI_ISL_1220321 | GH BORDEAUX | CNR Virus des Infections Respiratoires - France SUD | Antonin Bal et al |
| EPI_ISL_1265821 | GHE REUNION | CNR Virus des Infections Respiratoires - France SUD | Antonin Bal et al |
| EPI_ISL_775214 | Gonoshasthya-RNA Molecular Research Center | Gonoshasthya-RNA Molecular Research Center | Mohd. Raeed Jamiruddin et al |
| EPI_ISL_906306 | Gorgas memorial Institute For Health Studies | Gorgas memorial Institute For Health Studies | Diaz Y et al |
| EPI_ISL_496605,EPI_ISL_496606,EPI_ISL_496607,EPI_ISL_496609,EPI_ISL_496615,EPI_ISL_496617,EPI_ISL_496647,EPI_ISL_496651,EPI_ISL_496683,EPI_ISL_496736,EPI_ISL_496840,EPI_ISL_496907 | Gorgas Memorial Laboratory of Health Studies | Gorgas Memorial Laboratory of Health Studies | Danilo Franco et al |
| EPI_ISL_1225381,EPI_ISL_1225382,EPI_ISL_1225392,EPI_ISL_1225393,EPI_ISL_1225400,EPI_ISL_1225420,EPI_ISL_1225431,EPI_ISL_1225457,EPI_ISL_1225506,EPI_ISL_1225520,EPI_ISL_1225537,EPI_ISL_1225544,EPI_ISL_1225545,EPI_ISL_1225548,EPI_ISL_1225550,EPI_ISL_1225560,EPI_ISL_1225572,EPI_ISL_1225574,EPI_ISL_1225577,EPI_ISL_1225578,EPI_ISL_1225580,EPI_ISL_1225581,EPI_ISL_1225582 | Gorgas Memorial Laboratory of Health Studies | Gorgas Memorial Laboratory of Health Studies | Yamilka Diaz et al |
| EPI_ISL_495024 | Government Medical College, Bhavnagar | Gujarat Biotechnology Research Centre | Raghawendra Kumar et al |
| EPI_ISL_794819 | Greek Genome Center, Biomedical Research Foundation of the Academy of Athens (BRFAA) | Greek Genome Center, Biomedical Research Foundation of the Academy of Athens (BRFAA) | Emmanouil Athanasiadis et al |
| EPI_ISL_640040,EPI_ISL_640045,EPI_ISL_640051,EPI_ISL_640058 | Groote Schuur Hospital wc GSH | NHLS/UCT | Arash Iranzadeh et al |
| EPI_ISL_698143,EPI_ISL_698144,EPI_ISL_698277,EPI_ISL_699147 | Group 42 (G42) Healthcare, Abu Dhabi, United Arab Emirates; Department of Health, The United Arab Emirates | G42 Healthcare | Rong Liu et al |

| | | | |
|---|---|---|---|
| EPI_ISL_1040923,EPI_ISL_1040924,EPI_ISL_1040925 | Grupo de Investigación en Enfermedades Tropicales del Ejército (GINETEJ), Laboratorio de Referencia e Investigación, Dirección de Sanidad Ejército, Bogotá, Colombia | Centro de Investigaciones en Microbiología y Biotecnología-UR (CIMBIUR), Facultad de Ciencias Naturales, Universidad del Rosario, Bogotá, Colombia | Juan David Ramírez et al |
| EPI_ISL_447749 | Grupo de Investigaciones Microbiológicas-UR (GIMUR), Departamento de Biología, Facultad de Ciencias Naturales, Universidad del Rosario, Bogotá, Colombia | Grupo de Investigaciones Microbiológicas-UR (GIMUR), Departamento de Biología, Facultad de Ciencias Naturales, Universidad del Rosario, Bogotá, Colombia Instituto Nacional de Salud, Bogotá, Colombia Icahn School of Medicine at Mount Sinai, New York, USA | Juan David Ramírez et al |
| EPI_ISL_1273049,EPI_ISL_1273060,EPI_ISL_1273064,EPI_ISL_1273063,EPI_ISL_1273057,EPI_ISL_1273052,EPI_ISL_1273071,EPI_ISL_1273073 | Guam Public Health Laboratory | Centers for Disease Control and Prevention Division of Viral Diseases, Pathogen Discovery | Krista Queen et al |
| EPI_ISL_509710 | Guatemala Ministry of Public Health | Pathogen Discovery, Respiratory Viruses Branch, Division of Viral Diseases, Centers for Disease Control and Prevention | Jing Zhang et al |
| EPI_ISL_509696,EPI_ISL_509697,EPI_ISL_509698,EPI_ISL_509699,EPI_ISL_509700,EPI_ISL_509703 | Guatemala Ministry of Public Health | Pathogen Discovery, Respiratory Viruses Branch, Division of Viral Diseases, Centers for Disease Control and Prevention | Ying Tao et al |
| EPI_ISL_482582 | Hangzhou Center for Diseases Control and Prevention | Hangzhou Center for Diseases Control and Prevention | Jun Li et al |
| EPI_ISL_796021,EPI_ISL_796012,EPI_ISL_796016 | Hebei Provincial Center for Disease Control and Prevention, Shijiazhuang, Hebei Province; National Institute for Viral Disease Control and Prevention, China CDC | Hebei Provincial Center for Disease Control and Prevention, Shijiazhuang, Hebei Province; National Institute for Viral Disease Control and Prevention, China CDC | Shunxiang Qi et al |
| EPI_ISL_733433,EPI_ISL_733452,EPI_ISL_873038 | HELIX LLC | WHO National Influenza Centre Russian Federation | Andrey Komissarov et al |
| EPI_ISL_501250,EPI_ISL_501249 | Hellenic Pasteur Institute, National Influenza Reference laboratory of Southern Greece & Unit of Bioinformatics and Applied Genomics | Hellenic Pasteur Institute, National Influenza Reference laboratory of Southern Greece & Unit of Bioinformatics and Applied Genomics | Vasiliki Pogka et al |
| EPI_ISL_699810,EPI_ISL_699881,EPI_ISL_700031 | Hematopathology Laboratory, ACTREC, TMC | Hematopathology Laboratory, ACTREC, TMC | Hematopathology Laboratory et al |
| EPI_ISL_1170956 | HIV Molecular Lab, Ethiopian Public Health Institute | HIV Molecular Lab | Weldemariam et al |
| EPI_ISL_1170957 | HIV Molecular laboratory, Ethiopian Public Health Institute | HIV Molecular Laboratory | Weldemariam et al |
| EPI_ISL_1169919 | HIV Molecular Laboratory, Ethiopian Public Health Institute | HIV Molecular Laboratory, Ethiopian Public Health Institute | Weldemariam et al |
| EPI_ISL_1170958 | HIV Molecular Laboratory, Ethiopian Public Health Institute, Ethiopia | HIV molecular lab, Ethiopian Public Health Institute, Ethiopian | Weldemariam et al |
| EPI_ISL_1196011,EPI_ISL_1196012,EPI_ISL_1196014 | Homecare | National Institute for Communicable Diseases of the National Health Laboratory Service | Maphalala GP et al |
| EPI_ISL_414571 | Hong Kong Department of Health | School of Public Health, The University of Hong Kong | Dominic N.C. Tsang et al |
| EPI_ISL_1001389 | hopital | National Reference Center for Viruses of Respiratory Infections, Institut Pasteur, Paris | Marion Barbet et al |

| | | | |
|---|---|---|---|
| EPI_ISL_846649,EPI_ISL_872255,EPI_ISL_872276,EPI_ISL_935562,EPI_ISL_953961,EPI_ISL_954018,EPI_ISL_955075,EPI_ISL_955096,EPI_ISL_955100,EPI_ISL_995482,EPI_ISL_1001404,EPI_ISL_1060394,EPI_ISL_1096298,EPI_ISL_1110983,EPI_ISL_1110989,EPI_ISL_1110990,EPI_ISL_1110992,EPI_ISL_1110993,EPI_ISL_1110995,EPI_ISL_1110999,EPI_ISL_1111000,EPI_ISL_1111001,EPI_ISL_1111003,EPI_ISL_1111020,EPI_ISL_1111021,EPI_ISL_1111024,EPI_ISL_1111026,EPI_ISL_1111027,EPI_ISL_1111061,EPI_ISL_1118892,EPI_ISL_1118893,EPI_ISL_1239370,EPI_ISL_1259297,EPI_ISL_1259307,EPI_ISL_1301934,EPI_ISL_1168193,EPI_ISL_1168194,EPI_ISL_1259296 | Hopital | National Reference Center for Viruses of Respiratory Infections, Institut Pasteur, Paris | Marion Barbet et al |
| EPI_ISL_560589 | Hopital | National Reference Center for Viruses of Respiratory Infections, Institut Pasteur, Paris | Sylvie Behillil et al |
| EPI_ISL_560569 | hôpital | National Reference Center for Viruses of Respiratory Infections, Institut Pasteur, Paris | Sylvie Behillil et al |
| EPI_ISL_833198 | Hôpital Bichat Claude Bernard, Laboratoire de Virologie | IAME UMR1137 Inserm, Université de Paris, Hôpital Bichat | Antoine Bridier et al |
| EPI_ISL_940171,EPI_ISL_940190,EPI_ISL_940255,EPI_ISL_940543,EPI_ISL_1296407 | Hôpital Bichat Claude Bernard, Laboratoire de Virologie | IAME UMR1137 Inserm, Université de Paris, Hôpital Bichat | Antoine Bridier-Nahmias et al |
| EPI_ISL_710534,EPI_ISL_712060 | Hôpital Fattouma-Bourguiba de Monastir | Laboratoire des Procédés de Criblage Moléculaire et Cellulaire-Centre de Biotechnologie de Sfax | Souissi et al |
| EPI_ISL_1164947 | Hôpital Georges L. Dumont | National Microbiology Laboratory (NML) | Anna Majer et al |
| EPI_ISL_912734,EPI_ISL_982182,EPI_ISL_982183 | Hôpital Henri Mondor | Department of Virology, Henri Mondor University Hospital, Assistance Publique Hôpitaux de Paris, Université Paris-Est Créteil, INSERM U955 | Christophe Rodriguez et al |
| EPI_ISL_1200857,EPI_ISL_1201852 | HOPITAL SAINT ANDRE | CNR Virus des Infections Respiratoires - France SUD | Antonin Bal et al |
| EPI_ISL_1191681,EPI_ISL_1191682 | HOPITAL UNIVERSITAIRE DE FORT DE FRANCE | CNR Virus des Infections Respiratoires - France SUD | Antonin Bal et al |
| EPI_ISL_718236,EPI_ISL_718241,EPI_ISL_718244,EPI_ISL_855355,EPI_ISL_855382 | Hospital | National Reference Center for Viruses of Respiratory Infections, Institut Pasteur, Paris | Marion Barbet et al |
| EPI_ISL_560603,EPI_ISL_560612,EPI_ISL_560617,EPI_ISL_560618,EPI_ISL_560646 | Hospital | National Reference Center for Viruses of Respiratory Infections, Institut Pasteur, Paris | Sylvie Behillil et al |
| EPI_ISL_682239 | HOSPITAL CIUDAD NEILY | Inciensa, Instituto Costarricense de Investigación y Enseñanza en Nutrición y Salud | Francisco Duarte et al |
| EPI_ISL_541038 | Hospital Clínico Universitario de Santiago de Compostela | SeqCOVID-SPAIN consortium/Institute of Biomedicine of Valencia, IBV-CSIC | José Javier Costa Alcalde et al |
| EPI_ISL_672726 | Hospital das Clínicas da Faculdade de Medicina da Universidade de São Paulo (HC-FMUSP) | Laboratório de Parasitologia Médica - Instituto de Medicina Tropical - Universidade de São Paulo | Brazil-UK Centre for Arbovirus Discovery Diagnosis Genomics et al |
| EPI_ISL_722062,EPI_ISL_722069 | Hospital das Clínicas Universidade de São Paulo Medical School | Laboratório de Parasitologia Médica - Instituto de Medicina Tropical - Universidade de São Paulo | Brazil-UK Centre for Arbovirus Discovery Diagnosis Genomics et al |
| EPI_ISL_511578 | Hospital de Braga | Instituto Nacional de Saude (INSA) | Borges et al et al |

| | | | |
|---|---|---|---|
| EPI_ISL_914816 | HOSPITAL DE NIÑOS DR. CARLOS SAENZ HERRERA | Inciensa, Instituto Costarricense de Investigación y Enseñanza en Nutrición y Salud | Francisco Duarte et al |
| EPI_ISL_512672,EPI_ISL_512674,EPI_ISL_527750 | Hospital De Niños Dr. Carlos Saenz Herrera [San Jose/San Jose] | Inciensa, Instituto Costarricense de Investigación y Enseñanza en Nutrición y Salud | Francisco Duarte et al |
| EPI_ISL_1196433 | HOSPITAL DR. ENRIQUE BALTODANO BRICEÑO | Inciensa, Instituto Costarricense de Investigación y Enseñanza en Nutrición y Salud | Francisco Duarte et al |
| EPI_ISL_914795 | HOSPITAL DR. FERNANDO ESCALANTE PRADILLA | Inciensa, Instituto Costarricense de Investigación y Enseñanza en Nutrición y Salud | Francisco Duarte et al |
| EPI_ISL_1196430 | HOSPITAL DR. MAX PERALTA JIMENEZ | Inciensa, Instituto Costarricense de Investigación y Enseñanza en Nutrición y Salud | Francisco Duarte et al |
| EPI_ISL_770028 | Hospital Dr. Raul Blanco Cervantes | Inciensa, Instituto Costarricense de Investigación y Enseñanza en Nutrición y Salud | Francisco Duarte et al |
| EPI_ISL_491447 | Hospital Fernando Escalante Pradilla | Inciensa, Instituto Costarricense de Investigación y Enseñanza en Nutrición y Salud | Francisco Duarte et al |
| EPI_ISL_458028 | Hospital for Tropical Diseases | COVID-19 Network Investigations (CONI) Alliance | Elizabeth Batty et al |
| EPI_ISL_424731 | Hospital General Regional No.66, Ciudad Juárez, Chihuahua. | Laboratorio Central de Epidemiología-DLVIE / Laboratorio de Secuenciación-Centro de Instrumentos. Instituto Mexicano del Seguro Social | Muñoz-Medina JE et al |
| EPI_ISL_814032,EPI_ISL_814033 | Hospital General Universitario Gregorio Marañón | SeqCOVID-SPAIN consortium/IBV(CSIC) | Darío García de Viedma et al |
| EPI_ISL_481246,EPI_ISL_481247,EPI_ISL_481248,EPI_ISL_574431 | Hospital IESS Babahoyo | Institute of Microbiology, Universidad San Francisco de Quito | Belén Prado-Vivar et al |
| EPI_ISL_412964 | Hospital Israelita Albert Einstein | Instituto Adolfo Lutz Interdisciplinary Procedures Center Strategic Laboratory | Jaqueline Goes de Jesus et al |
| EPI_ISL_413016 | Hospital Israelita Albert Einstein | Instituto Adolfo Lutz, Interdisciplinary Procedures Center, Strategic Laboratory | Jaqueline Goes de Jesus et al |
| EPI_ISL_1196660 | Hospital Melaka | Institute for Medical Research, Infectious Disease Research Centre, National Institutes of Health, Ministry of Health Malaysia | Suppiah J et al |
| EPI_ISL_527739 | Hospital Mexico [San Jose/San Jose] | Inciensa, Instituto Costarricense de Investigación y Enseñanza en Nutrición y Salud | Francisco Duarte et al |
| EPI_ISL_480326,EPI_ISL_480327 | Hospital Nacional de Niños | Charité Virology-University of Costa Rica | Andres Moreira-Soto et al |
| EPI_ISL_471271,EPI_ISL_471270,EPI_ISL_471269 | Hospital Oncológico Solca Núcleo de Quito | Institute of Microbiology, Universidad San Francisco de Quito | Sully Márquez et al |
| EPI_ISL_1186114 | Hospital Queen Elizabeth | Institute for Medical Research, Infectious Disease Research Centre, National Institutes of Health, Ministry of Health Malaysia | Suppiah J et al |
| EPI_ISL_936489 | Hospital Raja Perempuan Zainab II | Institute for Medical Research, Infectious Disease Research Centre, National Institutes of Health, Ministry of Health Malaysia | Suppiah J et al |
| EPI_ISL_682262,EPI_ISL_682273 | HOSPITAL SAN JUAN DE DIOS | Inciensa, Instituto Costarricense de Investigación y Enseñanza en Nutrición y Salud | Francisco Duarte et al |
| EPI_ISL_414015,EPI_ISL_414016 | Hospital São Joaquim Beneficencia Portuguesa | Instituto Adolfo Lutz, Interdiciplinary Procedures Center, Strategic Laboratory | Claudio Tavares Sacchi et al |
| EPI_ISL_902747 | Hospital Universitari Germans Trias i Pujol (HUGTiP) / Fundació Lluita contra la SIDA (FLSida) | IrsiCaixa - Can Ruti CovidSeq | Fundació irsiCaixa. Hospital Universitari Germans Trias i Pujol(HUGTiP) et al |
| EPI_ISL_1017707,EPI_ISL_1017708,EPI_ISL_1017710 | Hospital Universitario Hernando Moncaleano Perdomo | Instituto Nacional de Salud- Dirección de Investigación en Salud Pública | Katherine Laiton-Donato et al |
| EPI_ISL_693536,EPI_ISL_693537 | Hospital Vila Franca de Xira | Instituto Nacional de Saude (INSA) | Borges et al et al |
| EPI_ISL_434771,EPI_ISL_787608,EPI_ISL_790183,EPI_ISL_790302 | Houston Methodist Hospital | Houston Methodist Hospital | S. Wesley Long et al |
| EPI_ISL_636975,EPI_ISL_636977,EPI_ISL_636978 | HP Pemba | KRISP, KZN Research Innovation and Sequencing Platform | Ismael N et al |
| EPI_ISL_445088 | Human Genetic Research Center, Kawsar Biotech Company | Human Genetic Research Center, Kawsar Biotech Company | Abbasalipour Bashash et al |
| EPI_ISL_645080 | Human Genome Variation Research Group, Malopolska Centre of Biotechnology | Human Genome Variation Research Group, Malopolska Centre of Biotechnology | Kowalski et al |
| EPI_ISL_526225 | Hungarian Defence Forces Military Medical Centre | National Laboratory of Virology, Szentágothai Research Centre | Endre Gábor Tóth et al |
| EPI_ISL_759956 | Husada Utama Hospital | Institute of Tropical Disease, Universitas Airlangga | Jezzy R Dewantari et al |

| | | | |
|---|---|---|---|
| EPI_ISL_1219022 | IAL Regional de Sorocaba | Instituto Adolfo Lutz, Interdiciplinary Procedures Center, Strategic Laboratory | Claudio Tavares Sacchi et al |
| EPI_ISL_404253 | IL Department of Public Health Chicago Laboratory | Pathogen Discovery, Respiratory Viruses Branch, Division of Viral Diseases, Centers for Disease Control and Prevention | Ying Tao et al |
| EPI_ISL_961988 | Illinois Department of Public Health | Gagnon Lab, Southern Illinois University | Keith Gagnon et al |
| EPI_ISL_1063818,EPI_ISL_1063886,EPI_ISL_1063895,EPI_ISL_1063896 | Immunogenomics lab, Institute of Life Sciences, Bhubaneswar | Immunogenomics lab, Institute of Life Sciences, Bhubaneswar | Sunil K. Raghav et al |
| EPI_ISL_1018099 | Immunology, Noguchi Memorial Institute for Medical Research | Immunology, Noguchi Memorial Institute for Medical Research | Adu et al |
| EPI_ISL_648212,EPI_ISL_648677 | INBIRS-UBA | Laboratorio Mixto de Biotecnología Acuática (LMBA) | Joaquín Ezpeleta et al |
| EPI_ISL_476704,EPI_ISL_476702 | Incubadora Venezolana de Ciencia, Venezuela | Incubadora Venezolana de Ciencia, Venezuela / Instituto Nacional de Salud, Bogotá, Colombia / Grupo de Investigaciones Microbiológicas-UR (GIMUR), Departamento de Biología, Facultad de Ciencias Naturales, Universidad del Rosario, Bogotá, Colombia / Icahn School of Medicine at Mount Sinai, New York, USA | Alberto Paniz-Mondolfi et al |
| EPI_ISL_413522 | Indian Council of Medical Research - National Institute of Virology | National Influenza Center, Indian Council of Medical Research - National Institute of Virology | Potdar V et al |
| EPI_ISL_413523 | Indian Council of Medical Research-National Institute of Virology | National Influenza Center, Indian Council of Medical Research-National Institute of Virology | Potdar V et al |
| EPI_ISL_496372 | Infectolab | Andersen lab at Scripps Research | SEARCH Alliance San Diego with Samuel Navarro Alvarez et al |
| EPI_ISL_677895,EPI_ISL_677902 | Innovative Genomics Institute, UC Berkeley | Innovative Genomics Institute, UC Berkeley | Stacia Wyman et al |
| EPI_ISL_806544 | INSPI Instituto Nacional de Investigación en Salud Pública | Av. Julián Coronel 905 entre Esmeraldas y José Mascote Av. Juan Tanca Marengo No. 100 y Av. de las Américas | Leandro Patiño et al |
| EPI_ISL_826829 | INSPI-CRN DE INFLUENZA Y OTROS VIRUS RESPIRATORIOS | Instituto de Salud Publica de Chile | Javier Tognarelli et al |
| EPI_ISL_811130 | Institut Central des Hôpitaux Valaisans ICHV/ZIWS Service des Maladies Infectieuses | Department of Biosystems Science and Engineering, ETH Zürich | Chaoran Chen et al |
| EPI_ISL_437932,EPI_ISL_583600 | Institut für Virologie am Department für Hygiene, Mikrobiologie und Public Health | Bergthaler laboratory, CeMM Research Center for Molecular Medicine of the Austrian Academy of Sciences | Alexandra Popa et al |
| EPI_ISL_934544,EPI_ISL_934545 | Institut für Virologie am Department für Hygiene, Mikrobiologie und Public Health | Bergthaler laboratory, CeMM Research Center for Molecular Medicine of the Austrian Academy of Sciences | Lukas Endler et al |
| EPI_ISL_1141634 | Institut für Virologie des Universitätsklinikums Tübingen | Robert Koch Institute | et al |
| EPI_ISL_1293351 | Institut National d'Hygiène (INH) | Unité Mixte Internationale TransVIHMI (UMI 233 IRD – U1175 INSERM - Université de Montpellier)IRD (Institut de recherche pour le développement) | Mounerou SALOU et al |
| EPI_ISL_418208,EPI_ISL_420072,EPI_ISL_480787,EPI_ISL_482878,EPI_ISL_485710,EPI_ISL_486861,EPI_ISL_486862,EPI_ISL_486859,EPI_ISL_486872,EPI_ISL_486867 | Institut Pasteur Dakar | Institut Pasteur de Dakar | Ndongo Dia et al |
| EPI_ISL_498232,EPI_ISL_498236,EPI_ISL_498238,EPI_ISL_498239,EPI_ISL_498251,EPI_ISL_498252 | Institut Pasteur de Dakar | Institut Pasteur de Dakar | Ndongo Dia et al |
| EPI_ISL_1013214,EPI_ISL_1013215,EPI_ISL_1013216,EPI_ISL_1013417,EPI_ISL_1013421,EPI_ISL_1013432 | Institut Pasteur de Guadeloupe | National Reference Center for Viruses of Respiratory Infections, Institut Pasteur, Paris | Marion Barbet et al |
| EPI_ISL_999032 | Institut Pasteur de Guinée | Institut Pasteur de Dakar | Grayo Solene et al |

| | | | |
|---|---|---|---|
| EPI_ISL_613455,EPI_ISL_613456,EPI_ISL_613453,EPI_ISL_613454,EPI_ISL_613451,EPI_ISL_613452,EPI_ISL_613429,EPI_ISL_613442,EPI_ISL_613448,EPI_ISL_613449,EPI_ISL_613446,EPI_ISL_613440 | Institut Pasteur de la Guadeloupe | Institut Pasteur de la Guadeloupe | Marion Barbet et al |
| EPI_ISL_459965,EPI_ISL_459966,EPI_ISL_459967,EPI_ISL_459973,EPI_ISL_459977,EPI_ISL_459983,EPI_ISL_459984 | Institut Pasteur du Maroc | Institut Pasteur du Maroc | Marion Barbet et al |
| EPI_ISL_1020296 | Institute for Hygiene and Microbiology | Center for Virology | Jeremy V. Camp et al |
| EPI_ISL_853784,EPI_ISL_853915 | Institute for Laboratory Diagnostics and Microbiology, Klinikum Klagenfurt am Worthersee | Bergthaler laboratory, CeMM Research Center for Molecular Medicine of the Austrian Academy of Sciences | Lukas Endler et al |
| EPI_ISL_933804 | Institute for Lung Diseases in Children - Skopje | Research Center for Genetic Engineering and Biotechnology Georgi D. Efremov" , Macedonian Academy of Sciences and Arts" | RCGEB - MASA et al |
| EPI_ISL_490089,EPI_ISL_490101,EPI_ISL_490102,EPI_ISL_596449,EPI_ISL_718267,EPI_ISL_718271,EPI_ISL_718269,EPI_ISL_718272,EPI_ISL_718277,EPI_ISL_718278,EPI_ISL_718281,EPI_ISL_718280,EPI_ISL_718282,EPI_ISL_718284,EPI_ISL_877228,EPI_ISL_968089,EPI_ISL_1055262,EPI_ISL_1114721,EPI_ISL_1114722,EPI_ISL_1196866,EPI_ISL_1263460 | Institute for Medical Research, Infectious Disease Research Centre, National Institutes of Health, Ministry of Health Malaysia | Institute for Medical Research, Infectious Disease Research Centre, National Institutes of Health, Ministry of Health Malaysia | Suppiah J et al |
| EPI_ISL_430442,EPI_ISL_430441 | Institute for Medical Research, Infectious Disease Research Centre, National Institutes of Health, Ministry of Health Malaysia | Institute for Medical Research, Infectious Disease Research Centre, National Institutes of Health, Ministry of Health Malaysia | Suppiah.J et al |
| EPI_ISL_455790,EPI_ISL_455791,EPI_ISL_455792 | Institute for Medical Research, Infectious Disease Research Centre, National Institutes of Health, Ministry of Health Malaysia | Malaysia Genome Institute | Mohd Noor Mat Isa et al |
| EPI_ISL_728561,EPI_ISL_733552,EPI_ISL_812954 | Institute for Urban Disease Control and Prevention | COVID-19 Network Investigations (CONI) Alliance | Kamolthip Atsawawaranunt et al |
| EPI_ISL_891270,EPI_ISL_891272 | Institute of Biocides and Medical Ecology, Belgarde, Serbia | Virology department Institute of microbiology and immunology Faculty of Medicine University of Belgrade | Banko Ana et al |
| EPI_ISL_1057036,EPI_ISL_1057038 | Institute of Biocides and Medical Ecology, Belgarde, Serbia | Virology Department Institute of Microbiology and Immunology Faculty of Medicine University of Belgrade | Banko Ana et al |
| EPI_ISL_600444,EPI_ISL_600476,EPI_ISL_600484,EPI_ISL_600512,EPI_ISL_600562 | Institute of Epidemiology Disease Control And Research | Institute for Developing Science and Health Initiatives | Lauren Cowley et al |
| EPI_ISL_877453,EPI_ISL_979251 | Institute of Microbiology and Immunology, Faculty of Medicine, University of Ljubljana | Institute of Microbiology and Immunology, Faculty of Medicine, University of Ljubljana | Samo Zakotnik et al |
| EPI_ISL_635279 | Institute of Microbiology and Immunology, Faculty of Medicine, University of Ljubljana | Institute of Microbiology and Immunology, Faculty of Medicine, University of Ljubljana | Tomaž Mark Zorec et al |
| EPI_ISL_697787 | Institute of Microbiology, Universidad San Francisco de Quito | Institute of Microbiology, Universidad San Francisco de Quito | Andrea Macias et al |
| EPI_ISL_486844,EPI_ISL_491932,EPI_ISL_527809,EPI_ISL_527815,EPI_ISL_697785,EPI_ISL_697788,EPI_ISL_697799,EPI_ISL_697800,EPI_ISL_728205,EPI_ISL_824293 | Institute of Microbiology, Universidad San Francisco de Quito | Institute of Microbiology, Universidad San Francisco de Quito | Belén Prado-Vivar et al |
| EPI_ISL_660529,EPI_ISL_660530,EPI_ISL_660537,EPI_ISL_728202 | Institute of Microbiology, Universidad San Francisco de Quito | Institute of Microbiology, Universidad San Francisco de Quito | Sully Márquez et al |
| EPI_ISL_548944,EPI_ISL_548945,EPI_ISL_548942,EPI_ISL_548943,EPI_ISL_548946 | Institute of Microbiology, University of Veterinary and Animal sciences | Institute of Microbiology, University of Veterinary and Animal sciences | Yaqub et al |

| | | | |
|---|---|---|---|
| EPI_ISL_1233843,EPI_ISL_1233855 | Institute of Molecular and Translational Medicine / Laboratory of Experimental Medicine, Faculty of Medicine and Dentistry, Palacky University | Institute of Molecular and Translational Medicine / Laboratory of Experimental Medicine | Rastislav Slavkovský et al |
| EPI_ISL_577740,EPI_ISL_577742,EPI_ISL_583487,EPI_ISL_875518 | Institute of Virology, Biomedical Research Center of the Slovak Academy of Sciences, Bratislava | Faculty of Natural Sciences, Comenius University, Bratislava | Broňa Brejová et al |
| EPI_ISL_577739,EPI_ISL_959621 | Institute of Virology, Biomedical Research Center of the Slovak Academy of Sciences, Bratislava | Faculty of Natural Sciences, Comenius University, Bratislava | Kristína Boršová et al |
| EPI_ISL_779655,EPI_ISL_1234472 | Institute of Virology, Biomedical Research Center of the Slovak Academy of Sciences, Bratislava | Faculty of Natural Sciences, Comenius University, Bratislava | Viktória Čabanová et al |
| EPI_ISL_577734,EPI_ISL_583481 | Institute of Virology, Biomedical Research Center of the Slovak Academy of Sciences, Bratislava | Faculty of Natural Sciences, Comenius University, Bratislava | Viktória Hodorová et al |
| EPI_ISL_417877 | Institute of Virology, Biomedical Research Center of the Slovak Academy of Sciences, Bratislava; Public Health Authority of the Slovak Republic, Bratislava | Institute of Virology, Biomedical Research Center of the Slovak Academy of Sciences, Bratislava; Comenius University Science Park, Bratislava | Monika Slávikova et al |
| EPI_ISL_1017679,EPI_ISL_1017687,EPI_ISL_1017691 | Institute of Virology, Vaccines and Sera "Torlak" | Institute of microbiology and Immunology, Faculty of Medicine, University of Belgrade | Knezevic et al |
| EPI_ISL_658901 | Instituto de Diagnostico y Referencia Epidemiologicos (INDRE) | Instituto de Diagnostico y Referencia Epidemiologicos (INDRE) | Ernesto Ramirez-Gonzalez et al |
| EPI_ISL_493337,EPI_ISL_516618,EPI_ISL_516619 | Instituto de Diagnostico y Referencia Epidemiologicos (INDRE) | Instituto de Diagnostico y Referencia Epidemiologicos (INDRE) | Gisela Barrera-Badillo et al |
| EPI_ISL_455455 | Instituto de Diagnostico y Referencia Epidemiologicos (INDRE) | Instituto de Diagnostico y Referencia Epidemiologicos (INDRE) | Rodriguez-Maldonado Abril et al |
| EPI_ISL_452139,EPI_ISL_452141 | Instituto de Diagnostico y Referencia Epidemiologicos (INDRE) | Instituto de diagnóstico y Referencia Epidemiologicos (INDRE) | Ramirez-Gonzalez Ernesto et al |
| EPI_ISL_1060676,EPI_ISL_1060681,EPI_ISL_1060684,EPI_ISL_1060699,EPI_ISL_1060729 | Instituto de Diagnostico y Referencia Epidemiologicos (INDRE)_RNLSP | Instituto de Diagnostico y Referencia Epidemiologicos (INDRE) | Claudia Wong-Arambula et al |
| EPI_ISL_1301455,EPI_ISL_1301548,EPI_ISL_1301568,EPI_ISL_1301691 | Instituto de Diagnostico y Referencia Epidemiologicos InDRE_RNLSP | Instituto de Biotecnología de la UNAM | Authors from IBT et al |
| EPI_ISL_913945,EPI_ISL_913947,EPI_ISL_913967,EPI_ISL_913968,EPI_ISL_1054955,EPI_ISL_1054960,EPI_ISL_1168605,EPI_ISL_1168608,EPI_ISL_1168609,EPI_ISL_1219715 | Instituto de Diagnostico y Referencia Epidemiologicos INDRE_RNLSP | Instituto de Diagnostico y Referencia Epidemiologicos (INDRE) | Claudia Wong-Arambula et al |
| EPI_ISL_491265 | Instituto Gulbenkian de Ciência | Instituto Gulbenkian de Ciência | Susana Ladeiro et al |
| EPI_ISL_412972 | Instituto Nacional de Enfermedades Respiratorias | Instituto de Diagnostico y Referencia Epidemiologicos (INDRE) | Ramirez-Gonzalez Ernesto et al |
| EPI_ISL_1301453,EPI_ISL_1302180,EPI_ISL_1302205,EPI_ISL_1302250,EPI_ISL_1302263,EPI_ISL_1302269,EPI_ISL_1302292,EPI_ISL_1302298,EPI_ISL_1302345,EPI_ISL_1302381 | Instituto Nacional de Enfermedades Respiratorias (INER) | Instituto de Biotecnología de la UNAM | Authors from IBT et al |
| EPI_ISL_837610,EPI_ISL_837613,EPI_ISL_837734 | Instituto Nacional de Enfermedades Respiratorias (INER) | Instituto Nacional de Enfermedades Respiratorias (INER) | Celia Boukadida et al |
| EPI_ISL_574294 | Instituto Nacional de Investigacion en Salud Pública | Instituto Nacional de Investigación en Salud Pública | Leandro Patino Patino et al |
| EPI_ISL_574293 | Instituto Nacional de Investigación en Salud Pública | Instituto Nacional de Investigación en Salud Publica | Leandro Patino Patino et al |
| EPI_ISL_491944,EPI_ISL_491945,EPI_ISL_491946,EPI_ISL_491947,EPI_ISL_491950 | Instituto Nacional de Investigación en Salud Pública - INSPI | INSPI - Charité | Alfredo Bruno Caicedo et al |

| | | | |
|---|---|---|---|
| EPI_ISL_933515,EPI_ISL_955190,EPI_ISL_1005569,EPI_ISL_1181706,EPI_ISL_1205214,EPI_ISL_1262637,EPI_ISL_1262646,EPI_ISL_1262647,EPI_ISL_1262652,EPI_ISL_1262657,EPI_ISL_1262667,EPI_ISL_1262668,EPI_ISL_1262673,EPI_ISL_1262674 | Instituto Nacional de Medicina Genomica | Instituto Nacional de Medicina Genomica | Hidalgo-Miranda A et al |
| EPI_ISL_418262 | Instituto Nacional de Salud | Instituto Nacional de Salud Universidad Cooperativa de Colombia Instituto Alexander von Humboldt Imperial College-London London School of Hygiene & Tropical Medicine | Marcela Mercado-Reyes et al |
| EPI_ISL_536477,EPI_ISL_536485,EPI_ISL_536492,EPI_ISL_536552 | Instituto Nacional de Salud | Laboratorio de Infecciones Respiratorias Agudas | Eduardo Juscamayta Lopez et al |
| EPI_ISL_456139,EPI_ISL_456142,EPI_ISL_456156 | Instituto Nacional de Salud - Unidad de Secuenciación y Análisis Genómico | Instituto Nacional de Salud, Universidad Cooperativa de Colombia, Instituto Alexander von Humboldt, Imperial College-London, London School of Hygiene & Tropical Medicine | Katherine Laiton-Donato et al |
| EPI_ISL_941955,EPI_ISL_941986 | Instituto Nacional de Salud, Bogotá, Colombia | Centro de Investigaciones en Microbiología y Biotecnología-UR (CIMBIUR), Facultad de Ciencias Naturales, Universidad del Rosario, Bogotá, Colombia Instituto Nacional de Salud, Bogotá, Colombia Icahn School of Medicine at Mount Sinai, New York, USA | Luz Helena Patiño et al |
| EPI_ISL_526932,EPI_ISL_526956,EPI_ISL_526971,EPI_ISL_526969,EPI_ISL_526967,EPI_ISL_653746,EPI_ISL_653754,EPI_ISL_653758,EPI_ISL_653759,EPI_ISL_653760,EPI_ISL_653762,EPI_ISL_739663,EPI_ISL_739672,EPI_ISL_739674,EPI_ISL_739675,EPI_ISL_739677 | Instituto Nacional de Salud, Bogotá, Colombia | Instituto Nacional de Salud, Bogotá, Colombia | Katherine Laiton-Donato et al |
| EPI_ISL_887420,EPI_ISL_887421,EPI_ISL_887424,EPI_ISL_887426,EPI_ISL_887429,EPI_ISL_887430,EPI_ISL_887434,EPI_ISL_887467,EPI_ISL_887473,EPI_ISL_887481,EPI_ISL_887490,EPI_ISL_887500,EPI_ISL_887503,EPI_ISL_964918,EPI_ISL_1132837,EPI_ISL_1132838 | Instituto Nacional de Saude (INS), Mozambique | KRISP, KZN Research Innovation and Sequencing Platform | Nalia Ismael et al |
| EPI_ISL_511044,EPI_ISL_693628,EPI_ISL_861567,EPI_ISL_1023463,EPI_ISL_1170140,EPI_ISL_1260855 | Instituto Nacional de Saude (INSA) | Instituto Nacional de Saude (INSA) | Borges et al et al |
| EPI_ISL_941739 | Instituto Nacional de Saude (INSA) and Instituto Gulbenkian de Ciencia (IGC) | Instituto Nacional de Saude (INSA) and Instituto Gulbenkian de Ciencia (IGC) | Borges et al et al |
| EPI_ISL_1167134,EPI_ISL_1167135,EPI_ISL_1167138,EPI_ISL_1167145,EPI_ISL_1167148,EPI_ISL_1167160,EPI_ISL_1167161,EPI_ISL_1167164,EPI_ISL_1167171,EPI_ISL_1167181 | Iressef Genomics lab | L'institut de Recherche en Santé, de Surveillance Épidémiologique et de Formation (IRESSEF) | Souleymane MBOUP et al |
| EPI_ISL_956386,EPI_ISL_956387 | Isolation - Virology Unit, Institut Pasteur du Cambodge; Sequencing - US National Institute of Allergy and Infectious Diseases Cambodia, US Naval Medical Research Unit -2, Cambodia National Institute for Public Health | Virology Unit, Institut Pasteur du Cambodge | Vireak Heang et al |

| | | | |
|---|---|---|---|
| EPI_ISL_474959,EPI_ISL_474961,EPI_ISL_474977,EPI_ISL_475017,EPI_ISL_514301,EPI_ISL_514302,EPI_ISL_516889,EPI_ISL_516895,EPI_ISL_516897,EPI_ISL_516899,EPI_ISL_516905,EPI_ISL_516913,EPI_ISL_516920,EPI_ISL_575333,EPI_ISL_575332,EPI_ISL_649072,EPI_ISL_649084,EPI_ISL_649094 | Israel Central Virology laboratory | Israel Central Virology laboratory | Neta Zuckerman et al |
| EPI_ISL_804057,EPI_ISL_889144,EPI_ISL_944237,EPI_ISL_944240,EPI_ISL_1073390,EPI_ISL_1073394,EPI_ISL_1209487,EPI_ISL_1240651 | Israel Central Virology laboratory | Israel National Consortium for SARS-CoV-2 sequencing | Neta Zuckerman et al |
| EPI_ISL_1240647,EPI_ISL_1240648 | Israel Central Virology Laboratory | Israel National Consortium for SARS-CoV-2 sequencing | Neta Zuckerman et al |
| EPI_ISL_594159,EPI_ISL_594160 | Israel Institute for Biological Research | Israel Institute for Biological Research | Galia Zaide et al |
| EPI_ISL_778697 | Istituto Zooprofilattico Sperimentale del Mezzogiorno | TIGEM | Antonio Grimaldi et al |
| EPI_ISL_525566 | Istituto Zooprofilattico Sperimentale Puglia e Basilicata; Dipartimento di Bioscienze, Biotecnologie e Biofarmaceutica dell'Università degli Studi di Bari "A.Moro"; Istituto di Biomembrane. Bioenergetica e Biotecnologie Molecolari del Consiglio Nazionale delle Ricerche di Bari | Beaconlab (Bioinformatics, Evolution and Comparative Genomics lab), Dept of Biosciences, University on Milan | Parisi A. et al |
| EPI_ISL_1086090 | IZSM | TIGEM | Antonio Grimaldi et al |
| EPI_ISL_422424 | Jaber Al Ahmad Al Sabah Hospital | Dasman diabetes Institute | Fahd Al-Mulla et al |
| EPI_ISL_422426,EPI_ISL_422427 | JABER AL AHMAD AL SABAH HOSPITAL – KUWAIT CITY | Dasman Diabetes Institute | Fahd Al-Mulla et al |
| EPI_ISL_450797,EPI_ISL_450799 | Jamaica Ministry of Health and Wellness | Pathogen Discovery, Respiratory Viruses Branch, Division of Viral Diseases, Centers for Disease Control and Prevention | Yan Li et al |
| EPI_ISL_779290,EPI_ISL_779255,EPI_ISL_779277,EPI_ISL_779274,EPI_ISL_779273,EPI_ISL_779278,EPI_ISL_779279,EPI_ISL_779286,EPI_ISL_779283,EPI_ISL_779261,EPI_ISL_779284,EPI_ISL_779269,EPI_ISL_779289 | Jamil-ur-Rahman Center for Genome Research, Dr. Panjwani Center for Molecular Medicine and Drug Research | Jamil-ur-Rahman Center for Genome Research, Dr. Panjwani Center for Molecular Medicine and Drug Research | Shakeel et al |
| EPI_ISL_451958 | Jamil-ur-Rahman Center for Genome Research, Dr. Panjwani Center for Molecular Medicine and Drug Research, International Center for Chemical and Biological Sciences, University of Karachi | Jamil-ur-Rahman Center for Genome Research, Dr. Panjwani Center for Molecular Medicine and Drug Research, International Center for Chemical and Biological Sciences, University of Karachi | Shakeel et al |
| EPI_ISL_416623,EPI_ISL_416618 | Japanese Quarantine Stations | Pathogen Genomics Center, National Institute of Infectious Diseases | Tsuyoshi Sekizuka et al |
| EPI_ISL_495435 | Kafkas University, Faculty of Medicine, Department of Medical Microbiology | Kafkas University, Faculty of Medicine, Department of Medical Microbiology | Murat Karamese et al |
| EPI_ISL_534235 | Karolinska universitetslaboratoriet SOLNA | The Public Health Agency of Sweden | Anna-Malin Linde et al |
| EPI_ISL_457855,EPI_ISL_457890 | KEMRI-CGMR-C | KEMRI-Wellcome Trust Research Programme/KEMRI-CGMR-C Kilifi | Githinji G. et al 2020 et al |
| EPI_ISL_568699,EPI_ISL_568710,EPI_ISL_568734,EPI_ISL_568813,EPI_ISL_568819,EPI_ISL_568860,EPI_ISL_568866,EPI_ISL_568867 | KEMRI-Wellcome Trust Research Programme/KEMRI-CGMR-C Kilifi | KEMRI-Wellcome Trust Research Programme/KEMRI-CGMR-C Kilifi | Githinji et al 2020 et al |

| | | | |
|---|---|---|---|
| EPI_ISL_806570,EPI_ISL_806575,EPI_ISL_806611,EPI_ISL_806617,EPI_ISL_806623,EPI_ISL_806624,EPI_ISL_806662,EPI_ISL_806663,EPI_ISL_806670,EPI_ISL_806681,EPI_ISL_806703,EPI_ISL_806704,EPI_ISL_806705,EPI_ISL_806706,EPI_ISL_806710,EPI_ISL_806711,EPI_ISL_806712,EPI_ISL_855535,EPI_ISL_855534,EPI_ISL_969074,EPI_ISL_969001,EPI_ISL_969004,EPI_ISL_969027,EPI_ISL_968858,EPI_ISL_968986,EPI_ISL_968869,EPI_ISL_968873,EPI_ISL_968910,EPI_ISL_968850,EPI_ISL_968810,EPI_ISL_1039228,EPI_ISL_1039229 | KEMRI-Wellcome Trust Research Programme/KEMRI-CGMR-C Kilifi | KEMRI-Wellcome Trust Research Programme/KEMRI-CGMR-C Kilifi | Githinji et al et al |
| EPI_ISL_512811,EPI_ISL_512812,EPI_ISL_512815 | Kenema Government Hospital, Ministry of Health and Sanitation | Kenema Government Hospital, Ministry of Health and Sanitation | Goba et al |
| EPI_ISL_1239480 | Kimberley Hospital, National Health Laboratory Services, Northern Cape, South Africa | National Institute for Communicable Diseases of the National Health Laboratory Service | Amoako DG et al |
| EPI_ISL_490007,EPI_ISL_490008 | King Fahad Medical City | King Fahad Medical City | Alosaimi et al |
| EPI_ISL_516946,EPI_ISL_516967 | King Georges Medical University | CSIR-National Botanical Research Institute | Priti Prasad et al |
| EPI_ISL_458032 | King Institute of Preventive Medicine & Research | CSIR-Centre for Cellular and Molecular Biology | K.Kaveri et al |
| EPI_ISL_483543,EPI_ISL_483545,EPI_ISL_483546,EPI_ISL_483549,EPI_ISL_483553,EPI_ISL_483564 | Kingdom of Bahrein Ministry of Health | Erasmus Medical Center | Bas Oude Munnink et al |
| EPI_ISL_833390 | Klinik Apotek Dein, Jakarta, Indonesia | Biosafety Level-3 Laboratory, Indonesian Institute of Sciences (LIPI) | Andri Wardiana et al |
| EPI_ISL_833389,EPI_ISL_833388 | Klinik Apotek Dein, Jakarta, Indonesia | Biosafety Level-3 Laboratory, Indonesian Institute of Sciences (LIPI) | Syam Budi Iryanto et al |
| EPI_ISL_654946,EPI_ISL_934334 | Klinisk mikrobiologi | The Public Health Agency of Sweden | Anna-Malin Linde et al |
| EPI_ISL_766696 | Klinisk Mikrobiologi | The Public Health Agency of Sweden | Department of Microbiology et al |
| EPI_ISL_984963,EPI_ISL_1091567 | Klinisk mikrobiologi, Region Västerbotten | CBRN Defence and Security, Swedish Defence Research Agency | Andreas Sjödin et al |
| EPI_ISL_534245 | Kliniskt mikrobiologiska laboratoriet | The Public Health Agency of Sweden | Anna-Malin Linde et al |
| EPI_ISL_582774 | Klinsisk mikrobiologi Linkoping | The Public Health Agency of Sweden | Anna-Malin Linde et al |
| EPI_ISL_407193 | Korea Centers for Disease Control & Prevention (KCDC) Center for Laboratory Control of Infectious Diseases Division of Viral Diseases | Korea Centers for Disease Control & Prevention (KCDC) Center for Laboratory Control of Infectious Diseases Division of Viral Diseases | Jeong-Min Kim et al |
| EPI_ISL_436684,EPI_ISL_455639 | KRISP, KZN Research Innovation and Sequencing Platform | KRISP, KZN Research Innovation and Sequencing Platform | Giandhari J et al |
| EPI_ISL_888702,EPI_ISL_888707,EPI_ISL_888716,EPI_ISL_888718,EPI_ISL_888721,EPI_ISL_1049150,EPI_ISL_1049151,EPI_ISL_1049173,EPI_ISL_1048849 | KU Leuven, Rega Institute, Clinical and Epidemiological Virology | KU Leuven, Rega Institute, Clinical and Epidemiological Virology | Tony Wawina-Bokalanga et al |
| EPI_ISL_475569 | Kungsholmsdoktorn | The Public Health Agency of Sweden | Oskar Karlsson Lindsjo et al |
| EPI_ISL_747239 | Kuningan Publlic Health Office | West Java Health Laboratory; School of Life Sciences and Technology, Institut Teknologi Bandung | Azzania Fibriani et al |
| EPI_ISL_982262 | Lab voor klinische biologie | Lab voor klinische biologie | Hannelore Hamerlinck et al |
| EPI_ISL_717616 | Lab voor klinische biologie | Onderzoeksgroep Virologie | Laurens Lambrechts et al |
| EPI_ISL_717618 | Lab voor klinische biologie | Onderzoeksgroep Virologie | Nick Vereecke et al |
| EPI_ISL_1154656 | LabKom - Labor Augsburg MVZ GmbH | Robert Koch Institute | et al |

| | | | |
|---|---|---|---|
| EPI_ISL_1156681 | LabKom - MVZ Labor Bochum MLB GmbH | Robert Koch Institute | et al |
| EPI_ISL_955071,EPI_ISL_955086 | Labo analyses Med | National Reference Center for Viruses of Respiratory Infections, Institut Pasteur, Paris | Marion Barbet et al |
| EPI_ISL_872258,EPI_ISL_872259,EPI_ISL_890353,EPI_ISL_935643,EPI_ISL_954092,EPI_ISL_1060400,EPI_ISL_1111064,EPI_ISL_1201521,EPI_ISL_1233246,EPI_ISL_1259298,EPI_ISL_1262787,EPI_ISL_1262803,EPI_ISL_1263007 | Labo Analyses Med | National Reference Center for Viruses of Respiratory Infections, Institut Pasteur, Paris | Marion Barbet et al |
| EPI_ISL_1146104,EPI_ISL_1154843,EPI_ISL_1211236 | Labor Dr. Schumacher MVZ | Robert Koch Institute | et al |
| EPI_ISL_775226,EPI_ISL_775220,EPI_ISL_775260 | Laboratoire Biolife | Laboratoire de Biotechnologie | Mouna Ouadghiri et al |
| EPI_ISL_1116592,EPI_ISL_1116470,EPI_ISL_1120732 | Laboratoire central de Virologie | Laboratoire de Biotechnologie | Hakima Kabbaj et al |
| EPI_ISL_1103576,EPI_ISL_1137621 | Laboratoire central de Virologie | Laboratoire de Biotechnologie | Myriam Seffar et al |
| EPI_ISL_660446,EPI_ISL_660450,EPI_ISL_660451,EPI_ISL_660452,EPI_ISL_660465,EPI_ISL_660467,EPI_ISL_660471,EPI_ISL_660473,EPI_ISL_660474,EPI_ISL_660477,EPI_ISL_660478,EPI_ISL_660479,EPI_ISL_660493,EPI_ISL_660495,EPI_ISL_660498,EPI_ISL_660503,EPI_ISL_660504,EPI_ISL_660515,EPI_ISL_660516,EPI_ISL_660519,EPI_ISL_660521,EPI_ISL_660522,EPI_ISL_660525 | Laboratoire de Microbiologie CHU Souro Sanou | Centre Muraz | Abdoul-Salam Ouedraogo et al |
| EPI_ISL_1116468,EPI_ISL_1116467,EPI_ISL_1116464,EPI_ISL_1116469,EPI_ISL_1118884 | Laboratoire de Microbiologie- CHU Habib Bourguiba – Sfax | Laboratoire des Procédés de Criblage Moléculaire et Cellulaire-Centre de Biotechnologie de Sfax | Souissi et al |
| EPI_ISL_712062,EPI_ISL_712063 | Laboratoire de Microbiologie- CHU Habib Bourguiba – Sfax adresse | Laboratoire des Procédés de Criblage Moléculaire et Cellulaire-Centre de Biotechnologie de Sfax | Souissi et al |
| EPI_ISL_476026 | Laboratoire de Recherche et d'Analyses Médicales de la Gendarmerie Royale | Laboratoire de Recherche et d'Analyses Médicales de la Gendarmerie Royale | Sanaâ Lemriss et al |
| EPI_ISL_826123,EPI_ISL_826125 | Laboratoire de santé publique du Québec | Laboratoire de santé publique du Québec | Sandrine Moreira et al |
| EPI_ISL_1197037,EPI_ISL_1208399 | Laboratoire de virologie clinique - Institut Pasteur de Tunis | 1-Laboratory of Microbiology, National Reference Lab, Charles Nicolle Hospital; 2-University of Tunis ElManar, Faculty of Medicine of Tunis, LR99ES09, Tunis, Tunisia | Sameh Trabelsi et al |
| EPI_ISL_414600 | Laboratoire de Virologie Institut de Virologie - INSERM U 1109 Hôpitaux Universitaires de Strasbourg | National Reference Center for Viruses of Respiratory Infections, Institut Pasteur, Paris | Mélnie Albert et al |
| EPI_ISL_666870 | Laboratoire de Virologie, CHU de Caen, Normandie, France. | GRAM2.0, Université de Caen Normandie | Meriadeg LE GOUIL et al |
| EPI_ISL_476823,EPI_ISL_476824,EPI_ISL_476825,EPI_ISL_476828,EPI_ISL_476830 | Laboratoire des Fièvres Hémorragiques Virales du Benin | Charité-Universitätsmedizin Berlin | Yadouleton et al |
| EPI_ISL_629082 | Laboratoire du Centre Hospitalier Annecy Genevois | CNR Virus des Infections Respiratoires - France SUD | Antonin Bal et al |
| EPI_ISL_910350,EPI_ISL_910352,EPI_ISL_910445,EPI_ISL_910650,EPI_ISL_910909,EPI_ISL_911130,EPI_ISL_911138,EPI_ISL_911234 | Laboratoire national de sante, Microbiology, Virology | Laboratoire national de sante, Microbiology, Microbial Genomics Platform | Anke Wienecke-Baldacchino et al |
| EPI_ISL_429721 | Laboratoire National de Sante, Microbiology, Virology | Laboratoire National de Sante, Microbiology, Epidemiology and Microbial Genomics | Anke Wienecke-Baldacchino et al |
| EPI_ISL_744295 | Laboratoire national de santé, Microbiology, Virology | Laboratoire national de santé, Microbiology, Epidemiology and Microbial Genomics | Anke Wienecke-Baldacchino et al |

| | | | |
|---|---|---|---|
| EPI_ISL_739733,EPI_ISL_740092,EPI_ISL_740127,EPI_ISL_755589 | Laboratoire national de santé, Microbiology, Virology | Laboratoire national de santé, Microbiology, Microbial Genomics Platform | Anke Wienecke-Baldacchino et al |
| EPI_ISL_1265882 | LABORATOIRE ORIADE | CNR Virus des Infections Respiratoires - France SUD | Antonin Bal et al |
| EPI_ISL_1288162 | Laboratorio Central de Epidemiología (LCE) | Instituto de Biotecnología de la UNAM | Consorcio Mexicano de Vigilancia Genómica (CoViGen-Mex). Authors (in alphabetical order): Julio Elias Alvarado-Yaah et al |
| EPI_ISL_1287776 | Laboratorio Central de Epidemiología (LCE) | Instituto Nacional de Enfermedades Respiratorias (INER): Centro de Investigación en Enfermedades Infecciosas (CIENI) | Consorcio Mexicano de Vigilancia Genómica (CoViGen-Mex). Authors (in alphabetical order): Julio Elias Alvarado-Yaah et al |
| EPI_ISL_792512 | Laboratorio Central de la Ciudad de Santa Fe | Grupo de Genómica y Bioinformática del Instituto de Investigación de la Cadena Láctea CONICET-INTA on behalf of 'Proyecto Argentino Interinstitucional de genomica de SARS-CoV-2' (PAIS Consortium) | Eberhardt et al |
| EPI_ISL_1239121 | Laboratório Central de Saúde Pública do Espírito Santo | Coordenação Geral de Laboratórios de Saúde Pública (CGLAB) | Vagner Fonseca et al et al |
| EPI_ISL_792636 | Laboratório Central de Saúde Pública do Estado da Paraíba (LACEN-PB) | Laboratory of Respiratory Viruses and Measles, Oswaldo Cruz Institute, FIOCRUZ | Paola Resende et al |
| EPI_ISL_415128 | Laboratório Central de Saúde Pública do Estado do Espírito Santo (LACEN-ES) | Laboratory of Respiratory Viruses and Measles, Oswaldo Cruz Institute, FIOCRUZ | Paola Resende et al |
| EPI_ISL_1181353 | Laboratorio Central de Saude Publica do Estado do Rio de Janeiro (LACEN-RJ) | Laboratory of Respiratory Viruses and Measles, Oswaldo Cruz Institute, FIOCRUZ | Paola Resende et al |
| EPI_ISL_792529,EPI_ISL_792530,EPI_ISL_792537 | Laboratorio Central, Ministerio de Salud Córdoba | Instituto de Patologia Vegetal (CIAP-INTA) on behalf of 'Proyecto Argentino Interinstitucional de genomica de SARS-CoV-2' (PAIS Consortium) | Fernández et al |
| EPI_ISL_1196432 | LABORATORIO CLINICO LABIN | Inciensa, Instituto Costarricense de Investigación y Enseñanza en Nutrición y Salud | Francisco Duarte et al |
| EPI_ISL_480321 | Laboratorio Clínico San José | Charité Virology-University of Costa Rica | Andres Moreira-Soto et al |
| EPI_ISL_755301,EPI_ISL_755303 | Laboratorio de Bioingeniería, Instituto de Ciencias de la Ingeniería, Universidad de O´Higgins | Center for Mathematical Modeling and Center for Genome Regulation. Santiago, Chile | M. Latorre et al |
| EPI_ISL_457953,EPI_ISL_457957,EPI_ISL_457962,EPI_ISL_457963,EPI_ISL_457964,EPI_ISL_457967,EPI_ISL_457968,EPI_ISL_457969,EPI_ISL_457971,EPI_ISL_457973,EPI_ISL_480429 | Laboratorio de Biología Molecular Asociación Española Primera en Salud | Departments of Pathology and Medicine, New York University School of Medicine | Maria Victoria Elizondo et al |
| EPI_ISL_842652 | Laboratorio de Biología Molecular Hospital Pedro de Elizalde | Grupo de Genómica y Bioinformática del Instituto de Investigación de la Cadena Láctea CONICET-INTA on behalf of 'Proyecto Argentino Interinstitucional de genomica de SARS-CoV-2' (PAIS Consortium) | Amadio et al |
| EPI_ISL_468755,EPI_ISL_468756 | Laboratorio de Biologia Molecular, Facultad de Medicina, Universidad de Atacama | Center for Mathematical Modeling and Center for Genome Regulation. Santiago, Chile | Gaete A et al |
| EPI_ISL_626560 | Laboratorio de Biología Molecular, Facultad de Medicina, Universidad de Atacama, Copiapo, Chile/ FONDAP CRG, Universidad Andrés Bello, Santiago, Chile | Center for Mathematical Modeling and Center for Genome Regulation. Santiago, Chile | Echeverría C et al |
| EPI_ISL_1278277 | Laboratorio de Biologia Molecular, Hospital San Pedro Claver | Molecular Genetics Laboratory, Instituto de Investigaciones Químicas, Universidad Mayor de San Andrés | Oscar M. Rollano-Peñaloza et al |
| EPI_ISL_1278281,EPI_ISL_1278284 | Laboratorio de Biologia Molecular, SEDES-Potosi | Molecular Genetics Laboratory, Instituto de Investigaciones Químicas, Universidad Mayor de San Andrés | Oscar M. Rollano-Peñaloza et al |

| | | | |
|---|---|---|---|
| EPI_ISL_833132,EPI_ISL_833133,EPI_ISL_1068104,EPI_ISL_1068105,EPI_ISL_1068106,EPI_ISL_1068109 | Laboratorio de Ecologia de Doencas Transmissiveis na Amazonia, Instituto Leonidas e Maria Deane - Fiocruz Amazonia | Laboratorio de Ecologia de Doencas Transmissiveis na Amazonia, Instituto Leonidas e Maria Deane - Fiocruz Amazonia | Valdinete Nascimento et al |
| EPI_ISL_591532 | Laboratorio de Infectologia y virologia molecular | Center for Mathematical Modeling and Center for Genome Regulation. Santiago, Chile | Valiente F et al |
| EPI_ISL_779182,EPI_ISL_961788,EPI_ISL_961793,EPI_ISL_961795 | Laboratorio de Infectología, Servicio de Infectología, Hospital Universitario Dr. José Eleuterio González - Universidad Autónoma de Nuevo León | Laboratorio de Infectología Molecular, Departamento de Bioquímica y Medicina Molecular,Facultad de Medicina Universidad Autónoma de Nuevo León | Kame A. Galán-Huerta et al |
| EPI_ISL_953402,EPI_ISL_953403,EPI_ISL_953410,EPI_ISL_953411,EPI_ISL_953412,EPI_ISL_953418,EPI_ISL_953420,EPI_ISL_953422 | Laboratorio de Investigaciones de Baney | Swiss Tropical and Public Health Institute"" | Carlos Cortes et al" |
| EPI_ISL_648304,EPI_ISL_648314,EPI_ISL_648318,EPI_ISL_648322,EPI_ISL_648337,EPI_ISL_648339,EPI_ISL_648343,EPI_ISL_648358,EPI_ISL_648362,EPI_ISL_648365,EPI_ISL_648379,EPI_ISL_649163 | Laboratorio de Investigaciones de Baney | University Hospital Basel, Clinical Bacteriology | Carlos Cortes et al |
| EPI_ISL_831939 | Laboratório de Microbiologia Molecular - Universidade FEEVALE | Universidade Federal de Ciências da Saúde de Porto Alegre | Vinícius Bonetti Franceschi et al |
| EPI_ISL_476416 | Laboratório de Patologia Clínica - UNICAMP | Laboratório de Estudos de Vírus Emergentes - UNICAMP | José Luiz Proença-Modena et al |
| EPI_ISL_1111416,EPI_ISL_1111421,EPI_ISL_1111425 | Laboratorio de Referencia Nacional de Enteropatógenos. Instituto Nacional de Salud del Perú | Laboratorio de Referencia Nacional de Enteropatógenos. Instituto Nacional de Salud del Perú | Ronnie Gavilan Chavez et al |
| EPI_ISL_517687,EPI_ISL_524473,EPI_ISL_527789 | Laboratorio de Referencia Nacional de Virus Respiratorio. Centro Nacional de Salud Publica. Instituto Nacional de Salud Peru. | Laboratorio de Referencia Nacional de Biotecnología y Biología Molecular. Centro Nacional de Salud Publica. Instituto Nacional de Salud Peru. | Carlos Padilla Rojas et al |
| EPI_ISL_514226,EPI_ISL_514317,EPI_ISL_833038,EPI_ISL_1092335,EPI_ISL_1092356,EPI_ISL_1093153,EPI_ISL_1093158,EPI_ISL_1093163,EPI_ISL_1137479,EPI_ISL_1137483,EPI_ISL_1138419,EPI_ISL_1138420 | Laboratorio de Referencia Nacional de Virus Respiratorio. Instituto Nacional de Salud Perú | Laboratorio de Referencia Nacional de Biotecnología y Biología Molecular. Instituto Nacional de Salud Perú | Carlos Padilla Rojas et al |
| EPI_ISL_1111116,EPI_ISL_1111138,EPI_ISL_1111168,EPI_ISL_1111194,EPI_ISL_1111197,EPI_ISL_1111215,EPI_ISL_1111274,EPI_ISL_1111373 | Laboratorio de Referencia Nacional de Virus Respiratorio. Instituto Nacional de Salud Perú | Laboratorio de Referencia Nacional de Enteropatógenos. Instituto Nacional de Salud del Perú | Ronnie Gavilan Chavez et al |
| EPI_ISL_540933,EPI_ISL_540953,EPI_ISL_540969,EPI_ISL_568542,EPI_ISL_568543,EPI_ISL_729876,EPI_ISL_729889 | Laboratorio de Referencia Nacional de Virus Respiratorios, Instituto Nacional de Salud Peru | Laboratorio de Genómica Microbiana, Universidad Peruana Cayetano Heredia | Pablo Tsukayama et al |
| EPI_ISL_1137615,EPI_ISL_1137619 | Laboratorio de Salud Publica de Cauca | Instituto Nacional de Salud- Dirección de Investigación en Salud Pública | Katherine Laiton-Donato et al |
| EPI_ISL_1091923 | Laboratorio de salud público de Atlantico | Instituto Nacional de Salud- Dirección de Investigación en Salud Pública | Katherine Laiton-Donato et al |
| EPI_ISL_430807,EPI_ISL_792189,EPI_ISL_792203,EPI_ISL_792223 | Laboratorio de Virología del Hospital de Niños Dr. Ricardo Gutierrez | Área de Secuenciación del Laboratorio de Virología del Hospital de Niños Dr. Ricardo Gutierrez on behalf of 'Proyecto Argentino Interinstitucional de genomica de SARS-CoV-2' (PAIS Consortium) | Nabaes Jodar et al |
| EPI_ISL_671974 | Laboratorio de Virología y Microbiología Molecular, Depto. de Microbiología, Facultad de Medicina, Universidad de El Salvador/INS-laboratorio de Ref. Ministerio de Salud | Laboratorio de Virología y Microbiología Molecular, Depto. de Microbiología, Facultad de Medicina, Universidad de El Salvador/INS-laboratorio de Ref. Ministerio de Salud | Rivera NR et al |
| EPI_ISL_792346 | Laboratorio del Hospital Interzonal General de Agudos Evita | Área de Secuenciación del Laboratorio de Virología del Hospital de Niños Dr. Ricardo Gutierrez on behalf of 'Proyecto Argentino Interinstitucional de genomica de SARS-CoV-2' (PAIS Consortium) | Nabaes Jodar et al |

| | | | |
|---|---|---|---|
| EPI_ISL_750163 | Laboratorio DILAVE/MGAP-INIA-UdelaR - Tacuarembó | Institut Pasteur de Montevideo | Daiana Mir et al |
| EPI_ISL_956306 | Laboratorio Gencore- Universidad de los Andes | Instituto Nacional de Salud- Dirección de Investigación en Salud Pública, Universidad de los Andes- Gencore | Katherine Laiton-Donato et al |
| EPI_ISL_1133255,EPI_ISL_1133269,EPI_ISL_1133270,EPI_ISL_1133273,EPI_ISL_1133276 | Laboratório Hermes Pardini | Laboratório de Biologia Integrativa, Instituto de Ciências Biológicas, Universidade Federal de Minas Gerais | Filipe Romero Rebello Moreira et al |
| EPI_ISL_837551,EPI_ISL_837552,EPI_ISL_837579,EPI_ISL_837583,EPI_ISL_837584,EPI_ISL_837585,EPI_ISL_837586,EPI_ISL_837587,EPI_ISL_837589,EPI_ISL_837590,EPI_ISL_837591,EPI_ISL_837592,EPI_ISL_837593,EPI_ISL_837595 | Laboratorio Nacional de Salud | Laboratory of Respiratory Viruses and Measles, Oswaldo Cruz Institute, FIOCRUZ | Paola Resende et al |
| EPI_ISL_1091248,EPI_ISL_1091252 | Laboratorio PGM | Laboratorio de Infectología Molecular, Departamento de Bioquímica y Medicina Molecular,Facultad de Medicina Universidad Autónoma de Nuevo León | Kame A. Galán-Huerta et al |
| EPI_ISL_682277 | LABORATORIOS LABIN | Inciensa, Instituto Costarricense de Investigación y Enseñanza en Nutrición y Salud | Francisco Duarte et al |
| EPI_ISL_961466 | Laboratorios Lister | Instituto de Diagnostico y Referencia Epidemiologicos (INDRE) | Claudia Wong-Arambula et al |
| EPI_ISL_882960,EPI_ISL_882961 | Laboratorium Diagnostyi Mikrobiologicznej z Pracownią Prątka Gruźlicy SPSzW im. Jana Bożego w Lublinie | National Institute of Public Health - National Institute of Hygiene | Wołkowicz Tomasz et al |
| EPI_ISL_1081200 | Laboratory Corporation of America | Respiratory Viruses Branch, Division of Viral Diseases, Centers for Disease Control and Prevention | Peter W. Cook et al |
| EPI_ISL_541654,EPI_ISL_644566 | Laboratory Diagnostic, Veterinary Specialized Institute Kraljevo | Laboratory Diagnostic, Veterinary Specialized Institute Kraljevo | Vidanovic et al |
| EPI_ISL_933533,EPI_ISL_1209407 | Laboratory for HIV and opportunistic infections diagnosis The Republican Research and Practical Center for Epidemiology and Microbiology (RRPCEM) | Laboratory for HIV and opportunistic infections diagnosis The Republican Research and Practical Center for Epidemiology and Microbiology (RRPCEM) | Elena Gasich et al |
| EPI_ISL_455477 | Laboratory for Respiratory Viruses, Cantacuzino National Military-Medical Institute for Research and Development | Cantacuzino Institute | M.Lazar et al |
| EPI_ISL_935017 | Laboratory for Respiratory Viruses, Cantacuzino National Military-Medical Institute for Research and Development | Cantacuzino Institute Virology | Luiza Ustea et al |
| EPI_ISL_417518,EPI_ISL_424978,EPI_ISL_464094,EPI_ISL_660543,EPI_ISL_660544,EPI_ISL_660545,EPI_ISL_667809,EPI_ISL_956321,EPI_ISL_956328 | Laboratory Medicine | Department of Laboratory Medicine, Lin-Kou Chang Gung Memorial Hospital, Taoyuan, Taiwan | Kuo-Chien Tsao et al |
| EPI_ISL_411915 | Laboratory Medicine | Department of Laboratory Medicine, Lin-Kou Chang Gung Memorial Hospital, Taoyuan, Taiwan. | Kuo-Chien Tsao et al |
| EPI_ISL_435045,EPI_ISL_435046,EPI_ISL_435047 | Laboratory of Applied Genetics | RSE National Center for Biotechnology"" | Alexandr Shevtsov et al |
| EPI_ISL_1138541 | Laboratory of Communicable Diseases | 1. Laboratory of Communicable Diseases (Estonia); 2. Eurofins Genomics Europe Sequencing GmbH | Liidia Dotsenko et al |
| EPI_ISL_452355 | Laboratory of Infectious Diseases Center of Beijing Ditan Hospital | Laboratory of Infectious Diseases Center of Beijing Ditan Hospital | Siyuan Yang et al |
| EPI_ISL_717979 | Laboratory of Microbiology and Infectious Diseases, Faculty of Veterinary Medicine, Aristotle University of Thessaloniki, University Campus, 541 24, Thessaloniki, Greece. | Laboratory of Biology, Department of Medicine, Democritus University of Thrace, Alexandroupolis, Greece | Dovrolis N. et al |
| EPI_ISL_654016 | Laboratory of Microbiology, National Reference Lab, Charles Nicolle Hospital; 2-University of Tunis ElManar, Faculty of Medicine of Tunis, LR99ES09, Tunis, Tunisia | 1-Clinical and Experimental Pharmacology Lab, LR16SP02, National Center of Pharmacovigilance, University of Tunis El Manar, Tunis, Tunisia. 2-Neurodegenerative diseases and psychiatric troubles, LR18SP03, Razi Hospital, University of Tunis El Manar, Tunis, Tunisia. 3- Ministry of Health, National Observatory of New and Emerging Diseases, 1006, Tunis, Tunisia | Ilhem Boutiba-Ben Boubaker et al |

| | | | |
|---|---|---|---|
| EPI_ISL_796782,EPI_ISL_803120 | Laboratory of Microbiology, National Reference Lab, Charles Nicolle Hospital; 2-University of Tunis ElManar, Faculty of Medicine of Tunis, LR99ES09, Tunis, Tunisia | Clinical and Experimental Pharmacology Lab, LR16SP02, National Center of Pharmacovigilance, University of Tunis El Manar, Tunis, Tunisia. 2- Neurodegenerative diseases and psychiatric troubles, LR18SP03, Razi Hospital, University of Tunis El Manar, Tunis, Tunisia. 3- Ministry of Health, National Observatory of New and Emerging Diseases, 1006, Tunis, Tunisia | Ilhem Boutiba-Ben Boubaker et al |
| EPI_ISL_852842 | Laboratory of Molecular Biology, Diagnostyka sp. z o.o. | genXone SA, Research & Development Laboratory | Maciej Sykulski et al |
| EPI_ISL_801546,EPI_ISL_801615,EPI_ISL_801635,EPI_ISL_801762,EPI_ISL_801765,EPI_ISL_801767,EPI_ISL_801786 | Laboratory of Molecular Virology, Pontificia Universidad Católica de Chile | MSHS Pathogen Surveillance Program | Leonardo I. Almonacid et al |
| EPI_ISL_454575,EPI_ISL_454586,EPI_ISL_454585 | Laboratory of virology, National Center of Expertise | Laboratory of molecular-genetic research, National Center for Expertise, Kazakhstan National Center for Biotechnology, Kazakhstan | Abdaliyev Askar et al |
| EPI_ISL_454579,EPI_ISL_454582 | Laboratory of virology, National Center of Expertise | Laboratory of molecular-genetic research, National Center of Expertise, Kazakhstan National Center for Biotechnology, Kazakhstan | Abdaliyev Askar et al |
| EPI_ISL_576119 | Laboratory, The Bio Arte Limited | Laboratory, The Bio Arte Limited | Biazzo et al |
| EPI_ISL_861864,EPI_ISL_1036069,EPI_ISL_1036070,EPI_ISL_1233628,EPI_ISL_1233637,EPI_ISL_1233663 | Labormedizinisches Zentrum Dr Risch | University Hospital Basel, Clinical Bacteriology | Tim Roloff et al |
| EPI_ISL_534335 | LabPLUS | Institute of Environmental Science and Research (ESR) | Matt Storey et al |
| EPI_ISL_1082266,EPI_ISL_1250703,EPI_ISL_1250705,EPI_ISL_1250708,EPI_ISL_1250709,EPI_ISL_1250711 | LabPLUS | Institute of Environmental Science and Research (ESR) | Rachel Boyle et al |
| EPI_ISL_548068,EPI_ISL_548080,EPI_ISL_579403,EPI_ISL_579425,EPI_ISL_637092,EPI_ISL_661257,EPI_ISL_661261,EPI_ISL_661263,EPI_ISL_755627,EPI_ISL_755632,EPI_ISL_794610,EPI_ISL_843193,EPI_ISL_877212,EPI_ISL_877221,EPI_ISL_877226,EPI_ISL_1016869,EPI_ISL_1082260 | LabPLUS | Institute of Environmental Science and Research (ESR) | Xiaoyun Ren et al |
| EPI_ISL_1016856 | LabTests | Institute of Environmental Science and Research (ESR) | Xiaoyun Ren et al |
| EPI_ISL_717796,EPI_ISL_717797 | LACEN RJ - Noel Nutels | Bioinformatics Laboratory / LNCC | Carolina M Voloch et al |
| EPI_ISL_572337,EPI_ISL_572349,EPI_ISL_572362,EPI_ISL_572381 | LACEN/PE | WallauLab, Aggeu Magalhaes Institute | Marcelo Henrique Santos Paiva et al |
| EPI_ISL_738323,EPI_ISL_738324,EPI_ISL_812260,EPI_ISL_812269,EPI_ISL_812270,EPI_ISL_812277,EPI_ISL_812278,EPI_ISL_812283 | Landstuhl Regional Medical Center | United States Air Force School of Aerospace Medicine | Anthony Fries et al |
| EPI_ISL_407079 | Lapland Central Hospital | Department of Virology, University of Helsinki and Helsinki University Hospital, Helsinki, Finland | Teemu Smura et al |
| EPI_ISL_1091784,EPI_ISL_1091786 | LDSP | Universidad Nacional de Colombia - Laboratorio Genómico One Health | Andres F. Cardona-Rios et al |
| EPI_ISL_498552,EPI_ISL_498551,EPI_ISL_498556 | Lebanese American University | Lebanese American University | Abi Habib et al |
| EPI_ISL_961467,EPI_ISL_961468,EPI_ISL_961469 | LESP Nuevo Leon/Grupo de Diagnostico ARIES | Instituto de Diagnostico y Referencia Epidemiologicos (INDRE) | Claudia Wong-Arambula et al |
| EPI_ISL_532294 | Lighthouse Lab in Glasgow | Wellcome Sanger Institute for the COVID-19 Genomics UK (COG-UK) Consortium | Harper VanSteenhouse et al |
| EPI_ISL_581117,EPI_ISL_601443,EPI_ISL_600093 | Lighthouse Lab in Milton Keynes | Wellcome Sanger Institute for the COVID-19 Genomics UK (COG-UK) consortium | The Lighthouse Lab in Milton Keynes et al |

| | | | |
|---|---|---|---|
| EPI_ISL_644092 | Lighthouse Lab in Milton Keynes | Wellcome Sanger Institute for the COVID-19 Genomics UK (COG-UK) Consortium | The Lighthouse Lab in Milton Keynes et al |
| EPI_ISL_1008713 | Lister Laboratorio de Referencia S.A. de C.V. | Instituto de diagnóstico y Referencia Epidemiologicos (INDRE) Departamento de Virología | Claudia Wong-Arambula et al |
| EPI_ISL_541870 | Lithuanian University of Health Sciences Hospital, Department of Laboratory Medicine | Lithuanian University of Health Sciences, Laboratory of Molecular Cardiology | Lukas Zemaitis et al |
| EPI_ISL_636604,EPI_ISL_636855 | Lithuanian University of Health Sciences Hospital, Department of Laboratory Medicine | Lithuanian University of Health Sciences, Molecular cardiology lab. | Lukas Zemaitis et al |
| EPI_ISL_453294,EPI_ISL_453340,EPI_ISL_1249267 | Liverpool Clinical Laboratories | COVID-19 Genomics UK (COG-UK) Consortium | Sam Haldenby et al |
| EPI_ISL_469052,EPI_ISL_469053,EPI_ISL_482737,EPI_ISL_482739 | LNR National Reference Laboratory, Mohammed VI University of Health Sciences | Medical Biotechnology Laboratory, Rabat Medical and Pharmacy School, Mohammed The Vth University in Rabat | Meriem LAAMARTI et al |
| EPI_ISL_728236 | LNR National Reference Laboratory, Mohammed VI University of Health Sciences | Medical Biotechnology Laboratory, Rabat Medical and Pharmacy School, Mohammed The Vth University in Rabat | Souad KARTTI et al |
| EPI_ISL_1069188 | Lumban Rural Health Unit | Research Institute for Tropical Medicine | Hannah Leah Morito et al |
| EPI_ISL_644823 | Madigan Army Medical Center | U.S. Air Force School of Aerospace Medicine | Emily Parsons et al |
| EPI_ISL_528743,EPI_ISL_528744,EPI_ISL_582124,EPI_ISL_977593,EPI_ISL_1059900 | Malaysia Genome Institute | Malaysia Genome Institute | Mohd Noor Mat Isa et al |
| EPI_ISL_1196017 | Manzini RRT | National Institute for Communicable Diseases of the National Health Laboratory Service | Maphalala GP et al |
| EPI_ISL_1233323 | Massachusetts State Public Health Laboratory | Massachusetts State Public Health Laboratory | Andrew Lang et al |
| EPI_ISL_1257929,EPI_ISL_1258411,EPI_ISL_1258412 | MB-Cadham Provincial laboratory | National Microbiology Laboratory (NML) | Anna Majer et al |
| EPI_ISL_1196000 | Mbabane Clinic | National Institute for Communicable Diseases of the National Health Laboratory Service | Maphalala GP et al |
| EPI_ISL_1196001,EPI_ISL_1196007 | Mbabane Gov Hospital | National Institute for Communicable Diseases of the National Health Laboratory Service | Maphalala GP et al |
| EPI_ISL_482761,EPI_ISL_482773,EPI_ISL_483038,EPI_ISL_1109484,EPI_ISL_1109486,EPI_ISL_1165080,EPI_ISL_1165081,EPI_ISL_1167189 | Medical Ain Shams Research Institute (MASRI), Ain Shams University | Medical Ain Shams Research Institute (MASRI), Ain Shams University | Hesham Elghazaly et al |
| EPI_ISL_873165 | Medical Laboratory Sciences, Arab American University | Medical Laboratory Sciences, Arab American University | Dumaidi et al |
| EPI_ISL_1254541 | Medical Microbiology Unit, Department for Laboratory Medicine, Drammen Hospital, Vestre Viken Health Trust, | Norwegian Institute of Public Health, Department of Virology | Kathrine Stene-Johansen et al |
| EPI_ISL_882645 | Medical Research Center, Faculty of Medicine, Syarif Hidayatullah State Islamic University Jakarta | Medical Research Center, Faculty of Medicine, Syarif Hidayatullah State Islamic University Jakarta | Chris Adhiyanto et al |
| EPI_ISL_1048371 | Megalab, Molecular and Cytogenetics Diagnostics | Department for Virology, Molecular Biology and Genome Research, R. G. Lugar Center for Public Health Research, National Center for Disease Control and Public Health (NCDC) of Georgia. | Giorgi Tomashvili et al |
| EPI_ISL_1020195 | Melania Hospital, Bogor , West Java | Biosafety Level-3 Laboratory, Indonesian Institute of Sciences (LIPI) | Anggia Prasetyoputri et al |
| EPI_ISL_568923,EPI_ISL_569109,EPI_ISL_569166,EPI_ISL_569176,EPI_ISL_569527,EPI_ISL_569546,EPI_ISL_644411,EPI_ISL_644444,EPI_ISL_804485,EPI_ISL_804494,EPI_ISL_804579,EPI_ISL_900088,EPI_ISL_900194,EPI_ISL_1020398 | MEPHI, Aix Marseille University | MEPHI, Aix Marseille University | Anthony LEVASSEUR et al |
| EPI_ISL_480334,EPI_ISL_480335,EPI_ISL_480338,EPI_ISL_480346 | Microbial Genomics Laboratory, Institut Pasteur de Montevideo | Microbial Genomics Laboratory, Institut Pasteur de Montevideo | Cecilia Salazar et al |
| EPI_ISL_875558 | Microbiologia e Virologia | Istituto Zooprofilattico Sperimentale delle Venezie | Adelaide Milani et al |

| | | | |
|---|---|---|---|
| EPI_ISL_562199,EPI_ISL_562204,EPI_ISL_562212,EPI_ISL_562221,EPI_ISL_562238,EPI_ISL_562252,EPI_ISL_562293 | Microbiological Diagnostic Unit - Public Health Laboratory (MDU-PHL) | MDU-PHL | Seemann et al |
| EPI_ISL_640543,EPI_ISL_640630,EPI_ISL_663451 | Microbiological Diagnostic Unit - Public Health Laboratory (MDU-PHL) | MDU-PHL | Seemann T. et al |
| EPI_ISL_418269 | Microbiology and Immunology department, Pasteur institute in Ho Chi Minh city | Microbiology and Immunology department, Pasteur institute in Ho Chi Minh city | Cao et al |
| EPI_ISL_418267 | Microbiology and Immunology department, Pasteur institute in Ho Chi Minh city | Microbiology and Immunology department, Pasteur institute in Ho Chi Minh city | Nguyen et al |
| EPI_ISL_1208560 | Microbiology Department, Laboratori Clínic Metropolitana Nord. Hospital Universitari Germans Trias i Pujol. | Can Ruti SARS-CoV-2 Sequencing Hub (HUGTiP/IrsiCaixa/IGTP) | Marc Noguera-Julian et al |
| EPI_ISL_1082257,EPI_ISL_1082267 | Middlemore Hospital | Institute of Environmental Science and Research (ESR) | Rachel Boyle et al |
| EPI_ISL_548036,EPI_ISL_548053,EPI_ISL_548132,EPI_ISL_548133,EPI_ISL_755624,EPI_ISL_794625,EPI_ISL_1016844 | Middlemore Hospital | Institute of Environmental Science and Research (ESR) | Xiaoyun Ren et al |
| EPI_ISL_718154,EPI_ISL_718161,EPI_ISL_718164,EPI_ISL_718220 | Ministry of Health Hospitals | Institute of Health and Community Medicine | David Perera et al |
| EPI_ISL_1302723 | Ministry of Health Turkey | Ministry of Health Turkey | Fatma Bayrakdar et al |
| EPI_ISL_903375,EPI_ISL_903377,EPI_ISL_903378,EPI_ISL_903379,EPI_ISL_903380,EPI_ISL_903381,EPI_ISL_903389 | MOH - Jaber Al-Ahmad Hospital (Innovation Research Laboratory) | MOH - Jaber Al-Ahmad Hospital (Innovation Research Laboratory) | Salman Al-Sabah et al |
| EPI_ISL_435124,EPI_ISL_435125,EPI_ISL_435134,EPI_ISL_435137,EPI_ISL_520698,EPI_ISL_520715,EPI_ISL_520720 | Mohammed Bin Rashid University of Medicine and Health Sciences | Al Jalila Genomics Center | Ahmad Abou Tayoun et al |
| EPI_ISL_904016,EPI_ISL_904015 | Molecular Biology and Virology lab, Faculty of Veterinary Medicine, Jordan University of Science and Technology | Molecular Biology and Virology lab, Faculty of Veterinary Medicine, Jordan University of Science and Technology | Mohammad Hussien Alboom et al |
| EPI_ISL_895793,EPI_ISL_895820 | Molecular biology division, Institute of Clinical Biochemistry and Diagnostics, Charles University, Faculty of Medicine in Hradec Králové and University Hospital Hradec Králové | Molecular biology division, Institute of Clinical Biochemistry and Diagnostics, Charles University, Faculty of Medicine in Hradec Králové and University Hospital Hradec Králové | Helena Kovaříková et al |
| EPI_ISL_486827,EPI_ISL_534346 | Molecular diagnostic laboratory of Federal Budget Institution of Science Central Research Institute of Epidemiology" of The Federal Service on Customers' Rights Protection and Human Well-being Surveillance" | Group of Genomics and Postgenomic Technologies of Central Research Institute of Epidemilology | Speranskaya AS et al |
| EPI_ISL_614347,EPI_ISL_614348,EPI_ISL_614349,EPI_ISL_614350,EPI_ISL_614355,EPI_ISL_614361,EPI_ISL_614367,EPI_ISL_614385,EPI_ISL_614386,EPI_ISL_614387,EPI_ISL_614388,EPI_ISL_614389,EPI_ISL_614392,EPI_ISL_614393,EPI_ISL_614395,EPI_ISL_681830,EPI_ISL_681831,EPI_ISL_681840,EPI_ISL_681842 | Molecular diagnostic unit for viral haemorrhagic fevers and emerging viruses, Bouaké CHU Laboratory | Project group Epidemology of Highly Pathogenic Microorganisms, Robert Koch-Institute | Chantal Akoua-Koffi et al |
| EPI_ISL_451957 | Molecular Pathology Division, Department of Pathology, Hong Kong Sanatorium & Hospital | Molecular Pathology Division, Department of Pathology, Hong Kong Sanatorium & Hospital | Chun Hang AU et al |
| EPI_ISL_640039 | Mossel Bay Hospital wc MBY | NHLS/UCT | Arash Iranzadeh et al |
| EPI_ISL_889363,EPI_ISL_889367 | Motol University Hospital | Institute of Applied Biotechnologies a.s. | Petr Klempt et al |
| EPI_ISL_954242,EPI_ISL_954270,EPI_ISL_954299 | MRC/UVRI & LSHTM Uganda Research Unit | Where sequence data have been generated and submitted to GISAID | Matthew Cotten et al |

| | | | |
|---|---|---|---|
| EPI_ISL_561014,EPI_ISL_561015,EPI_ISL_561040,EPI_ISL_561217,EPI_ISL_561234,EPI_ISL_561237,EPI_ISL_561238,EPI_ISL_561267,EPI_ISL_561269,EPI_ISL_561276,EPI_ISL_561283,EPI_ISL_810971,EPI_ISL_810974,EPI_ISL_810982,EPI_ISL_915421,EPI_ISL_915192,EPI_ISL_1216081,EPI_ISL_1216121,EPI_ISL_1216127,EPI_ISL_1234525,EPI_ISL_1234526,EPI_ISL_1234528,EPI_ISL_1234529,EPI_ISL_1234530,EPI_ISL_1234531,EPI_ISL_1234536 | MRCG at LSHTM Genomics lab | MRCG at LSHTM Genomics lab | Abdul Karim sesay et al |
| EPI_ISL_471159,EPI_ISL_471160,EPI_ISL_471161,EPI_ISL_471162,EPI_ISL_471168,EPI_ISL_471170 | MRCG at LSHTM Genomics lab | MRCG at LSHTM Genomics lab | Sesay et al et al |
| EPI_ISL_802416 | MSHS Clinical Microbiology Laboratories | MSHS Pathogen Surveillance Program | Ana S. Gonzalez-Reiche et al |
| EPI_ISL_861758 | Murphy Medical Associates | Grubaugh Lab - Yale School of Public Health | Tara Alpert et al |
| EPI_ISL_1141924 | MVZ Labor Dr. Fenner und Kollegen (Standort Hamburg) | Robert Koch Institute | et al |
| EPI_ISL_1144206 | MVZ Labor Dr. Limbach & Kollegen GbR | Robert Koch Institute | et al |
| EPI_ISL_447918 | n/a | National Institute of Health. Department of medical Sciences, Ministry of Public Health, Thailand | Pilailuk et al |
| EPI_ISL_523968 | National Agency for Public Health, Republic of Moldova | Charite Universitatsmedizin Berlin, Institute of Virology | Victor M Corman et al |
| EPI_ISL_1231534,EPI_ISL_1233470,EPI_ISL_1233521,EPI_ISL_1233558,EPI_ISL_1233559 | National Center for Infectious and Parasitic Diseases (NCIPD) | National Center for Infectious and Parasitic Diseases (NCIPD) | Alexiev et al et al |
| EPI_ISL_1139148,EPI_ISL_1139154,EPI_ISL_1139155,EPI_ISL_1139156 | National Center of Disease Control and Prevention of the Republic of Armenia | Institute of Molecular Biology NAS RA, Republic of Armenia, Department of Bioengineering, BioinformaticsInstitute and Molecular Biology IBMPh RAU, Republic of Armenia | Arsen Arakelyan et al |
| EPI_ISL_454572 | National Center of Expertise | National Center for Expertise, Kazakhstan National Center for Biotechnology, Kazakhstan | Abdaliyev Askar et al |
| EPI_ISL_454571 | National Center of Expertise | National Center for Expertise, National Center for Biotechnology, Kazakhstan | Abdaliyev Askar et al |
| EPI_ISL_497879 | National Centre For Cell Science | National Centre For Cell Science | Dhiraj Paul et al |
| EPI_ISL_436413 | National Centre for Disease control (NCDC) | NCDC/CSIR-IGIB | Pramod Kumar# et al |
| EPI_ISL_435070 | National Centre for Disease control (NCDC), CSIR-Institute of Genomics and Integrative Biology (CSIR-IGIB) | NCDC/CSIR-IGIB | Pramod Kumar et al |
| EPI_ISL_1252724 | National Food and Veterinary Risk Assessment Institute | Vilnius University Hospital Santaros Klinikos, Center of Laboratory Medicine | Gytis Dudas et al |
| EPI_ISL_528604 | National Genomics Core-Center for DNA Fingerprinting and Diagnostics | National Genomics Core- Center for DNA Fingerprinting and Diagnostics (NGC-CDFD)- DBT's PAN-INDIA-1000 Genome consortium | Ashwin Dalal et al |
| EPI_ISL_466843,EPI_ISL_466852,EPI_ISL_528602 | National Genomics Core-Center for DNA Fingerprinting and Diagnostics | National Genomics Core- Center for DNA Fingerprinting and Diagnostics (NGC-CDFD)- DBT's PAN-INDIA-1000 Genome consortium | Bala Pratyusha et al |
| EPI_ISL_528542,EPI_ISL_528543,EPI_ISL_528566,EPI_ISL_528583,EPI_ISL_528628,EPI_ISL_528880 | National Genomics Core-Center for DNA Fingerprinting and Diagnostics | National Genomics Core- Center for DNA Fingerprinting and Diagnostics (NGC-CDFD)- DBT's PAN-INDIA-1000 Genome consortium | G Shashikanth et al |
| EPI_ISL_528554,EPI_ISL_528557,EPI_ISL_528620,EPI_ISL_528635 | National Genomics Core-Center for DNA Fingerprinting and Diagnostics | National Genomics Core- Center for DNA Fingerprinting and Diagnostics (NGC-CDFD)- DBT's PAN-INDIA-1000 Genome consortium | Heena Shah et al |
| EPI_ISL_770475 | National health Laboratory | Botswana Institute for Technology Research and Innovation | Kefentse Arnold Tumedi et al |
| EPI_ISL_770470,EPI_ISL_770473,EPI_ISL_770474,EPI_ISL_770471,EPI_ISL_770472 | National Health laboratory | Botswana Institute for Technology Research and Innovation | Kefentse Arnold Tumedi et al |
| EPI_ISL_560386 | National Health Laboratory | Botswana Institute for Technology Research and innovation | Kefentse Arnold Tumedi et al |

| | | | |
|---|---|---|---|
| EPI_ISL_560385,EPI_ISL_560387,EPI_ISL_560388,EPI_ISL_560389,EPI_ISL_560390 | National Health Laboratory | Botswana Institute for Technology Research and Innovation | Kefentse Arnold Tumedi et al |
| EPI_ISL_622980 | National Health Laboratory Service | National Institute for Communicable Diseases of the National Health Laboratory Service | Allam M et al |
| EPI_ISL_1048553,EPI_ISL_1048554,EPI_ISL_1048556,EPI_ISL_1048563,EPI_ISL_1048564,EPI_ISL_1048567 | National Health Laboratory Service, South Africa | KRISP, KZn Research Innovation and Sequencing Platform | Giandhari J et al |
| EPI_ISL_1080937 | National Health Laboratory Services | National Health Laboratory Services | Mushal Ali et al |
| EPI_ISL_1072960,EPI_ISL_1071111 | National Health Laboratory Services | National Health Laboratory Services, Virology | Mushal Ali et al |
| EPI_ISL_456599,EPI_ISL_456600,EPI_ISL_456604,EPI_ISL_456605,EPI_ISL_456606,EPI_ISL_456612 | National Health Laboratory, Timor-Leste | Microbiological Diagnostic Unit Public Health Laboratory, The Peter Doherty Institute for Infection and Immunity | Soares da Silva et al |
| EPI_ISL_435308,EPI_ISL_435312,EPI_ISL_455707,EPI_ISL_455709,EPI_ISL_455710,EPI_ISL_455712,EPI_ISL_455714,EPI_ISL_455718,EPI_ISL_511892 | National Hospital of Tropical Diseases | Oxford University Clinical Research Unit, Hanoi, Vietnam | Nguyen Thi Tam et al |
| EPI_ISL_486889 | National Influenza Center, Bahrain | National Influenza Center, Bahrain | Altaif et al |
| EPI_ISL_486887 | National Influenza Center, Bahrain | National Influenza Center, Bahrain | Zaed et al |
| EPI_ISL_416431,EPI_ISL_416429 | National Influenza Center, National Institute of Hygiene and Epidemiology (NIHE) | National Influenza Center, National Institute of Hygiene and Epidemiology (NIHE) | Le Quynh Mai et al |
| EPI_ISL_660542 | National Influenza Center, National Institute of Hygiene and Epidemiology (NIHE) | National Institute of Hygiene and Epidemiology (NIHE) | Le Quynh Mai et al |
| EPI_ISL_862078,EPI_ISL_1014679,EPI_ISL_1014684 | National Influenza Center, Virology Department | National Influenza Center | A Nejati et al |
| EPI_ISL_862077,EPI_ISL_862080,EPI_ISL_959277,EPI_ISL_959280,EPI_ISL_959282,EPI_ISL_1014683,EPI_ISL_1014687 | National Influenza Center, Virology Department | National Influenza Center | J Yavarian et al |
| EPI_ISL_862079,EPI_ISL_959279,EPI_ISL_1014680,EPI_ISL_1014685 | National Influenza Center, Virology Department | National Influenza Center | K Sadeghi et al |
| EPI_ISL_862076,EPI_ISL_862081,EPI_ISL_959276,EPI_ISL_959281,EPI_ISL_959283,EPI_ISL_1014677,EPI_ISL_1014682,EPI_ISL_1014686 | National Influenza Center, Virology Department | National Influenza Center | NZ Shafiei Jandaghi et al |
| EPI_ISL_862075,EPI_ISL_959275,EPI_ISL_959284,EPI_ISL_1014676,EPI_ISL_1014681 | National Influenza Center, Virology Department | National Influenza Center | V Salimi et al |
| EPI_ISL_476835 | National Influenza Centre for Northern Greece | National Influenza Centre for Northern Greece | Maria Christoforidi et al |
| EPI_ISL_410301 | National Influenza Centre, National Public Health Laboratory, Kathmandu, Nepal | The University of Hong Kong | Ranjit Sah et al |
| EPI_ISL_402125 | National Institute for Communicable Disease Control and Prevention (ICDC) Chinese Center for Disease Control and Prevention (China CDC) | National Institute for Communicable Disease Control and Prevention (ICDC) Chinese Center for Disease Control and Prevention (China CDC) | Zhang et al |
| EPI_ISL_450296,EPI_ISL_450495,EPI_ISL_490257,EPI_ISL_490258,EPI_ISL_490265,EPI_ISL_490269,EPI_ISL_514413,EPI_ISL_622925 | National Institute for Communicable Diseases of the National Health Laboratory Service | National Institute for Communicable Diseases of the National Health Laboratory Service | Allam M et al |
| EPI_ISL_469254 | National Institute for Viral Disease Control and Prevention, China CDC | Institute of Viral Disease Control and Prevention, China CDC | Wenjie Tan et al |
| EPI_ISL_469255 | National Institute for Viral Disease Control and Prevention, China CDC | Institute of Viral Disease Control and Prevention, China CDC | Xiang Zhao et al |
| EPI_ISL_591272,EPI_ISL_591280,EPI_ISL_591279,EPI_ISL_591275,EPI_ISL_591274,EPI_ISL_591277,EPI_ISL_591276 | National Institute for Viral Disease Control and Prevention, China CDC | National Institute for Viral Disease Control and Prevention, China CDC | Huilai Ma et al |

| | | | |
|---|---|---|---|
| EPI_ISL_469256,EPI_ISL_498691,EPI_ISL_498692,EPI_ISL_498694,EPI_ISL_850948,EPI_ISL_850949,EPI_ISL_850950,EPI_ISL_850951 | National Institute for Viral Disease Control and Prevention, China CDC | National Institute for Viral Disease Control and Prevention, China CDC | Xiang Zhao et al |
| EPI_ISL_709542 | National Institute of Blood Diseases (NIBD), Molecular Biology Lab | Genomics Lab NIBD | Samina Naz Mukry et al |
| EPI_ISL_1169046 | National Institute of Health Research and Development | National Institute of Health Research and Development | Hana Apsari Pawestri et al |
| EPI_ISL_538508 | National Institute of Health Research and Development | National Institute of Health Research and Development | Pawestri et al |
| EPI_ISL_747234 | National Institute of Health Research and Development | National Institute of Health Research and Development | Subangkit et al |
| EPI_ISL_479758,EPI_ISL_479757 | National Institute of Hygiene and Epidemiology (NIHE) | National Key Laboratory of Gene Technology, Institute of Biotechnology (IBT) | Le Tung Lam et al |
| EPI_ISL_1116448,EPI_ISL_1279947,EPI_ISL_1279967 | National Institute of Infectious Diseases-Prof. Dr. Matei Bals Molecular Diagnostics Laboratory | National Institute of Infectious Diseases-Prof. Dr. Matei Bals Molecular Diagnostics Laboratory | Leontina Banica et al |
| EPI_ISL_603243 | National Institute of Laboratory Medicine and Referral Center | Genomic Research Lab, BCSIR | Barna Goswami et al |
| EPI_ISL_603244,EPI_ISL_982505 | National Institute of Laboratory Medicine and Referral Center | Genomic Research Lab, BCSIR | Iffat Jahan et al |
| EPI_ISL_983330 | National Institute of Laboratory Medicine and Referral Center | Genomic Research Lab, BCSIR | Md. Ahashan Habib et al |
| EPI_ISL_959386 | National Institute of Laboratory Medicine and Referral Center | Genomic Research Lab, BCSIR | Md. Maruf Ahmed Molla et al |
| EPI_ISL_603240 | National Institute of Laboratory Medicine and Referral Center | Genomic Research Lab, BCSIR | Tanjina Akhter Banu et al |
| EPI_ISL_539781,EPI_ISL_539778 | National Institute of Public Health (Czech Republic) | State Veterinary Institute Prague | Nagy et al |
| EPI_ISL_434558 | National Institutes of Health, University of the Philippines Manila | Philippine Genome Center | Carlo M. Lapid et al |
| EPI_ISL_1267011,EPI_ISL_1267023 | National Laboratory for Health, Environment and Food, OMM, Maribor | CISLD (Clinical Institute of Special Laboratory Diagnostics), University Children's Hospital, University Medical Center Ljubljana | Jernej Kovač et al |
| EPI_ISL_512616,EPI_ISL_512633,EPI_ISL_512640 | National Laboratory for Influenza/Virology reference laboratory, Public Health Center of the Ministry of Health of Ukraine | Respiratory Virus Unit, Microbiology Services Colindale, Public Health England | PHE Covid Sequencing Team et al |
| EPI_ISL_1257898 | National Medicines Institute | DNA Sequencing and Synthesis Facility (oligo.pl), Institute of Biochemistry and Biophysics PAS | Gawor Jan et al |
| EPI_ISL_1191785,EPI_ISL_1191797,EPI_ISL_1191816,EPI_ISL_1191818,EPI_ISL_1191819,EPI_ISL_1191822,EPI_ISL_1191824,EPI_ISL_1191828,EPI_ISL_1191830,EPI_ISL_1191831,EPI_ISL_1191832,EPI_ISL_1191833,EPI_ISL_1191834,EPI_ISL_1191845,EPI_ISL_1191898,EPI_ISL_1191947,EPI_ISL_1191991,EPI_ISL_1192002,EPI_ISL_1192020,EPI_ISL_1192028 | National Microbiology Reference Laboratory | Quadram Institute Bioscience | Tapfumanei Mashe et al |
| EPI_ISL_647971,EPI_ISL_647977,EPI_ISL_647978,EPI_ISL_647980 | National Microbiology Reference Laboratory | Quadram Institute Bioscience | Thanh Le Viet et al |
| EPI_ISL_1039982,EPI_ISL_1095610,EPI_ISL_1195207 | National Public Health Center, COVID Laboratory | National Public Health Center, National Biosafety Laboratory | Bernadett Pályi et al |
| EPI_ISL_457834,EPI_ISL_457843,EPI_ISL_457844 | National Public Health Laboratory | KEMRI-Wellcome Trust Research Programme/KEMRI-CGMR-C Kilifi | Githinji G. et al 2020 et al |
| EPI_ISL_416866,EPI_ISL_416885,EPI_ISL_416884,EPI_ISL_416907 | National Public Health Laboratory | Malaysia Genome Institute | Mohd Noor Mat Isa et al |
| EPI_ISL_754072 | National Public Health Laboratory | Nepal Health Research Council | Pradip Gyanwali et al |

| | | | |
|---|---|---|---|
| EPI_ISL_845546,EPI_ISL_845548,EPI_ISL_845549,EPI_ISL_845550,EPI_ISL_845551,EPI_ISL_845552,EPI_ISL_845553,EPI_ISL_845554,EPI_ISL_845557,EPI_ISL_845558,EPI_ISL_845560,EPI_ISL_845561,EPI_ISL_845562,EPI_ISL_845563,EPI_ISL_845565 | National Public Health Laboratory, Cameroon | African Centre of Excellence for Genomics of Infectious Diseases (ACEGID), Redeemer's University | Oluniyi P.E. et al et al |
| EPI_ISL_418994,EPI_ISL_462332,EPI_ISL_469113,EPI_ISL_475982,EPI_ISL_475984,EPI_ISL_475998,EPI_ISL_479594,EPI_ISL_479597,EPI_ISL_483615,EPI_ISL_493404,EPI_ISL_498579,EPI_ISL_498591,EPI_ISL_498617,EPI_ISL_536423,EPI_ISL_536427,EPI_ISL_536453,EPI_ISL_536454,EPI_ISL_548976 | National Public Health Laboratory, National Centre for Infectious Diseases | National Public Health Laboratory, National Centre for Infectious Diseases | Mak TM et al |
| EPI_ISL_596480,EPI_ISL_596491,EPI_ISL_596495,EPI_ISL_605817,EPI_ISL_626628,EPI_ISL_728189,EPI_ISL_754076,EPI_ISL_754082,EPI_ISL_754083,EPI_ISL_803959,EPI_ISL_803960,EPI_ISL_803961,EPI_ISL_803962,EPI_ISL_825083,EPI_ISL_833373,EPI_ISL_833375,EPI_ISL_857471,EPI_ISL_862820,EPI_ISL_937516,EPI_ISL_981010,EPI_ISL_981011,EPI_ISL_995295,EPI_ISL_1173250,EPI_ISL_1173251,EPI_ISL_1252449 | National Public Health Laboratory, National Centre for Infectious Diseases | National Public Health Laboratory, National Centre for Infectious Diseases | Tze Minn Mak et al |
| EPI_ISL_1122419,EPI_ISL_1122420,EPI_ISL_1122421 | National Public Health Laboratory, National Centre for Infectious Diseases | National Virology Reference Laboratory | Tze Minn Mak et al |
| EPI_ISL_1273391,EPI_ISL_1273392,EPI_ISL_1273393,EPI_ISL_1273394,EPI_ISL_1273395,EPI_ISL_1273396,EPI_ISL_1273399,EPI_ISL_1273401,EPI_ISL_1273406,EPI_ISL_1273407,EPI_ISL_1273408 | National Reference Laboratory - Ministry of Health Maseru Lesotho | National Institute for Communicable Diseases of the National Health Laboratory Service | Gorova V et al |
| EPI_ISL_480297,EPI_ISL_480299,EPI_ISL_480302 | National Reference Laboratory Influenza and acute respiratory diseases"" | NRL-HIV | Ivan Ivanov et al |
| EPI_ISL_625683,EPI_ISL_833578 | National Reference Laboratory for COVID-19, Pasteur Institute of Iran | National Reference Laboratory for COVID-19, Pasteur Institute of Iran | Zahra Ahmadi et al |
| EPI_ISL_737200,EPI_ISL_1073135 | National Reference Laboratory, Nigeria Centre for Disease Control. | National Reference Laboratory, Nigeria Centre for Disease Control, Gaduwa,Abuja, Nigeria | Dr Ndodo Nnaemeka et al |
| EPI_ISL_435674,EPI_ISL_435675,EPI_ISL_435676,EPI_ISL_443187 | National Virology Reference Laboratory | National Public Health Laboratory, National Centre for Infectious Diseases | Mak Tze Minn et al |
| EPI_ISL_962877,EPI_ISL_962878 | National Virology Reference Laboratory | National Public Health Laboratory, National Centre for Infectious Diseases | Tze Minn Mak et al |
| EPI_ISL_501260,EPI_ISL_578219,EPI_ISL_639897,EPI_ISL_732476,EPI_ISL_837363,EPI_ISL_875367,EPI_ISL_909901,EPI_ISL_909902 | National Virus Reference Laboratory | National Virus Reference Laboratory | Michael Carr et al |
| EPI_ISL_1252151,EPI_ISL_1299429 | National Virus Reference Laboratory | National Virus Reference Laboratory | Zoe Yandle et al |
| EPI_ISL_444996,EPI_ISL_444998,EPI_ISL_444999,EPI_ISL_445000 | Naval Health Research Center | Naval Medical Research Center Biological Defense Research Directorate | Logan Voegtly et al |

| | | | |
|---|---|---|---|
| EPI_ISL_1272242,EPI_ISL_1272243 | NB-Hôpital Georges L. Dumont | National Microbiology Laboratory (NML) | Anna Majer et al |
| EPI_ISL_754068,EPI_ISL_754069,EPI_ISL_754070 | Nepal Korea Friendship Municipality Hospital | Nepal Health Research Council | Pradip Gyanwali et al |
| EPI_ISL_678581,EPI_ISL_678586 | Netcare | KRISP, KZN Research Innovation and Sequencing Platform | Giandhari J et al |
| EPI_ISL_1061033 | New South Wales Health Pathology Royal Prince Alfred Hospital | Microbiology RPAH | Foster et al |
| EPI_ISL_854991,EPI_ISL_854988 | NGS Lab, DNA SOLUTION LTD. | NGS Lab, DNA SOLUTION LTD. | Khan et al |
| EPI_ISL_1196008,EPI_ISL_1196009,EPI_ISL_1196010 | Nhlangano Health Centre | National Institute for Communicable Diseases of the National Health Laboratory Service | Maphalala GP et al |
| EPI_ISL_467492,EPI_ISL_495516,EPI_ISL_495522,EPI_ISL_495526,EPI_ISL_495560,EPI_ISL_498054,EPI_ISL_509370,EPI_ISL_515609,EPI_ISL_515632,EPI_ISL_515765,EPI_ISL_518033,EPI_ISL_518036,EPI_ISL_529727,EPI_ISL_535401,EPI_ISL_535453,EPI_ISL_602792,EPI_ISL_602805,EPI_ISL_605789 | NHLS-IALCH | KRISP, KZN Research Innovation and Sequencing Platform | Giandhari J et al |
| EPI_ISL_418242,EPI_ISL_420037,EPI_ISL_766864,EPI_ISL_766865,EPI_ISL_766866,EPI_ISL_766869,EPI_ISL_766871,EPI_ISL_766873 | NIC Viral Respiratory Unit - Institut Pasteur of Algeria | National Reference Center for Viruses of Respiratory Infections, Institut Pasteur, Paris | Mélanie Albert et al |
| EPI_ISL_516936,EPI_ISL_516934 | Nicolae Testemitanu State University of Medicine and Pharmacy | International Centre for Genetic Engineering and Biotechnology (ICGEB) and ARGO Open Lab Platform for Genome Sequencing | Ulinici M et al |
| EPI_ISL_825714,EPI_ISL_1073139,EPI_ISL_1073640,EPI_ISL_1073639,EPI_ISL_1197066,EPI_ISL_1197065,EPI_ISL_1197129 | Nigeria Centre For Disease Control | National reference Laboratory, NCDC, Gaduwa, Abuja | Dr Ndodo Nnaemeka et al |
| EPI_ISL_872601,EPI_ISL_872602,EPI_ISL_872604,EPI_ISL_941281,EPI_ISL_941283,EPI_ISL_941285,EPI_ISL_1242019 | Nigeria Centre for Disease Control (NCDC) | African Centre of Excellence for Genomics of Infectious Diseases (ACEGID), Redeemer's University | Oluniyi P.E. et al et al |
| EPI_ISL_455429,EPI_ISL_487092,EPI_ISL_487099,EPI_ISL_487098 | Nigeria Centre for Disease Control (NCDC) | African Centre of Excellence for Genomics of Infectious Diseases (ACEGID), Redeemer's University, Ede, Osun State, Nigeria | Oluniyi P.E. et al |
| EPI_ISL_527880,EPI_ISL_527911,EPI_ISL_729939,EPI_ISL_729942,EPI_ISL_729944,EPI_ISL_729945,EPI_ISL_729949,EPI_ISL_729971,EPI_ISL_729972,EPI_ISL_729973,EPI_ISL_729975,EPI_ISL_729976,EPI_ISL_729977,EPI_ISL_729978,EPI_ISL_729979,EPI_ISL_729980,EPI_ISL_729981,EPI_ISL_729982,EPI_ISL_729984,EPI_ISL_729985,EPI_ISL_730001,EPI_ISL_730025,EPI_ISL_730030,EPI_ISL_730032,EPI_ISL_730043 | Nigeria Centre for Disease Control (NCDC) | African Centre of Excellence for Genomics of Infectious Diseases (ACEGID), Redeemer's University, Ede, Osun State, Nigeria | Oluniyi P.E. et al et al |
| EPI_ISL_977540 | Nigeria Centre of Disease Control (NCDC) | African Centre of Excellence for Genomics of Infectious Diseases (ACEGID), Redeemer’s University | Olawoye I. B. et al et al |
| EPI_ISL_1093433,EPI_ISL_1235667 | Nigerian Centre for Disease Control (NCDC) | African Centre of Excellence for Genomics of Infectious Diseases (ACEGID), Redeemer’s University | Olawoye et al |
| EPI_ISL_479506,EPI_ISL_1034170,EPI_ISL_1034184 | NIV Influenza | NIV Influenza | Potdar V et al |
| EPI_ISL_1254248 | NJDOH, Public Health and Environmental Laboratories | NJ_PHEL | Lindsey Bodnar et al |

| | | | |
|---|---|---|---|
| EPI_ISL_1278101 | NL-Dr. Leonard A. Miller Centre for Health Services | National Microbiology Laboratory (NML) | Anna Majer et al |
| EPI_ISL_960304 | NLZOH, Laboratory for Virology | NLZOH, Laboratory for Virology | Katarina Prosenc (Laboratory for Virology) et al |
| EPI_ISL_422404,EPI_ISL_422405,EPI_ISL_422399,EPI_ISL_422394,EPI_ISL_422384 | NMIMR, Department of Virology | WACCBIP, University of Ghana | Joyce M. Ngoi et al |
| EPI_ISL_548062,EPI_ISL_579093,EPI_ISL_579094,EPI_ISL_579095,EPI_ISL_579096,EPI_ISL_579406 | North Shore Hospital | Institute of Environmental Science and Research (ESR) | Xiaoyun Ren et al |
| EPI_ISL_453501 | Northumbria University / South Tees Hospitals NHS Foundation Trust / North Cumbria Integrated Care NHS Foundation Trust / North Tees and Hartlepool NHS Foundation Trust / Newcastle Hospitals NHS Foundation Trust | COVID-19 Genomics UK (COG-UK) Consortium | Darren L Smith et al |
| EPI_ISL_416742 | NRL for Influenza, Centrum Epidemiology and Microbiology of National Institute of Public Health, Czech Republic | Charite Universitaetsmedizin Berlin, Institute of Virology | Victor M Corman et al |
| EPI_ISL_1272330,EPI_ISL_1272332,EPI_ISL_1272340,EPI_ISL_1272343 | NS-QEII Health Sciences Centre | National Microbiology Laboratory (NML) | Anna Majer et al |
| EPI_ISL_735447,EPI_ISL_735448 | Nucleic Acid Testing - Rwanda National Reference Laboratory | GIGA Medical Genomics | Yvan Butera et al |
| EPI_ISL_925891,EPI_ISL_925893,EPI_ISL_925900,EPI_ISL_925901,EPI_ISL_925902,EPI_ISL_925904,EPI_ISL_960228,EPI_ISL_960236,EPI_ISL_960237,EPI_ISL_960242,EPI_ISL_960265,EPI_ISL_960276,EPI_ISL_960278,EPI_ISL_960283,EPI_ISL_960300,EPI_ISL_960301,EPI_ISL_960302,EPI_ISL_1301743,EPI_ISL_1301744,EPI_ISL_1301746,EPI_ISL_1301759,EPI_ISL_1301795,EPI_ISL_1301796,EPI_ISL_1301797,EPI_ISL_1302680,EPI_ISL_1302681 | Nucleic Acid Testing, National Reference Laboratory | GIGA Medical Genomics | Yvan Butera et al |
| EPI_ISL_1117409,EPI_ISL_1117411,EPI_ISL_1117418,EPI_ISL_1117446,EPI_ISL_1117448 | Nucleo de Pesquisa em Inovacao Terapeutica - UFPE | LABBE, Federal University of Pernambuco | Wilson Jose da Silva Junior et al |
| EPI_ISL_1060650,EPI_ISL_1060652,EPI_ISL_1262510 | NYU Langone Health | Departments of Pathology and Medicine, New York University School of Medicine | Adriana Heguy et al |
| EPI_ISL_1138863 | Office of Diseases Prevention and Control Region 4 Saraburi | COVID-19 Network Investigations (CONI) Alliance | Elizabeth Batty et al |
| EPI_ISL_667382 | OHSU Lab Services Molecular Microbiology Lab | Oregon SARS-CoV-2 Genome Sequencing Center | Brendan L. O'Connell et al |
| EPI_ISL_745126 | Okiep CHC | National Health Laboratory Service (NHLS), Tygerberg | Susan Engelbrecht et al |
| EPI_ISL_889372 | Olomouc University Hospital | Institute of Applied Biotechnologies a.s. | Petr Klempt et al |
| EPI_ISL_491162,EPI_ISL_491163,EPI_ISL_491164 | Oman-National Influenza Center | Biotechnology & OMICs Laboratory | Sajjad Asaf et al |
| EPI_ISL_491136,EPI_ISL_491139,EPI_ISL_491140 | Oman-National Influenza Center | Biotechnology & OMICs Laboratory | Samiha Al-Kharusi et al |
| EPI_ISL_492065 | Oman-National Influenza Center | Department of Microbiology and Immunology-SQUH Department of Microbiology and Immunology, Sultan Qaboos University Hopsital, P.O 35, Postal code 123 | Samira Al-Maruqi et al |
| EPI_ISL_766569,EPI_ISL_766570 | Oman-National Influenza Center | Oman-National Influenza Center | Samiha Al-Kharusi et al |
| EPI_ISL_457703 | Oman-NIC | Department of Microbiology and Immunology- SQUH | Fahad Zadjali et al |

| | | | |
|---|---|---|---|
| EPI_ISL_491969,EPI_ISL_492001,EPI_ISL_492014,EPI_ISL_492022,EPI_ISL_492024 | Oman-NIC | Department of Microbiology and Immunology-SQUH | Fahad Zadjali et al |
| EPI_ISL_457701,EPI_ISL_457704,EPI_ISL_457979,EPI_ISL_457989,EPI_ISL_457992 | Oman-NIC | Oman-NIC | Samira Al-Maruqi et al |
| EPI_ISL_457705 | OMAN-NIC | Department of Microbiology and Immunology- SQUH | Fahad Zadjali et al |
| EPI_ISL_569785 | Omsk Research Institute of Natural Focal Infections | WHO National Influenza Centre Russian Federation | Artem Fadeev et al |
| EPI_ISL_854800,EPI_ISL_854801,EPI_ISL_854802,EPI_ISL_854806 | Ontario's COVID-19 Genomics Rapid Response Coalition | McMaster University | Ana Cabrera et al |
| EPI_ISL_648507 | Orange County Public Health Lab | Chan-Zuckerberg Biohub | CZB Cliahub Consortium et al |
| EPI_ISL_1040028,EPI_ISL_1040029,EPI_ISL_1040030 | Original detection - Virology Unit, Institut Pasteur du Cambodge; Sequencing - US National Institute of Allergy and Infectious Diseases Cambodia | Virology Unit, Institut Pasteur du Cambodge | Jennifer Bohl et al |
| EPI_ISL_590910,EPI_ISL_635172 | Oslo University Hospital, Department of Medical Microbiology | Norwegian Institute of Public Health, Department of Virology | Kathrine Stene-Johansen et al |
| EPI_ISL_429233,EPI_ISL_418261 | Ospedale Civile Giuseppe Mazzini | Istituto Zooprofilattico Sperimentale dell'Abruzzo e Molise G. Caporale"" | Lorusso A et al |
| EPI_ISL_420592 | Ospedale Civile Giuseppe Mazzini | Istituto Zooprofilattico Sperimentale dell'Abruzzo e Molise G.Caporale"" | Lorusso A et al |
| EPI_ISL_635116 | Ostfold Hospital Trust - Kalnes, Centre for Laboratory Medicine, Section for gene technology and infection serology | Norwegian Institute of Public Health, Department of Virology | Kathrine Stene-Johansen et al |
| EPI_ISL_498184,EPI_ISL_498186 | OUCRU | OUCRU | Nguyen Van Vinh Chau et al |
| EPI_ISL_1001385,EPI_ISL_1001438 | Outre mer | National Reference Center for Viruses of Respiratory Infections, Institut Pasteur, Paris | Marion Barbet et al |
| EPI_ISL_954044,EPI_ISL_954125,EPI_ISL_1201529 | Outre Mer | National Reference Center for Viruses of Respiratory Infections, Institut Pasteur, Paris | Marion Barbet et al |
| EPI_ISL_1273214,EPI_ISL_1293350 | Oxford University Clinical Research Unit (OUCRU) | Oxford University Clinical Research Unit (OUCRU) | Nguyen Van Vinh Chau et al |
| EPI_ISL_596508,EPI_ISL_596519,EPI_ISL_596522,EPI_ISL_596525,EPI_ISL_596528,EPI_ISL_596529 | Palestinian Ministry of Health | Molecular Genetics Lab | Nouar Qutob et al |
| EPI_ISL_955905 | Pamela Youde Nethersole Eastern Hospital | Hong Kong Department of Health | Alan K.L. Tsang et al |
| EPI_ISL_1173020,EPI_ISL_1258495 | Pandemic Response Lab - NYC | Pandemic Response Lab, R&D | Henry Lee et al |
| EPI_ISL_430845 | Pasig City General Hospital | Research Institute for Tropical Medicine | Medado et al |
| EPI_ISL_654881 | Pasteur Institute in Ho Chi Minh city | Department of Microbiology and Immunology - Pasteur Institute in Ho Chi Minh city | Lương Chấn Quang et al |
| EPI_ISL_812922 | Pasteur Institute in Ho Chi Minh city | Department of Microbiology and Immunology - Pasteur Institute in Ho Chi Minh city | Manh Huy Dao et al |
| EPI_ISL_442523 | Pasteur Institute of Iran | Kawsar Human Genetic Research Company | Sirous Zeinali et al |
| EPI_ISL_463749 | Pasteur Institute of Iran | Rapid Response Team | Mahboobeh Rafigh et al |
| EPI_ISL_660153,EPI_ISL_660155,EPI_ISL_660156,EPI_ISL_912401 | PathCare | National Health Laboratory Service (NHLS), Tygerberg | Susan Engelbrecht et al |

| | | | |
|---|---|---|---|
| EPI_ISL_591411,EPI_ISL_591435,EPI_ISL_591448,EPI_ISL_591449,EPI_ISL_591466,EPI_ISL_591467,EPI_ISL_591468,EPI_ISL_591469,EPI_ISL_591470,EPI_ISL_591535,EPI_ISL_667606,EPI_ISL_667654,EPI_ISL_667667,EPI_ISL_685390,EPI_ISL_685395,EPI_ISL_685396,EPI_ISL_685584,EPI_ISL_685644,EPI_ISL_689173,EPI_ISL_690635,EPI_ISL_690698,EPI_ISL_690711,EPI_ISL_690939,EPI_ISL_692046,EPI_ISL_721574,EPI_ISL_735442,EPI_ISL_736891,EPI_ISL_768658,EPI_ISL_768687,EPI_ISL_768707,EPI_ISL_768714,EPI_ISL_779220,EPI_ISL_779238,EPI_ISL_892703,EPI_ISL_900981,EPI_ISL_1129120,EPI_ISL_1129123,EPI_ISL_1129151,EPI_ISL_1129152,EPI_ISL_1129154,EPI_ISL_1129157 | Pathogen Genomics Center, National Institute of Infectious Diseases | Pathogen Genomics Center, National Institute of Infectious Diseases | Tsuyoshi Sekizuka et al |
| EPI_ISL_512999 | Pathogen Genomics Lab King Abdullah University of Science and Technology(KAUST) | Pathogen Genomics Lab King Abdullah University of Science and Technology(KAUST) | Amit Kumar Subudhi et al |
| EPI_ISL_677914,EPI_ISL_677925 | Pathogen Genomics Lab King Abdullah University of Science and Technology(KAUST) | Pathogen Genomics Lab King Abdullah University of Science and Technology(KAUST) | Muhammad Shuaib et al |
| EPI_ISL_678095 | Pathogen Genomics Lab King Abdullah University of Science and Technology(KAUST) | Pathogen Genomics Lab King Abdullah University of Science and Technology(KAUST) | Olga Douvropoulou et al |
| EPI_ISL_512890,EPI_ISL_512898,EPI_ISL_678181,EPI_ISL_678191,EPI_ISL_751222 | Pathogen Genomics Lab King Abdullah University of Science and Technology(KAUST) | Pathogen Genomics Lab King Abdullah University of Science and Technology(KAUST) | Raeece Naeem et al |
| EPI_ISL_513053 | Pathogen Genomics Lab King Abdullah University of Science and Technology(KAUST) | Pathogen Genomics Lab King Abdullah University of Science and Technology(KAUST) | Rahul P Salunke et al |
| EPI_ISL_512946,EPI_ISL_512979,EPI_ISL_677920,EPI_ISL_751236 | Pathogen Genomics Lab King Abdullah University of Science and Technology(KAUST) | Pathogen Genomics Lab King Abdullah University of Science and Technology(KAUST) | Sara Mfarrej et al |
| EPI_ISL_512910 | Pathogen Genomics Lab King Abdullah University of Science and Technology(KAUST) | Pathogen Genomics Lab King Abdullah University of Science and Technology(KAUST) | Sharif Hala et al |
| EPI_ISL_733164,EPI_ISL_733213 | Pathogenic Microorganisms Variability Laboratory | WHO National Influenza Centre Russian Federation | Andrey Komissarov et al |
| EPI_ISL_956048 | Pathology and Laboratory Medicine Institute, Cleveland Clinic, Ohio, USA | Pathology and Laboratory Medicine Institute, Cleveland Clinic, Ohio, USA | Frank P. Esper et al |
| EPI_ISL_596779,EPI_ISL_596781,EPI_ISL_605881,EPI_ISL_810796,EPI_ISL_810965,EPI_ISL_933801,EPI_ISL_1198803 | PathWest Laboratory Medicine WA | PathWest Laboratory Medicine WA Microbial Surveillance Unit | PathWest Laboratory Medicine WA Microbial Surveillance Unit et al |
| EPI_ISL_1063627,EPI_ISL_1063774 | PGIMER, Chandigarh | ICMR-NATIONAL INSTITUTE OF VIROLOGY, MICROBIAL CONTAINMENT COMPLEX | Pragya D. Yadav et al |
| EPI_ISL_1081793,EPI_ISL_1081794 | Philippine Red Cross | Philippine Genome Center | Francis A. Tablizo et al |
| EPI_ISL_1241841 | PHV-FSS | PHV-FSS | Son Nguyen et al |
| EPI_ISL_1194612 | Plateforme de testing Namuroise | Plateforme de testing Namuroise | Céline Maschietto et al |
| EPI_ISL_539851 | Pok Oi Hospital | Hong Kong Department of Health | Alan K.L. Tsang et al |
| EPI_ISL_759955 | Primasatya Husada Citra Hospital | Institute of Tropical Disease, Universitas Airlangga | Aldise M Nastri et al |
| EPI_ISL_956307 | Primasatya Husada Citra Hospital | Institute of Tropical Disease, Universitas Airlangga | Krisnoadi Rahardjo et al |
| EPI_ISL_417064 | Prince of Wales Hospital | Hong Kong Department of Health | Alan K.L. Tsang et al |
| EPI_ISL_1093397 | Princess Haya Biotechnology Center/ Jordan University of Science & Technology | Princess Haya Biotechnology Center/ Jordan University of Science & Technology | Saied Jaradat et al |

| | | | |
|---|---|---|---|
| EPI_ISL_574492 | Programme in Emerging Infectious Diseases, Duke-NUS Medical School | National Public Health Laboratory, National Centre for Infectious Diseases | Tze Minn Mak et al |
| EPI_ISL_523959 | Pronto Socorro Municipal de Perus | Instituto Adolfo Lutz, Interdiciplinary Procedures Center, Strategic Laboratory | Claudio Tavares Sacchi et al |
| EPI_ISL_693231 | Pronto Socorro Municipal de Santa Branca | Instituto Adolfo Lutz, Interdiciplinary Procedures Center, Strategic Laboratory | Claudio Tavares Sacchi et al |
| EPI_ISL_887244 | Protzer Lab | Protzer Lab, Gagneur Lab, Robert Koch Institut | Ulrike Protzer et al |
| EPI_ISL_1112753 | Public Health Center of Ukraine | Charité Universitätsmedizin Berlin, Institute of Virology | Victor M Corman et al |
| EPI_ISL_636973 | Public Health Lab | Public Health Lab | Alwasti et al |
| EPI_ISL_425177 | Public Health Ontario | Public Health Agency of Canada - National Microbiology Laboratory | Amrit S. Boese et al |
| EPI_ISL_413015 | Public Health Ontario Laboratory | National Microbiology Laboratory | Shari Tyson et al |
| EPI_ISL_693282,EPI_ISL_693284,EPI_ISL_693285 | Public Health Virology Laboratory, Forensic and Scientific Services (PHV-FSS) | Public Health Virology Laboratory, Forensic and Scientific Services (PHV-FSS) | Son Nguyen et al et al |
| EPI_ISL_513312,EPI_ISL_513313 | Public Health, United States Air Force School of Aerospace Medicine | Public Health, United States Air Force School of Aerospace Medicine | Fries et al |
| EPI_ISL_1055750 | QEII Health Sciences Centre | National Microbiology Laboratory (NML) | Anna Majer et al |
| EPI_ISL_1249065 | Queens Medical Centre, Clinical Microbiology Department / DeepSeq Nottingham | COVID-19 Genomics UK (COG-UK) Consortium | Gemma Clark et al |
| EPI_ISL_416482 | R. G. Lugar Center for Public Health Research, National Center for Disease Control and Public Health (NCDC) of Georgia. | R. G. Lugar Center for Public Health Research, National Center for Disease Control and Public Health (NCDC) of Georgia. | Adam Kotorashvili et al |
| EPI_ISL_416481 | R. G. Lugar Center for Public Health Research, National Center for Disease Control and Public Health (NCDC) of Georgia. | R. G. Lugar Center for Public Health Research, National Center for Disease Control and Public Health (NCDC) of Georgia. | Gvantsa Chanturia et al |
| EPI_ISL_416478,EPI_ISL_416479 | R. G. Lugar Center for Public Health Research, National Center for Disease Control and Public Health (NCDC) of Georgia. | R. G. Lugar Center for Public Health Research, National Center for Disease Control and Public Health (NCDC) of Georgia. | Marine Murtskhvaladze et al |
| EPI_ISL_415641,EPI_ISL_415642,EPI_ISL_415644 | R. G. Lugar Center for Public Health Research, National Center for Disease Control and Public Health (NCDC) of Georgia. | R. G. Lugar Center for Public Health Research, National Center for Disease Control and Public Health (NCDC) of Georgia. | Nato Kotaria et al |
| EPI_ISL_637110,EPI_ISL_637112,EPI_ISL_637113 | Rafik Hariri University Hospital | Microbial Pathogenomics Lab | Georgi Merhi et al |
| EPI_ISL_450508,EPI_ISL_450509,EPI_ISL_450511,EPI_ISL_450512 | Rafik Hariri University Hospital | Rafik Hariri University Hospital | Rita Feghali et al |
| EPI_ISL_768616 | Rajavithi Hospital | National Institute of Health, Department of Medical Sciences, Ministry of Public Health, Thailand | Pilailuk Okada et al |
| EPI_ISL_445380,EPI_ISL_447006,EPI_ISL_447028,EPI_ISL_455918,EPI_ISL_693383,EPI_ISL_1073964,EPI_ISL_1138846 | Ramathibodi Hospital | COVID-19 Network Investigations (CONI) Alliance | Elizabeth Batty et al |
| EPI_ISL_768615 | Regional Medical Sciences Center 5 Samut Songkhram | National Institute of Health, Department of Medical Sciences, Ministry of Public Health, Thailand | Pilailuk Okada et al |
| EPI_ISL_708737,EPI_ISL_708807,EPI_ISL_708806,EPI_ISL_708804 | Regional medical sciences center 6 chonburi | National Institute of Health, Department of Medical Sciences, Ministry of Public Health, Thailand | Pilailuk Okada et al |
| EPI_ISL_661271 | Research platform for Transfusion-transmitted Disease, Institute of Blood Transfusion, Chinese Academy of Medical Sciences | Research platform for Transfusion-transmitted Disease, Institute of Blood Transfusion, Chinese Academy of Medical Sciences | He et al |
| EPI_ISL_407071 | Respiratory Virus Unit, Microbiology Services Colindale, Public Health England | Respiratory Virus Unit, Microbiology Services Colindale, Public Health England | Monica Galiano et al |
| EPI_ISL_906851 | Respiratory Viruses Branch, Centers for Disease Control and Prevention | Respiratory Viruses Branch, Centers for Disease Control and Prevention | Tao et al |
| EPI_ISL_430456 | Rizal Medical Center | Research Institute for Tropical Medicine | Medado et al |
| EPI_ISL_779406 | Royal Darwin Hospital Pathology | MDU-PHL | Meumann et al |
| EPI_ISL_1284048 | RS Bhakti Husada Cikarang | Eijkman Institute for Molecular Biology, Ministry of Research and Technology/National Agency for Research and Innovation | Edison Johar et al |
| EPI_ISL_568693 | RS Freeport Tembagapura | Eijkman Institute for Molecular Biology, Ministry of Research and Technology/National Agency for Research and Innovation | Frilasita A Yudhaputri et al |
| EPI_ISL_947290 | RS Hermina Bitung | Eijkman Institute for Molecular Biology, Ministry of Research and Technology/National Agency for Research and Innovation | Hidayat Trimarsanto et al |
| EPI_ISL_833510 | RS Melania, Bogor, Indonesia | Biosafety Level-3 Laboratory, Indonesian Institute of Sciences (LIPI) | Anik Budhi Dharmayanthi et al |

| | | | |
|---|---|---|---|
| EPI_ISL_766036 | RS Mitra Keluarga Bintaro | Eijkman Institute for Molecular Biology, Ministry of Research and Technology/National Agency for Research and Innovation | Frilasita A Yudhaputri et al |
| EPI_ISL_1284314 | RS Mitra Keluarga Kalideres | Eijkman Institute for Molecular Biology, Ministry of Research and Technology/National Agency for Research and Innovation | Edison Johar et al |
| EPI_ISL_1284327 | RS Mitra Keluarga Kalideres | Eijkman Institute for Molecular Biology, Ministry of Research and Technology/National Agency for Research and Innovation | Frilasita A Yudhaputri et al |
| EPI_ISL_833393 | RS MMC Jakarta, Indonesia | Biosafety Level-3 Laboratory, Indonesian Institute of Sciences (LIPI) | Anggia Prasetyoputri et al |
| EPI_ISL_833503 | RS MMC, Jakarta, Indonesia | Biosafety Level-3 Laboratory, Indonesian Institute of Sciences (LIPI) | Anggia Prasetyoputri et al |
| EPI_ISL_833493 | RS PMI Bogor, Indonesia | Biosafety Level-3 Laboratory, Indonesian Institute of Sciences (LIPI) | Isa Nuryana et al |
| EPI_ISL_833506 | RS PMI, Bogor, Indonesia | Biosafety Level-3 Laboratory, Indonesian Institute of Sciences (LIPI) | Isa Nuryana et al |
| EPI_ISL_1117451,EPI_ISL_1137601 | RS PMI, Bogor, West Java | Biosafety Level-3 Laboratory, Indonesian Institute of Sciences (LIPI) | Anik Budhi Dharmayanthi et al |
| EPI_ISL_833502 | RS Sayang Bunda, Makasar, Indonesia | Biosafety Level-3 Laboratory, Indonesian Institute of Sciences (LIPI) | Andri Wardiana et al |
| EPI_ISL_947278 | RS Umum Ciputra Hospital Citragarden City | Eijkman Institute for Molecular Biology, Ministry of Research and Technology/National Agency for Research and Innovation | Hidayat Trimarsanto et al |
| EPI_ISL_454497,EPI_ISL_454501,EPI_ISL_454502 | RSE National Center for Biotechnology"" | RSE National Center for Biotechnology"" | Alexandr Shevtsov et al |
| EPI_ISL_1196005 | RSP Clinic | National Institute for Communicable Diseases of the National Health Laboratory Service | Maphalala GP et al |
| EPI_ISL_947304 | RSU Harapan Bunda | Eijkman Institute for Molecular Biology, Ministry of Research and Technology/National Agency for Research and Innovation | Iskandar Adnan et al |
| EPI_ISL_576383 | RSUD Budi Rahayu Kota Magelang | Genetics Working Group (Pokja Genetik) Faculty of Medicine, Public Health and Nursing Universitas Gadjah Mada (FK-KMK UGM); Disease Investigation Center Wates Ministry of Agriculture Indonesia; Department of Microbiology FK-KMK UGM; Laboratorium Diagnostik Yayasan Tahija World Mosquito Program (WMP) Yogyakarta Center for Tropical Medicine FK-KMK UGM; Integrated Research Center FK-KMK UGM; Department of Computer Science and Electronics FMIPA UGM; Balai Besar Teknik Kesehatan Lingkungan dan Pengendalian Penyakit (BBTKLPP) Yogyakarta | Gunadi et al |
| EPI_ISL_766032 | RSUP Dr. Sardjito | Eijkman Institute for Molecular Biology, Ministry of Research and Technology/National Agency for Research and Innovation | Frilasita A Yudhaputri et al |
| EPI_ISL_491298 | Rural Health Unit - Calauan, Laguna | Research Institute for Tropical Medicine | Tujan et al |
| EPI_ISL_707789,EPI_ISL_1063915,EPI_ISL_1064149 | Rwanda National Reference Laboratory | Rwanda National Reference Laboratory | Enatha Mukantwari et al |
| EPI_ISL_1064153 | Rwanda National Reference Laboratory | Rwanda National Reference Laboratory | ENatha Mukantwari et al |
| EPI_ISL_1138512,EPI_ISL_1138513,EPI_ISL_1138515 | SA Pärnu Hospital Laboratory | 1. Laboratory of Communicable Diseases (Estonia); 2. Eurofins Genomics Europe Sequencing GmbH | Liidia Dotsenko et al |
| EPI_ISL_732961,EPI_ISL_771367 | SA Pathology | SA Pathology | Lex Leong et al |
| EPI_ISL_684881,EPI_ISL_684964,EPI_ISL_690678 | Saitama Prefectural Institute of Public Health | Pathogen Genomics Center, National Institute of Infectious Diseases | Tsuyoshi Sekizuka et al |
| EPI_ISL_749238,EPI_ISL_749474,EPI_ISL_749706,EPI_ISL_750161,EPI_ISL_750162 | Sanatorio Americano | Institut Pasteur de Montevideo | Daiana Mir et al |
| EPI_ISL_738684 | Santa Clara County Public Health Laboratory | Chan-Zuckerberg Biohub | CZB Cliahub Consortium et al |
| EPI_ISL_528747 | Santo Borromeus Hospital | School of Pharmacy & School of Life Sciences and Technology - Institut Teknologi Bandung; Molecular Genetics Laboratory-Faculty of Medicine-Universitas Padjadjaran; Laboratorium Kesehatan Provinsi Jawa Barat | Catur Riani et al |

| | | | |
|---|---|---|---|
| EPI_ISL_528749 | Santosa Hospital Bandung Central | School of Life Sciences and Technology & School of Pharmacy-Institut Teknologi Bandung; Molecular Genetics Laboratory-Faculty of Medicine-Universitas Padjadjaran; Laboratorium Kesehatan Provinsi Jawa Barat | Husna Nugrahapraja et al |
| EPI_ISL_479799 | Sapporo City Institute of Public Health | Pathogen Genomics Center, National Institute of Infectious Diseases | Tsuyoshi Sekizuka et al |
| EPI_ISL_1046762,EPI_ISL_1046763,EPI_ISL_1046765,EPI_ISL_1128143,EPI_ISL_1131134,EPI_ISL_1198826,EPI_ISL_1198827,EPI_ISL_1198843,EPI_ISL_1198846,EPI_ISL_1198847 | SARS-CoV-2 testing team, National Institute of Infectious Diseases | Pathogen Genomics Center, National Institute of Infectious Diseases | Tsuyoshi Sekizuka et al |
| EPI_ISL_1009128,EPI_ISL_1009123,EPI_ISL_1009626,EPI_ISL_1009017,EPI_ISL_1009656,EPI_ISL_1009630,EPI_ISL_1009213,EPI_ISL_1009016,EPI_ISL_1009160 | School of Pharmacy | School of Pharmacy | Ahmed Kandeil et al |
| EPI_ISL_510529 | School of Veterinary Medicine, Disease Control | School of Veterinary Medicine, Disease Control | Simulundu et al |
| EPI_ISL_661189,EPI_ISL_676576 | Scientific Veterinary Institute Novi Sad | Veterinary Specialized Institute Kraljevo", Serbia" | Vidanovic et al |
| EPI_ISL_1015756 | Seattle Flu Study | Seattle Flu Study | Deborah A. Nickerson et al |
| EPI_ISL_1220328 | SELAS ASTRALAB | CNR Virus des Infections Respiratoires - France SUD | Antonin Bal et al |
| EPI_ISL_421514 | Sentinelles network | National Reference Center for Viruses of Respiratory Infections, Institut Pasteur, Paris | Mélanie Albert et al |
| EPI_ISL_593936 | Sentinelles, Fondettes | National Reference Center for Viruses of Respiratory Infections, Institut Pasteur, Paris | Sylvie Behillil et al |
| EPI_ISL_846654 | Servicio de Microbiología Clínica (Complejo Hospitalario de Navarra, Pamplona), Instituto de Investigación Sanitaria de Navarra (IdiSNA) | SeqCOVID-SPAIN consortium/IBV(CSIC) | Carmen Ezpeleta Baquedano et al |
| EPI_ISL_537780,EPI_ISL_537781,EPI_ISL_537785,EPI_ISL_654434 | Servicio de Microbiología, Hospital Miguel Servet, Zaragoza | SeqCOVID-SPAIN consortium/IBV(CSIC) | Antonio Rezusta López et al |
| EPI_ISL_541036 | Servicio de Microbiología, Hospital Miguel Servet, Zaragoza | SeqCOVID-SPAIN consortium/Institute of Biomedicine of Valencia, IBV-CSIC | Antonio Rezusta López et al |
| EPI_ISL_691659 | Servicio de Microbiología, Hospital Universitario Son Espases | SeqCOVID-SPAIN consortium/IBV(CSIC) | Carla López-Causapé et al |
| EPI_ISL_419689 | Servicio de Microbiología. Consorcio Hospital General Universitario de Valencia | Sequencing and Bioinformatics Service and Molecular Epidemiology Research Group. FISABIO-Public Health | Neris Garcia-Gonzalez et al |
| EPI_ISL_420600,EPI_ISL_849181,EPI_ISL_849279,EPI_ISL_849281,EPI_ISL_849286,EPI_ISL_849289,EPI_ISL_849302,EPI_ISL_849350,EPI_ISL_849351,EPI_ISL_856763,EPI_ISL_856768,EPI_ISL_856775,EPI_ISL_856790,EPI_ISL_856799,EPI_ISL_856801 | Servicio Virosis Respiratorias-Departamento Virología-INEI | Instituto Nacional Enfermedades Infecciosas C.G.Malbran | Baumeister E. et al |
| EPI_ISL_529008 | Servizio di igiene e sanità pubblica (SIESP)-Teramo | Istitituto Zooprofilattico Sperimentale dell'Abruzzo e Molise G.Caporale"" | Lorusso A et al |
| EPI_ISL_416334,EPI_ISL_416327,EPI_ISL_416386 | Shanghai Public Health Clinical Center, Shanghai Medical College, Fudan University | National Research Center for Translational Medicine (Shanghai), Ruijin Hospital affiliated to Shanghai Jiao Tong University School of Medicine & Shanghai Public Health Clinical Center | Shengyue Wang et al |
| EPI_ISL_497950 | Shaoxing CDC | Zhejiang Provincial Center for Disease Control and Prevention | Yin Chen et al |
| EPI_ISL_1295691 | Sharp HealthCare Laboratory | Andersen lab at Scripps Research | SEARCH Alliance San Diego with Aaron Harding et al |
| EPI_ISL_582126 | Sheikh Khalifa Medical City | Molecular Surveillance lab Sheikh Khalifa Medical City | Amirtharaj Francis et al |

| | | | |
|---|---|---|---|
| EPI_ISL_582608,EPI_ISL_582614,EPI_ISL_582626,EPI_ISL_582631,EPI_ISL_582633,EPI_ISL_582645,EPI_ISL_582664,EPI_ISL_582667,EPI_ISL_582671,EPI_ISL_582676,EPI_ISL_582685 | Sheikh Khalifa Medical City | Molecular/Surveillance lab Sheikh Khalifa Medical City | Amirtharaj Francis et al |
| EPI_ISL_906091,EPI_ISL_906098 | Shimantik Pathology and Diagnostic Center | Child Health Research Foundation | Senjuti Saha et al |
| EPI_ISL_482678,EPI_ISL_482679,EPI_ISL_482681,EPI_ISL_610153 | Singapore General Hospital | Department of Microbiology | Nurdyana Abdul Rahman et al |
| EPI_ISL_406973 | Singapore General Hospital | National Public Health Laboratory | Mak et al |
| EPI_ISL_407987 | Singapore General Hospital | Programme in Emerging Infectious Diseases, Duke-NUS Medical School | Danielle E Anderson et al |
| EPI_ISL_833338 | Siniloan Rural Health Unit | Research Institute for Tropical Medicine | Hannah Leah Morito et al |
| EPI_ISL_759957 | Siti Khodijah Hospital | Institute of Tropical Disease, Universitas Airlangga | Rima R Prasetya et al |
| EPI_ISL_1143242 | Sonic - Labor Dr. von Froreich GmbH | Robert Koch Institute | et al |
| EPI_ISL_1145074,EPI_ISL_1145118 | Sonic - MVZ Medizinisches Labor Bremen GmbH | Robert Koch Institute | et al |
| EPI_ISL_696289 | Sonora Quest Laboratories, Laboratory Sciences of Arizona | TGen North | Jolene Bowers et al |
| EPI_ISL_672661 | South Eastern Area Laboratory Services (SEALS) | NSW Health Pathology - Institute of Clinical Pathology and Medical Research; Westmead Hospital; University of Sydney | CIDM-PH et al. et al |
| EPI_ISL_1081795 | SOUTH SUPER HI-WAY | Philippine Genome Center | Francis A. Tablizo et al |
| EPI_ISL_849736,EPI_ISL_849737 | Special Operations Medical Research Division, Defence Services Medical Research Centre | Special Operations Medical Research Division, Defence Services Medical Research Centre | Oo et al |
| EPI_ISL_412912 | State Health Office Baden-Württemberg | Charité Universitätsmedizin Berlin, Institute of Virology | Victor M Corman et al |
| EPI_ISL_428901 | State Research Center of Virology and Biotechnology VECTOR, Department of Collection of Microorganisms | State Research Center of Virology and Biotechnology VECTOR, Department of Collection of Microorganisms | Sergey A. Bodnev et al |
| EPI_ISL_747241 | Subang Public Health Office | West Java Health Laboratory; School of Life Sciences and Technology, Institut Teknologi Bandung | Azzania Fibriani et al |
| EPI_ISL_747236 | Sukabumi Public Health | West Java Health Laboratory; School of Life Sciences and Technology, Institut Teknologi Bandung | Azzania Fibriani et al |
| EPI_ISL_450747 | Sunnybrook Health Sciences Centre | Department of Laboratory Medicine and Molecular Diagnostics, Sunnybrook Health Sciences Centre | Jalees A. Nasir et al |
| EPI_ISL_1290501 | Swedish national genomic surveillance program of SARS-CoV-2 | The Public Health Agency of Sweden | Swedish national genomic surveillance program of SARS-CoV-2 et al |
| EPI_ISL_860779 | Swiss National Reference Centre for Influenza | Swiss National Reference Centre for Influenza | Ana Rita Gonçalves Cabecinhas et al |
| EPI_ISL_1151558 | SYNLAB MVZ Ettlingen | Robert Koch Institute | et al |
| EPI_ISL_768522 | Synphaet Hospital | National Institute of Health, Department of Medical Sciences, Ministry of Public Health, Thailand | Pilailuk Okada et al |
| EPI_ISL_411926,EPI_ISL_411927 | Taiwan Centers for Disease Control | Taiwan Centers for Disease Control | Ji-Rong Yang et al |
| EPI_ISL_902922 | Tanjungpura University Hospital | Tanjungpura University Hospital | Andriani et al |
| EPI_ISL_911675 | Tanjungpura University Hospital | Tanjungpura University Hospital | Mahyarudin et al |
| EPI_ISL_966940 | Technical Support Units for Scientific Research (UATRS), National Centre for Scientific and Technical Research (CNRST) | Technical Support Units for Scientific Research (UATRS), National Centre for Scientific and Technical Research (CNRST) | Touil et al |
| EPI_ISL_733573 | Temporary Specimen Collection Centre | Hong Kong Department of Health | Alan K.L. Tsang et al |
| EPI_ISL_1182031 | Tempus | Grubaugh Lab - Yale School of Public Health | Joseph Fauver et al |
| EPI_ISL_1201880 | Tempus | Grubaugh Lab - Yale School of Public Health | Mary Petrone et al |
| EPI_ISL_861754 | Tempus | Grubaugh Lab - Yale School of Public Health | Tara Alpert et al |
| EPI_ISL_1296445 | Thai Red Cross Emerging Infectious Diseases Health Science Centre, Chulalongkorn Hospital, Faculty of Medicine, Chulalongkorn University | Thai Red Cross Emerging Infectious Diseases Center and Faculty of Medicine, Chulalongkorn University | Opass Putcharoen et al |

| | | | |
|---|---|---|---|
| EPI_ISL_956275 | Thai Red Cross Emerging Infectious Diseases Health Science Centre, Chulalongkorn Hospital, Faculty of Medicine, Chulalongkorn University | Thai Red Cross Emerging Infectious Diseases Center and Faculty of Medicine, Chulalongkorn University | Ratima samorn et al |
| EPI_ISL_918166,EPI_ISL_956272,EPI_ISL_984303,EPI_ISL_984304 | Thai Red Cross Emerging Infectious Diseases Health Science Centre, Chulalongkorn Hospital, Faculty of Medicine, Chulalongkorn University | Thai Red Cross Emerging Infectious Diseases Center and Faculty of Medicine, Chulalongkorn University | Rome Buathong et al |
| EPI_ISL_672094 | The Ashley Laboratory, Stanford University | Chan-Zuckerberg Biohub | CZB Cliahub Consortium et al |
| EPI_ISL_756311,EPI_ISL_756357,EPI_ISL_756367 | The Caribbean Public Health Agency | Carrington Lab, Department of PreClinical Sciences, Faculty of Medical Sciences, The University of the West Indies | Nikita S. D. Sahadeo et al |
| EPI_ISL_429080 | The First Affiliated Hospital of Guangzhou Medical University | BGI-shenzhen & The First Affiliated Hospital of Guangzhou Medical University | et al |
| EPI_ISL_429089 | The First Affiliated Hospital of Guangzhou Medical University | BGI-shenzhen & The First Affiliated Hospital of Guangzhou Medical University | Yanqun Wang et al |
| EPI_ISL_1213586 | The Lord's Grace Medical and Industrial Clinic | Philippine Genome Center | Francis A. Tablizo et al |
| EPI_ISL_547966,EPI_ISL_547967,EPI_ISL_577635,EPI_ISL_626580,EPI_ISL_626595,EPI_ISL_693681,EPI_ISL_792696,EPI_ISL_792699,EPI_ISL_850667 | The National Institute of Public Health | State Veterinary Institute Prague | Nagy et al |
| EPI_ISL_513511 | The National Institute of Public Health | The National Institute of Public Health and State Veterinary Institute Prague | Nagy et al |
| EPI_ISL_434572 | The National Institute of Public Health Center for Epidemiology and Microbiology | The National Institute of Public Health Center for Epidemiology and Microbiology | Alexander Nagy et al |
| EPI_ISL_417765,EPI_ISL_829970 | The National University Hospital of Iceland | deCODE genetics | Daniel F Gudbjartsson et al |
| EPI_ISL_891223 | The Oncology Institute Prof. Dr. Ion Chiricuta" Cluj Napoca" | Stefan cel Mare" University Metagenomics Lab" | Lobiuc Andrei et al |
| EPI_ISL_754229,EPI_ISL_754230,EPI_ISL_754231 | The Republican Research and Practical Center for Epidemiology and Microbiology (RRPCEM) | WHO National Influenza Centre Russian Federation | Elena Gasich et al |
| EPI_ISL_906062 | Tilia Laboratories s.r.o. | Tilia Laboratories s.r.o. | Sona Pekova et al |
| EPI_ISL_1196006 | TLC Clinic | National Institute for Communicable Diseases of the National Health Laboratory Service | Maphalala GP et al |
| EPI_ISL_684511 | Tokyo Metropolitan Institute of Public Health | Pathogen Genomics Center, National Institute of Infectious Diseases | Tsuyoshi Sekizuka et al |
| EPI_ISL_586277,EPI_ISL_591105,EPI_ISL_591194,EPI_ISL_755737,EPI_ISL_755738,EPI_ISL_755739,EPI_ISL_755810,EPI_ISL_755889,EPI_ISL_755890,EPI_ISL_755891,EPI_ISL_933658 | Toronto Invasive Bacterial Diseases Network | McMaster University | Allison McGeer et al |
| EPI_ISL_853143 | Toronto Invasive Bacterial Diseases Network | Ontario Institute for Cancer Research | Allison McGeer et al |
| EPI_ISL_717692,EPI_ISL_717694,EPI_ISL_717697,EPI_ISL_717699,EPI_ISL_717700,EPI_ISL_756362,EPI_ISL_756363 | Trinidad Public Health Laboratory | Carrington Lab, Department of PreClinical Sciences, Faculty of Medical Sciences, The University of the West Indies | Nikita S. D. Sahadeo et al |
| EPI_ISL_426631,EPI_ISL_427392,EPI_ISL_436099,EPI_ISL_436108,EPI_ISL_447593,EPI_ISL_457726,EPI_ISL_457733,EPI_ISL_457730 | TSGH-CP molecular lab | TSGH-CP molecular lab | Cherng-Lih Perng et al |
| EPI_ISL_444610 | U.S. Naval Medical Research Center Biological Defense Research Directorate | U.S. Naval Medical Research Center Biological Defense Research Directorate | Voegtly et al |
| EPI_ISL_537521,EPI_ISL_537601 | UCLA Pathology Clinical Microbiology Lab | Kruglyak Lab | Guo et al. et al |
| EPI_ISL_625661 | UCSF Clinical Microbiology Laboratory | Chan-Zuckerberg Biohub | CZB Cliahub Consortium et al |

| | | | |
|---|---|---|---|
| EPI_ISL_737933,EPI_ISL_737941,EPI_ISL_737947,EPI_ISL_737963,EPI_ISL_737967,EPI_ISL_737977,EPI_ISL_737989,EPI_ISL_737990,EPI_ISL_738004,EPI_ISL_738009,EPI_ISL_738012,EPI_ISL_738019,EPI_ISL_738020,EPI_ISL_738021,EPI_ISL_738023,EPI_ISL_738039,EPI_ISL_738040,EPI_ISL_738041 | Uganda Central Public Health Lab and Uganda Virus Research Institute | MRC/UVRI & LSHTM Uganda Research Unit | Matthew Cotten et al |
| EPI_ISL_451185,EPI_ISL_451190,EPI_ISL_451191,EPI_ISL_451192,EPI_ISL_451197 | Uganda Virus Research Institute | MRC/UVRI & LSHTM Uganda Research Unit | Dan Lule Bugembe et al |
| EPI_ISL_628761,EPI_ISL_648124,EPI_ISL_648125,EPI_ISL_861460 | UHAS COVID-19 Lab | UHAS COVID-19 Lab | Kwabena O. Duedu et al |
| EPI_ISL_732993 | UMMC-Health | WHO National Influenza Centre Russian Federation | Andrey Komissarov et al |
| EPI_ISL_1020194 | UMMI Hospital, Bogor, West Java | Biosafety Level-3 Laboratory, Indonesian Institute of Sciences (LIPI) | Anggia Prasetyoputri et al |
| EPI_ISL_738314 | Unilabs Laboratory Medicine | Norwegian Institute of Public Health, Department of Virology | Kathrine Stene-Johansen et al |
| EPI_ISL_1299865,EPI_ISL_1299878 | Unit of lab surveillance of viral emerging diseases, National Lab of Influenza | Respiratory Virus Unit, National Infection Service, Public Health England | PHE Covid Sequencing Team et al |
| EPI_ISL_526277,EPI_ISL_569984,EPI_ISL_570015,EPI_ISL_609810,EPI_ISL_609824 | Unity Health Toronto | Ontario Institute for Cancer Research | Ramzi Fattouh et al |
| EPI_ISL_445317 | UNIV.DE CHILE HOSP.CLINICO | Instituto de Salud Publica de Chile | Andrés E Castillo et al |
| EPI_ISL_525467,EPI_ISL_525468 | Universidad Iberoamericana | International Centre for Genetic Engineering and Biotechnology (ICGEB) and ARGO Open Lab Platform | Robert Paulino-Ramirez et al |
| EPI_ISL_523812 | Universidad Iberoamericana, Instituto de Medicina Tropical & Salud Global | International Centre for Genetic Engineering and Biotechnology (ICGEB) and ARGO Open Lab Platform | Robert Paulino-Ramirez et al |
| EPI_ISL_1272076 | Universidade Federal do Norte do Tocantins (UFNT) | Laboratório de Bioinformática e Biotecnologia (Labinftec/UFT) | Ueric José Borges de Souza et al |
| EPI_ISL_529962 | Universitas Airlangga Hospital | Institute of Tropical Disease, Universitas Airlangga | Jezzy R Dewantari et al |
| EPI_ISL_462990 | University Clinical Centre of the Republic of Srpska | University of Sarajevo, Veterinary Faculty | Teufik et al |
| EPI_ISL_462753 | University Clinical Hospital of Mostar | University of Sarajevo Veterinary Faculty | Goletic et al |
| EPI_ISL_812967,EPI_ISL_812966 | University Clinical Research Center, University of Sciences | University Clinical Research Center, University of Sciences | Diarra et al |
| EPI_ISL_679532,EPI_ISL_1247987 | University College London, Great Ormond Street Hospital for Children NHS Foundation Trust, Imperial College Healthcare NHS Trust | COVID-19 Genomics UK (COG-UK) Consortium | Sergi Castellano et al |
| EPI_ISL_982798 | University Health Network/Mount Sinai Hospital Department of Microbiology | Ontario Institute for Cancer Research | Marie-Ming Aynaud et al |
| EPI_ISL_418275 | University Hospital Basel, Clinical Virology | University Hospital Basel, Clinical Bacteriology | Hirsch et al |
| EPI_ISL_527919,EPI_ISL_581680 | University Hospital Basel, Clinical Virology | University Hospital Basel, Clinical Bacteriology | Madlen Stange et al |
| EPI_ISL_930923,EPI_ISL_931078,EPI_ISL_931189,EPI_ISL_931209,EPI_ISL_931028,EPI_ISL_931222,EPI_ISL_931359 | University Hospital Basel, Clinical Virology | University Hospital Basel, Clinical Bacteriology | Tim Roloff et al |
| EPI_ISL_1272020 | University Hospital Centre Zagreb | Croatian Institute of Public Health | Irena Tabain et al |
| EPI_ISL_710555,EPI_ISL_710556 | University Hospital Dubrava | Ruđer Boškovic Institute; Forensic Science Centre Ivan Vučetić; University of Zagreb Faculty of Science | Robert Beلužić et al |

| | | | |
|---|---|---|---|
| EPI_ISL_847941,EPI_ISL_864644,EPI_ISL_897720,EPI_ISL_897736,EPI_ISL_897779,EPI_ISL_897911,EPI_ISL_897916,EPI_ISL_897931,EPI_ISL_897942,EPI_ISL_897958,EPI_ISL_897959,EPI_ISL_897961,EPI_ISL_897982,EPI_ISL_953683,EPI_ISL_953688,EPI_ISL_953693,EPI_ISL_953703,EPI_ISL_953712,EPI_ISL_953746,EPI_ISL_953747,EPI_ISL_953748,EPI_ISL_953790,EPI_ISL_953792,EPI_ISL_953800,EPI_ISL_953802,EPI_ISL_953827,EPI_ISL_953833,EPI_ISL_953853,EPI_ISL_953908,EPI_ISL_1040366,EPI_ISL_1040368,EPI_ISL_1040409,EPI_ISL_1040418,EPI_ISL_1084969,EPI_ISL_1084987,EPI_ISL_1166653 | University Hospitals of Geneva, Laboratory of Virology | HUG, Laboratory of Virology and the Health2030 Genome Center | Samuel Cordey et al |
| EPI_ISL_775652,EPI_ISL_775722 | University Medical Center Hamburg Eppendorf | Heinrich Pette Institute, Leibniz Institute for Experimental Virology | Alexis Robitaille et al |
| EPI_ISL_1072985 | University of Balamand | Microbial Genomics Lab, Lebanese American University, Byblos | Mira El Chaar et al |
| EPI_ISL_476072,EPI_ISL_476075,EPI_ISL_476077,EPI_ISL_477013 | University of Debrecen, Department of Medical Microbiology | National Laboratory of Virology, Szentágothai Research Centre | Endre Gábor Tóth et al |
| EPI_ISL_896373 | University of Medicine and Pharmacy of Craiova | Stefan cel Mare" University Metagenomics Lab" | Lobiuc Andrei et al |
| EPI_ISL_955173 | University of Sarajevo, Veterinary Faculty, Laboratory for Molecular Diagnostic and Research Laboratory | University of Sarajevo, Veterinary Faculty, Laboratory for Molecular Diagnostic and Research Laboratory | Goletić Š. et al |
| EPI_ISL_1016843 | University of Sarajevo, Veterinary Faculty, Laboratory for Molecular Diagnostic and Research Laboratory | University of Sarajevo, Veterinary Faculty, Laboratory for Molecular Diagnostic and Research Laboratory | Goletic T. et al |
| EPI_ISL_955155 | University of Sarajevo, Veterinary Faculty, Laboratory for Molecular Diagnostic and Research Laboratory | University of Sarajevo, Veterinary Faculty, Laboratory for Molecular Diagnostic and Research Laboratory | Goletić T. et al |
| EPI_ISL_477623,EPI_ISL_677807 | University of Szeged, Institute of Clinical Microbiology | National Laboratory of Virology, Szentágothai Research Centre | Endre Gábor Tóth et al |
| EPI_ISL_977255,EPI_ISL_977256,EPI_ISL_977258,EPI_ISL_977260,EPI_ISL_977270,EPI_ISL_977272,EPI_ISL_977274,EPI_ISL_977276,EPI_ISL_977278,EPI_ISL_977280,EPI_ISL_977284,EPI_ISL_977287,EPI_ISL_977292,EPI_ISL_977296,EPI_ISL_977297,EPI_ISL_977298,EPI_ISL_977335,EPI_ISL_977341,EPI_ISL_977346,EPI_ISL_977347,EPI_ISL_977348,EPI_ISL_977357,EPI_ISL_977383,EPI_ISL_977385,EPI_ISL_977386,EPI_ISL_977391,EPI_ISL_977395,EPI_ISL_977397,EPI_ISL_977398,EPI_ISL_977399,EPI_ISL_977408,EPI_ISL_977410,EPI_ISL_977411,EPI_ISL_977421,EPI_ISL_977460,EPI_ISL_977464,EPI_ISL_977469 | University of Zambia, School of Veterinary Medicine | UNZAVET and PATH | Mulenga Mwenda-Chimfwembe et al |
| EPI_ISL_856695 | University of Zambia, School of Veterinary Medicine, Disease Control | University of Zambia, School of Veterinary Medicine, Disease Control | Simulundu et al |
| EPI_ISL_458285 | unknown | Bundeswehr Institute of Microbiology | Handrick et al |
| EPI_ISL_463006 | unknown | Clinical virology | Fares et al |

| | | | |
|---|---|---|---|
| EPI_ISL_455693 | unknown | Department of Microbiology | Gao et al |
| EPI_ISL_437615,EPI_ISL_437616,EPI_ISL_437617,EPI_ISL_437621,EPI_ISL_437623,EPI_ISL_437624 | unknown | Faculty of Medicine | Rodpan et al |
| EPI_ISL_507020,EPI_ISL_507021 | unknown | Infectious Diseases Research, King Abdullah International Medical Research Center (KAIMRC) | Alghoribi et al |
| EPI_ISL_454152 | unknown | Instituto Nacional de Saude (INSA) | Borges et al et al |
| EPI_ISL_476559 | unknown | Laboratoire Sciences et Technologies de la Santé (STS) Institut Supérieur des Sciences de la Santé Université Hassan 1er, Settat, Morocco | Hajar Lemriss et al |
| EPI_ISL_462434 | unknown | Laboratory Diagnostic | Vidanovic et al |
| EPI_ISL_450409 | unknown | Microbiology | To et al |
| EPI_ISL_434564 | unknown | Microbiology, The University of Hong Kong | To et al |
| EPI_ISL_483064 | unknown | Virology, Ecole Nationale Veterinaire de Toulouse | Bessiere et al |
| EPI_ISL_681952 | UPMC Clinical Microbiology Laboratory | Microbial Genomic Epidemiology Laboratory, University of Pittsburgh | Mustapha M. Mustapha et al |
| EPI_ISL_676525 | Uppsala klinisk mikrobiologi | The Public Health Agency of Sweden | Department of Microbiology et al |
| EPI_ISL_708815,EPI_ISL_708818,EPI_ISL_708817,EPI_ISL_708816 | Urban Institute for Disease Prevention and Control | National Institute of Health, Department of Medical Sciences, Ministry of Public Health, Thailand | Pilailuk Okada et al |
| EPI_ISL_470667 | Utah Public Health Laboratory | Utah Public Health Laboratory | Erin Young et al |
| EPI_ISL_470723,EPI_ISL_487216 | Utah Public Health Laboratory | Utah Public Health Laboratory | Heidi Butz et al |
| EPI_ISL_734931,EPI_ISL_734935,EPI_ISL_734938,EPI_ISL_734941,EPI_ISL_734953,EPI_ISL_734955,EPI_ISL_734972,EPI_ISL_734999,EPI_ISL_735004,EPI_ISL_735049,EPI_ISL_735123,EPI_ISL_735152,EPI_ISL_735187,EPI_ISL_735241,EPI_ISL_735243 | UZ Leuven, National Reference Laboratory for Coronaviruses, Laboratory Medicine, Leuven, Belgium | KU Leuven, Rega Institute, Clinical and Epidemiological Virology | Tony Wawina-Bokalanga et al |
| EPI_ISL_940866 | Vaccines and Infectious Diseases Analytics Research Unit (VIDA) | KRISP, KZN Research Innovation and Sequencing Platform | Baillie Vicky et al |
| EPI_ISL_708820 | Vajira Hospital | National Institute of Health, Department of Medical Sciences, Ministry of Public Health, Thailand | Pilailuk Okada et al |
| EPI_ISL_640097,EPI_ISL_640102 | Vanguard CHC wc VGC | NHLS/UCT | Arash Iranzadeh et al |
| EPI_ISL_710578 | Vardcentralen Brinken | The Public Health Agency of Sweden | Department of Microbiology et al |
| EPI_ISL_1164509,EPI_ISL_1225202 | Vault Health | Minnesota Department of Health, Public Health Laboratory | Alexandra Lorentz et al |
| EPI_ISL_644577 | Veterinary Specialized Institute Kraljevo", Serbia" | Veterinary Specialized Institute Kraljevo", Serbia" | Vidanovic et al |
| EPI_ISL_454795 | Veterinary Specialized Institute Kraljevo | Veterinary Specialized Institute Kraljevo | Vidanovic et al |
| EPI_ISL_708823 | Vibharam Hospital | National Institute of Health, Department of Medical Sciences, Ministry of Public Health, Thailand | Pilailuk Okada et al |
| EPI_ISL_456517 | Victorian Infectious Diseases Reference Laboratory (VIDRL) | Microbiological Diagnostic Unit Public Health Laboratory and Victorian Infectious Diseases Reference Laboratory, Doherty Institute | Caly L. et al |
| EPI_ISL_430626,EPI_ISL_430603 | Victorian Infectious Diseases Reference Laboratory (VIDRL) | Microbiological Diagnostic Unit Public Health Laboratory and Victorian Infectious Diseases Reference Laboratory, The Peter Doherty Institute for Infection and Immunity | Caly L. et al |
| EPI_ISL_419734 | Victorian Infectious Diseases Reference Laboratory (VIDRL) | Victorian Infectious Diseases Reference Laboratory and Microbiological Diagnostic Unit Public Health Laboratory, Doherty Institute | Caly L. et al |
| EPI_ISL_562200,EPI_ISL_562228,EPI_ISL_562231 | Victorian Infectious Diseases Reference Laboratory (VIDRL) | VIDRL and MDU-PHL | Caly et al |
| EPI_ISL_480650,EPI_ISL_480651,EPI_ISL_480700,EPI_ISL_592783,EPI_ISL_640337,EPI_ISL_779605,EPI_ISL_779606,EPI_ISL_854759,EPI_ISL_1249996 | Victorian Infectious Diseases Reference Laboratory (VIDRL) | VIDRL and MDU-PHL | Caly L. et al |
| EPI_ISL_1005231 | Vilnius university hospital Santaros Klinikos, Center of Laboratory Medicine | Vilnius university hospital Santaros Klinikos, Center of Laboratory Medicine | Gytis Dudas et al |

| | | | |
|---|---|---|---|
| EPI_ISL_934018 | Vilnius university hospital Santaros Klinikos, Center of Laboratory Medicine | Vilnius university hospital Santaros Klinikos, Center of Laboratory Medicine | Ingrida Olendraite et al |
| EPI_ISL_560405 | Vilnius University Hospital Santaros Klinikos, Vilnius University | Institute of Biotechnology, Life Sciences Center, Vilnius University and Thermo Fisher Scientific | Justinas Slikas et al |
| EPI_ISL_796574,EPI_ISL_899901,EPI_ISL_1001837,EPI_ISL_1002795,EPI_ISL_1002974,EPI_ISL_1002985,EPI_ISL_1003633,EPI_ISL_1003635,EPI_ISL_1004295,EPI_ISL_1004322,EPI_ISL_1004967,EPI_ISL_1194910,EPI_ISL_1129568,EPI_ISL_1195151,EPI_ISL_1259494 | Viollier AG | Department of Biosystems Science and Engineering, ETH Zürich | Chaoran Chen et al |
| EPI_ISL_728882,EPI_ISL_899913,EPI_ISL_1002389,EPI_ISL_1003335,EPI_ISL_468294,EPI_ISL_468301,EPI_ISL_486447,EPI_ISL_486528,EPI_ISL_498619,EPI_ISL_539418,EPI_ISL_603675,EPI_ISL_1129547,EPI_ISL_1129779,EPI_ISL_1259774 | Viollier AG | Department of Biosystems Science and Engineering, ETH Zürich | Christian Beisel et al |
| EPI_ISL_420849,EPI_ISL_420033,EPI_ISL_420031,EPI_ISL_487369,EPI_ISL_487365,EPI_ISL_513611,EPI_ISL_513612,EPI_ISL_513624,EPI_ISL_514113,EPI_ISL_527506,EPI_ISL_527521,EPI_ISL_591083,EPI_ISL_591084,EPI_ISL_591085,EPI_ISL_965122,EPI_ISL_965123,EPI_ISL_965124 | Viral Respiratory Lab, National Institute for Biomedical Research (INRB) | Pathogen Sequencing Lab, National Institute for Biomedical Research (INRB) | Placide Mbala-Kingebeni et al |
| EPI_ISL_528386 | Viral vaccines, VSVRI- Veterinary serum and vaccine research institute | Viral vaccines, VSVRI- Veterinary serum and vaccine research institute | Saleh et al |
| EPI_ISL_435427 | Virological Research Group, Szentágothai Research Centre | Bioinformatics Research Group, Szentágothai Research Centre | Péter Urbán et al |
| EPI_ISL_679990 | Virology Department, Sheffield Teaching Hospitals NHS Foundation Trust/Department of Infection, Immunity and Cardiovascular Disease, The Medical School, University of Sheffield | COVID-19 Genomics UK (COG-UK) Consortium | Thushan de Silva et al |
| EPI_ISL_1191597,EPI_ISL_1191600,EPI_ISL_1191602,EPI_ISL_1191603,EPI_ISL_1191604,EPI_ISL_1191605 | Virology Department, Victoria Hospital, Plaine-Wilhems, Mauritius | National Institute for Communicable Diseases of the National Health Laboratory Service | Ramuth M et al |
| EPI_ISL_636987 | Virology Lab, National Institute for Biomedical Research (INRB) | Project group Epidemology of Highly Pathogenic Microorganisms, Robert Koch-Institute | Jean-Jacques Muyembe Tamfum et al |
| EPI_ISL_467666 | Virology lab, NIC, NCCD, Ulaanbaatar, Mongolia | National Centre for Communicable Deseases (NCCD) | Naranzul Ts et al |
| EPI_ISL_508862,EPI_ISL_508863,EPI_ISL_625456,EPI_ISL_677634,EPI_ISL_677635,EPI_ISL_677636 | Virology Unit, Institut Pasteur de Madagascar | Virology Unit, Institut Pasteur de Madagascar | Christian Ranaivoson et al |
| EPI_ISL_918373,EPI_ISL_918374,EPI_ISL_918375,EPI_ISL_918376,EPI_ISL_933780,EPI_ISL_933781,EPI_ISL_933784,EPI_ISL_933785 | Virology Unit, Institut Pasteur du Cambodge | Virology Unit, Institut Pasteur du Cambodge | Sokhoun Yann et al |
| EPI_ISL_411902 | Virology Unit, Institut Pasteur du Cambodge. | Virology Unit, Institut Pasteur du Cambodge (Sequencing done by: Jessica E Manning/Jennifer A Bohl at Malaria and Vector Research Research Laboratory, National Institute of Allergy and Infectious Diseases and Vida Ahyong from Chan-Zuckerberg Biohub) | Erik A Karlsson et al |
| EPI_ISL_862814 | Virology, Ecole Nationale Veterinaire de Toulouse | Virology, Ecole Nationale Veterinaire de Toulouse | Bessiere et al |

| | | | |
|---|---|---|---|
| EPI_ISL_890237 | Virology, International Centre for Diarrhoeal Disease Research, Bangladesh (ICDDR,B) | International Centre for Diarrhoeal Disease Research (ICDDR,B) | Hossain et al |
| EPI_ISL_548102 | Waikato Hospital | Institute of Environmental Science and Research (ESR) | Xiaoyun Ren et al |
| EPI_ISL_456398,EPI_ISL_456399 | Wellington SCL | Institute of Environmental Science and Research (ESR) | Matt Storey et al |
| EPI_ISL_1250695 | Wellington SCL (WN) | Institute of Environmental Science and Research (ESR) | Rachel Boyle et al |
| EPI_ISL_1255108,EPI_ISL_1255146,EPI_ISL_1255163,EPI_ISL_1255166,EPI_ISL_1255182,EPI_ISL_1255194,EPI_ISL_1255197,EPI_ISL_1255250,EPI_ISL_1255275 | West African Centre for Cell Biology of Infectious Pathogens (WACCBIP), University of Ghana, Accra, Ghana | West African Centre for Cell Biology of Infectious Pathogens (WACCBIP), University of Ghana, Volta Road, Legon-Accra, Ghana | Collins M. Morang'a et al |
| EPI_ISL_451345 | West China Hospital of Sichuan University | State Key Laboratory of Biotherapy of Sichuan University | Baowen Du et al |
| EPI_ISL_745033 | West Java Health Laboratory | West Java Health Laboratory | Azzania Fibriani et al |
| EPI_ISL_733286,EPI_ISL_1036272 | WHO National Influenza Centre Russian Federation | WHO National Influenza Centre Russian Federation | Andrey Komissarov et al |
| EPI_ISL_860805 | WHO/Minsk | Charité Universitätsmedizin Berlin, Institut für Virologie | Victor M Corman et al |
| EPI_ISL_1196002 | Women correctional | National Institute for Communicable Diseases of the National Health Laboratory Service | Maphalala GP et al |
| EPI_ISL_708813,EPI_ISL_708812 | World Medical Hospital | National Institute of Health, Department of Medical Sciences, Ministry of Public Health, Thailand | Pilailuk Okada et al |
| EPI_ISL_1239697 | WSSE w Krakowie | 1. National Institute of Public Health - National Institute of Hygiene; 2. Eurofins Genomics Europe Sequencing GmbH | Wołkowicz Tomasz et al |
| EPI_ISL_402124 | Wuhan Jinyintan Hospital | Wuhan Institute of Virology, Chinese Academy of Sciences | Peng Zhou et al |
| EPI_ISL_779153,EPI_ISL_779154 | Yale Pathology Lab | Grubaugh Lab - Yale School of Public Health | Tara Alpert et al |
| EPI_ISL_684814 | Yamagata Prefectural Institute of Public Health | Pathogen Genomics Center, National Institute of Infectious Diseases | Tsuyoshi Sekizuka et al |
| EPI_ISL_422425 | Zhejiang Provincial Center for Disease Control and Prevention | Zhejiang Provincial Center for Disease Control and Prevention | Yanjun Zhang et al |
